# People exposed to proton pump inhibitors shortly preceding COVID-19 diagnosis are not at an increased risk of subsequent hospitalizations and mortality: a nation-wide matched cohort study

**DOI:** 10.1101/2022.04.30.22274526

**Authors:** Ivan Kodvanj, Jan Homolak, Vladimir Trkulja

## Abstract

**Aim:** To assess whether exposure to proton pump inhibitors (PPIs) shortly preceding COVID-19 diagnosis affected the risk of subsequent hospitalizations and mortality.

**Methods:** This population-based study embraced first COVID-19 episodes in adults diagnosed up to August 15 2021 in Croatia. Patients were classified based on exposure to PPIs and burden of PPI-requiring morbidities as “non-users” (no issued prescriptions, no recorded treatment-requiring conditions between January 1 2019 and COVID-19 diagnosis), “possible users” (no issued prescriptions, but morbidities present; self-medication possible) and “users” (at least one prescription within 3 months prior to the COVID-19 diagnosis; morbidities present). Subsets were mutually exactly matched for pre-COVID-19 characteristics. The contrast between “users” and “possible users” informed about the effect of PPIs that is separate of the effect of PPI-requiring conditions.

**Results:** Among 433609 patients, “users” and “possible users” were matched 41195 (of 55098) to 17334 (of 18170) in the primary and 33272 to 16434 in the sensitivity analysis. There was no relevant difference between them regarding mortality [primary: RR=0.93 (95%CI 0.85-1.02; RD= - 0.34% (−0.73, 0.03); sensitivity: RR=0.88 (0.78-0.98); RD=-0.45% (−0.80, -0.11)] or hospitalizations [primary: RR=1.04 (0.97-1.13); RD=0.29% (−0.16, 0.73); sensitivity: RR=1.05 (0.97-1.15); RD=0.32% (−0.12, 0.75)]. The risks of both were slightly higher in “possible users” or “users” than in “non-users” (absolutely by ∼0.4%-1.6%) indicating the effect of PPI-requiring morbidities.

**Conclusion:** Pre-morbid exposure to PPIs does not affect the risk of death or hospitalization in adult COVID-19 patients, but PPI-requiring morbidities seemingly slightly increase the risk of both.

**What is already known about this subject?:**

- It appears biologically plausible that pre-morbid exposure to proton pump inhibitors (PPIs) might contribute to unfavorable course of COVID-19 disease
- Two population-based studies in the early pandemics stage indicated no effect of PPIs, while one suggested a higher risk of death/severe COVID-19 disease in PPI pre-exposed subjects

**What this study adds?:**

- Exposure to PPIs directly preceding SarsCov-2 infection has no effect on the risk of subsequent hospitalizations and mortality
- Conditions requiring PPI treatment seem to mildly increase the risk of both

## Introduction

When used rationally, proton pump inhibitors (PPIs) are well tolerated and safe,^1–3^ and are commonly available over-the-counter (OTC).^4^ Long-term PPI use (continuous or intermittent), as required in some peptic acid disorders^2,3^ has been suggested associated with a range of untoward health outcomes,^5,6^. but evidence of a causal PPI effect is highly questionable.^2,7^ Concerns were also raised that PPI use might be an unfavorable factor in respect to COVID-19, by analogy to previously reported associations with other respiratory viral infections.^8^ At the same time, their antiviral effects and potential use *against* SarsCov-2 were also discussed.^9-11^ Biological plausibility for the presumed PPI - COVID-19 relationship is based on reduced antiviral resistance due to prolonged gastrointestinal hypochlorhydria and subsequently increased pulmonary colonization.^5,6,8^ Two questions about pre-COVID PPI use seem to naturally arise from this mechanistic rationale: one pertaining to the risk of infection and the other one to the risk of severe COVID-19.^8^ Four recent systematic reviews^12–15^ identified a number of observational studies dealing with these questions. Among them, as reviewed^12^, five low risk of bias community-based case-control studies referring to periods before mass vaccination consistently showed no association between PPI use and odds of COVID-19 infection. In addition to being medically relevant, this fact informs also about applicable methods in studies focused on PPI exposure and the risk of unfavorable COVID-19 course: they may validly include only COVID-19 infected patients. Otherwise, such studies would need to embrace both infected and non-infected people; by inclusion of only COVID-19 infected patients, they would (in such a case) condition (by selective inclusion) on a variable that is on a path between the exposure (PPIs) and the outcome (COVID-19 course), and this would introduce bias.^16^ As reviewed,^12-15^ most of the studies assessing the relationship between PPI use and COVID-19 outcomes included only hospitalized patients, thus suffering from this very type of bias, as well as from selection bias (the effect in non-hospitalized patients remained unknown).^16^ Three community-based (hence, devoid of such limitations) studies formed matched cohorts (propensity score-based) of “PPI current users” and “never users” among COVID-19 positive patients. In a Danish study,^12^ “current users” (at least one PPI prescription within 90 days prior to the index COVID-19 diagnosis) and “never users” (no such prescriptions over the preceding 15 years) (3955 current vs. 3955 never users) had closely similar risk of a severe disease or death, while the risk of hospitalization was marginally and irrelevantly^12^ higher in “current users” (RR=1.13; 1.03-1.24). In a UK study,^17^ mortality was closely similar in “regular users” and “non-users” (1516 vs. 1516 patients, no definitions provided). In a South Korean study,^18^ the risk of several unfavorable outcomes was higher in “current users” (at least one PPI prescription within 30 days prior to the index COVID-19 diagnosis) than in “never users” (no prescriptions within one year): point odds ratios between 1.63 and 1.90 for 267 vs. 267 patients. In an additional analysis, the odds were particularly higher in “current users” prescribed with PPIs twice daily, and not (or less so) in those with once daily prescriptions.^19^

Owing to these somewhat inconsistent reports, we aimed to address the same question. In doing so, we adopted the following logic: PPIs are not prescribed to treat/prevent COVID-19, but to treat peptic acid disorders and are not prescribed when such conditions are absent. If identification of exposure to PPIs is based only on issued prescriptions, “current users” and “never users” differ in respect to presence/absence of PPI-requiring conditions. No method of confounding control (regression, matching, weighting) can ascertain balance: even if matched on a practically identical propensity score, a “current user” and a “never user” are likely to differ in this respect.^20^ Moreover, a complete “non-exposure” to PPIs/PPI requiring conditions shortly preceding COVID-19 diagnosis is difficult to assume: milder forms of gastrointestinal reflux disease or functional dyspepsia may remain underdiagnosed^21^ and might be self-medicated (OTC PPIs). In order to discern the risk attributable to specifically PPIs from the risk attributable to PPI-requiring conditions, COVID-19 patients prescribed with (and presumably exposed to) PPIs shortly preceding the COVID-19 diagnosis and their non-prescribed peers (controls) should come from the same target population of people suffering peptic acid disorders.

Having this in mind, using a nationwide cohort of adults and adolescents diagnosed with COVID-19 up to August 15 2021, we aimed to estimate whether those prescribed with PPIs shortly preceding the COVID-19 diagnosis differed from their non-prescribed peers regarding subsequent mortality and hospitalizations.

## Subjects and Methods

### Study outline

Anonymized routinely collected data were linked into a database including all subjects in the country diagnosed with COVID-19 between start of the pandemic (February 25, 2020) and August 15, 2021. Linked were data on: date and mode of COVID-19 diagnosis; demographics and COVID-19 vaccination status at diagnosis; medical histories from January 1, 2019 to October 31, 2021, including comorbidities (International Classification of Diseases [ICD-10] codes), all issued prescriptions (Anatomical Therapeutic Chemical codes, ATC) and other medical care, hospital admissions and diagnoses and dates and causes of death.

Having in mind certain country specifics [OTC availability of pantoprazole 20×20 mg tablets, of H_2_ receptor antagonists (extremely rarely prescribed, i.e., <3000 prescriptions issued between January 1, 2019 and October 31, 2021 in the present cohort comprising ∼15% of total population aged ≥years), and of most of the classical nonsteroidal anti-inflammatory drugs (NSAIDs)^22^; as well as inability to link OTC purchases to specific individuals] and our views on the problem of confounding control in the present setting, we defined patient subsets based on timing of issuance of PPI prescriptions as well as the burden of PPI-treatment requiring conditions. The subsets were mutually exactly matched and compared regarding mortality and hospitalizations subsequent to the index COVID-19 diagnosis.

Since anonymized administrative data standardly collected on routine procedures were used in this observational study, ethical approval was waived by the Ethics Committee of the Zagreb University School of Medicine and Croatian Institute for Public Health.

### Patient subsets regarding PPI exposure and comparisons of interest

We defined three patient subsets (details in Table 1): 1) *PPI users* – (i) patients issued at least one prescription for PPIs within 3 months^12^ prior to the index COVID-19 diagnosis (with different number of prescriptions issued over 12 months) (Table 1); and (ii) burdened with respective morbidities/exposure to NSAIDs (Table 1); 2) *PPI possible users* – (i) no PPI/H_2_ receptor antagonist prescriptions issued between January 1 and the index COVID-19 diagnosis, but (ii) burdened with respective morbidities/exposure to NSAIDs (Table 1). Hence, PPI OTC self-medication shortly preceding COVID-19 diagnosis cannot be absolutely excluded; 3) *PPI non-users* – (i) no PPI/H_2_ receptor antagonist prescriptions issued within 12 months prior to the index COVID-19 diagnosis, (ii) no burden of respective morbidities/exposure to NSAIDs (Table 1). Considering the subset characteristics, the most informative about the “effect” of PPIs (i.e., of the fact of being prescribed PPIs) – hence, comparison of primary interest - is that between *PPI users* and *possible users* (Table 1). Other comparisons provide supportive information (Table 1).

**Table 1.**
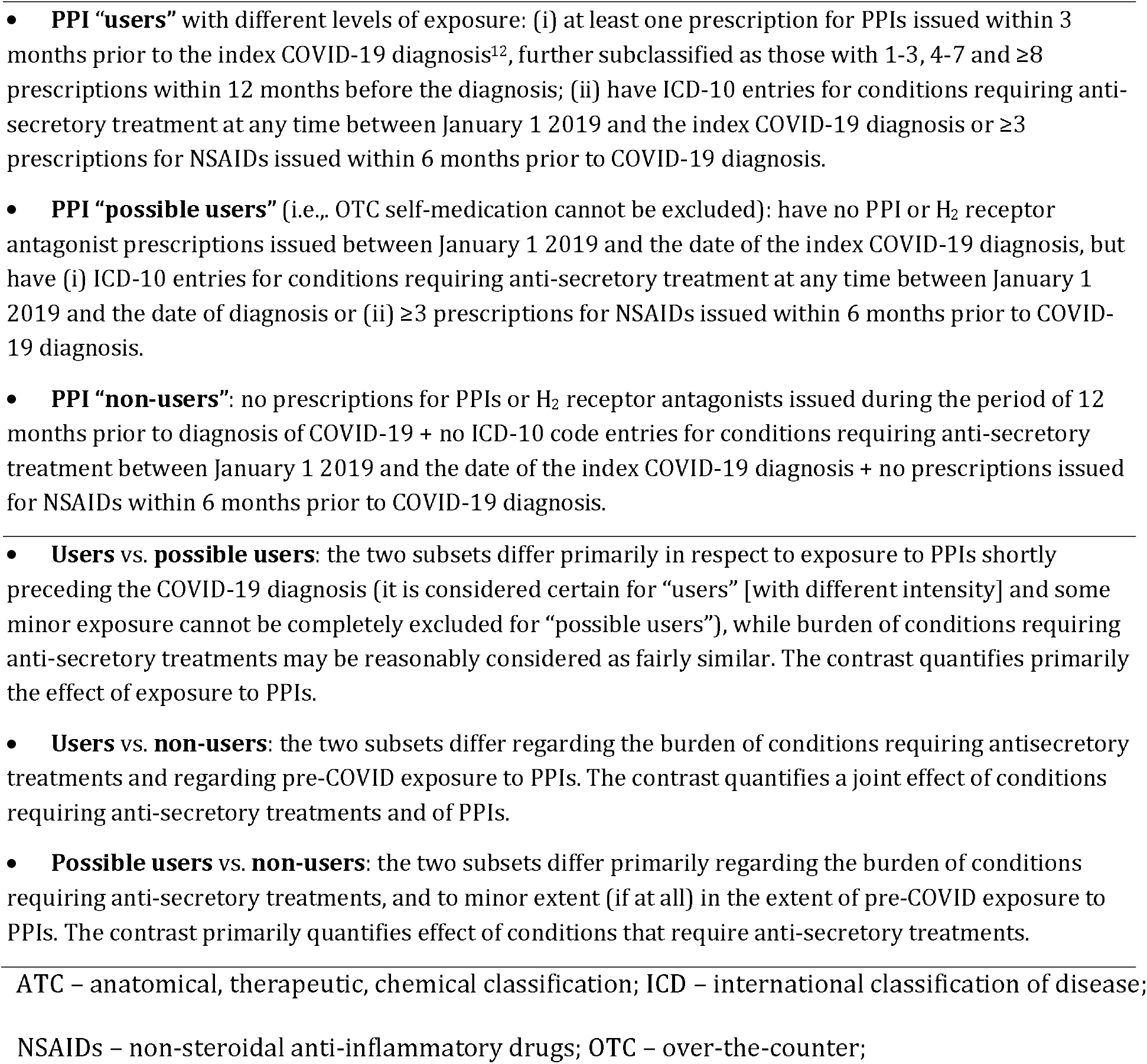
Subsets of patients diagnosed with COVID-19 (up to August 15, 2021) in respect to issuance and timing of proton pump inhibitor (PPI) prescriptions up to the date of the index COVID-19 diagnosis, and contrasts of interest.

### Potential problem of misclassification of “possible users”

We were aware that not all patients would be classifiable into categories of interest and deemed it reasonable to assume that at least some of the “non-classifiable” subjects might have suffered – at certain points between January 1, 2019 and the index COVID-19 diagnosis – milder and commonly underdiagnosed forms of acid peptic disorders. By this virtue, such conditions would remain unregistered, and even if these patients were issued (occasional) PPI prescriptions (and would not meet criteria for “non-users”), they would not qualify for the “possible users” subset – although they could bear similar risks as subjects in the category of “possible users.”“. Therefore, we generated also a matched comparison between *PPI users* and *unclassified* patients.

### Data sources and curation

Raw data were prepared by the Croatian Institute for Public Health (CIPH) from nationwide databases on: (i) COVID-19 laboratory test results (polymerase chain reaction [PCR]-based or rapid antigen tests [RAT]) and COVID-19 patients diagnosed on clinical/epidemiological criteria (without laboratory tests); (ii) COVID-19 vaccinations; (iii) all hospitalizations; (iv) deceased individuals; (v) Central Health Information System (CEZIH) - primary healthcare database maintained by the Ministry of Health.

All subjects diagnosed with COVID-19 (any means) between February 25 2020 and October 15 2021 were identified, and data were linked to the hospitalizations database, database of deceased persons and to their primary healthcare data (January 1 2019 - October 31 2021 for all) (Figure 1A). We received anonymized merged database (Figure 1B) and excluded patients <16 years of age, those for whom data on sex, date of birth, COVID-19 testing date/result/date of diagnosis, or vaccination status/dates were missing or were erroneously entered. We identified subjects with more than one COVID-19 episode: we considered that positive PCR/RAT tests or ICD-10 code U07.1/U07.2 entries or hospitalizations related to COVID-19 that were ≥30 days apart indicated two separate COVID-19 episodes. Only the first documented COVID-19 episode for each subject was included. We set the cut-off date for COVID-19 diagnosis at August 15 2021, to allow for a follow-up period long-enough for outcomes to occur (until October 31). We identified patients subsets, comorbidities and treatments needed for matching as well as the outcomes of interest (Figure 1B).

**Figure 1.**
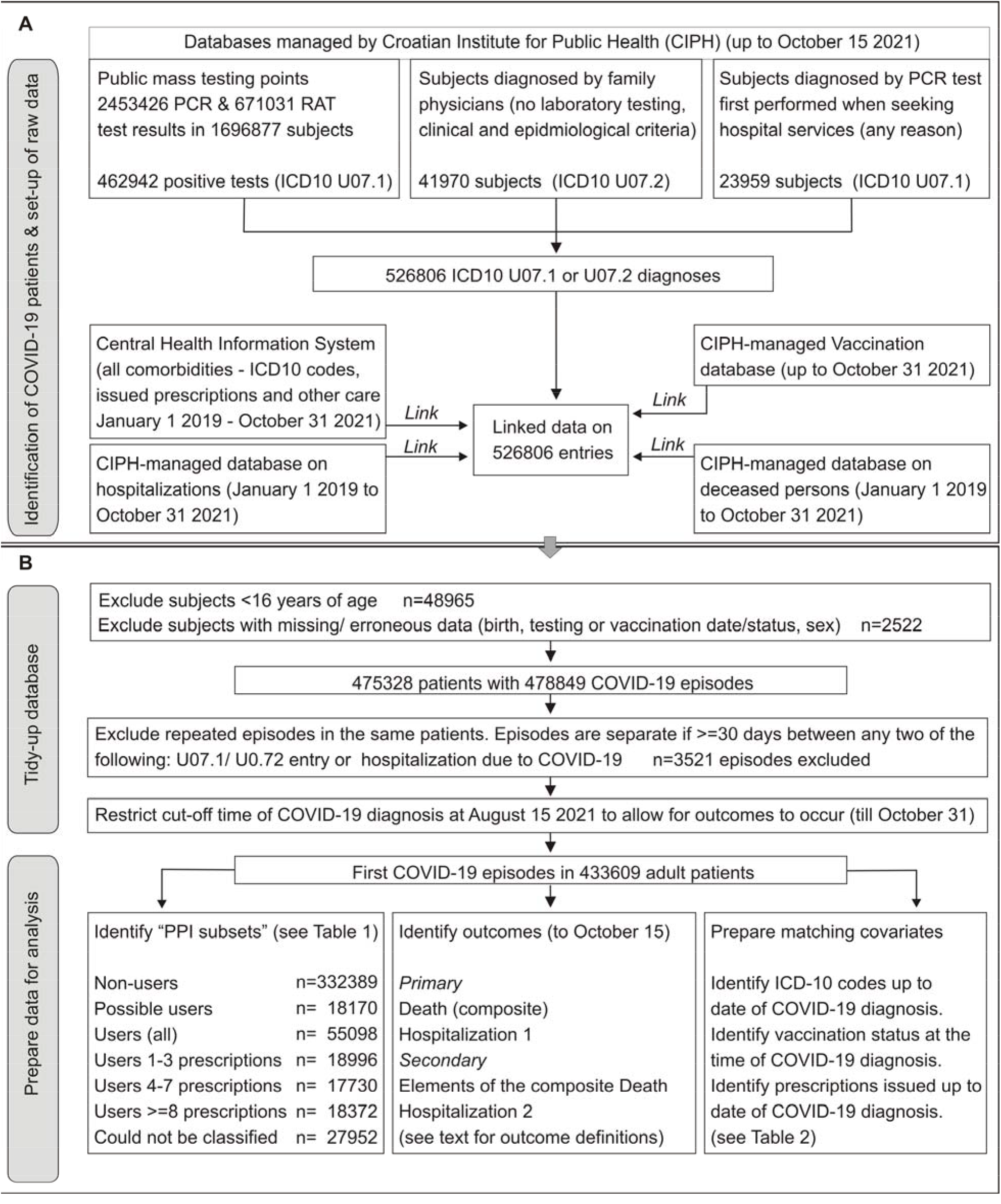
Data sources and curation (see text for details). **A**. Raw data was prepared by the Croatian Institute for Public Health from several databases that it maintains. COVID-19 patients were identified based on positive polymerase chain reaction (PCR) or rapid antigen testing (RAT) performed at dedicated public testing points or hospitals (ICD-10 code U07.1), or based on epidemiological/clinical criteria (ICD-10 code U07.2) and individual data were linked to databases on vaccination, deceased persons, hospitalizations and Central Heath Information System. **B**. Anonymized data were “tidied-up” by exclusion of subjects younger than 16 years and those with missing/erroneous entries on key variables. Also, repeated COVID-19 episodes were excluded and cut-off date for index COVID-19 diagnosis was set at August 15, 2021. Based on International Classification of Disease version 10 (ICD-10) code entries, Anatomical Therapeutic Chemical (ATC) code entries and COVID-19 vaccination status between January 1 2019 and date of COVID-19 diagnosis, subsets of patients in respect to exposure to proton pump inhibitors (PPIs) and health/treatment data relevant for covariate matching were identified. Data on hospitalizations and registry of deceased persons up to October 31 2021 were used to identify outcomes of interest for each subject.

### Identification of treatments, comorbidities and of vaccination status

Exposure to PPIs, H_2_ antagonists and NSAIDs was identified on their respective ATC codes (A02BC, A02BA, M01A). All identified treatments are detailed in Appendix 1 – Table A1. Subjects were considered burdened with anti-secretory treatment-requiring morbidities based on any of the following ICD-10 entries between January 1, 2019 and date of COVID-19 diagnosis: R12, K20, K21, K22, K23, K24, K25, K26, K27, K28, K29, K30, K31. Identification of all other comorbidities is detailed in Appendix 1 – Table A2-Table A4, and section on immunocompromised and diabetic patients. Regarding vaccination, patients were classified as “not vaccinated”, or as: a) vaccinated with a single-dose vaccine; b) received 1^st^ dose of a two-dose vaccine; c) received 2^nd^ (full) dose of a two-dose vaccine; and were further sub-classified based on time elapsed between the last vaccine administration and the index COVID-19 diagnosis (<14 days, 14-90 days and >90 days).

### Outcomes

We defined two *primary outcomes*: a) *Death* as a composite of (i) death occurring after the index COVID-19 diagnosis with U07.1 or U07.2 as the cause of death, regardless of the time elapsed since the COVID-19 diagnosis (minimum follow-up window is 77 days for those diagnosed on August 15, 2021) (Death 1); ii) –death in hospital, where hospitalization followed within 45 days since the index COVID-19 diagnosis with U07.1/U07.2 was the leading diagnosis, or within 30 days of the index COVID-19 diagnosis and U07.1/U07.2 were listed among discharge diagnoses (regardless of immediate cause of death or elapsed time) (Death 2); iii) all-cause death within 14 days since the index COVID-19 diagnosis (Death 3); b) *Hospitalization 1* – hospitalized within 45 days since the index COVID-19 diagnosis with U07.1/U07.2 as the leading diagnosis, or hospitalized within 30 days since the index COVID-19 diagnosis and U07.1/U07.2 is listed among diagnoses. We considered elements of the composite Death outcome and all-cause hospitalization within 14 days since the index COVID-19 diagnosis (*Hospitalization 2*) as secondary outcomes.

### Matching and data analysis

We implemented exact matching in package *MatchIt* ^23^ in R ^24^ with average treatment effect (ATE) as the estimand. Two matched sets were generated for each contrast – one for *primary analysis* and one for a *sensitivity analysis* with some shared and some different covariates used for matching (Table 2). Matched data were analyzed by fitting weighted generalized linear models (distribution=binary) with robust sandwich variance estimator to generate relative risks (RR) (link=log) and risk differences (link=identity). To assess susceptibility of effects of PPI exposure to unmeasured confounding, we calculated E-values (package *Evalue* in R)^25^. Confidence intervals were not adjusted for multiplicity - we considered this a more conservative approach in order not to miss possible untoward effects of exposure to PPIs. In a sensitivity analysis to account for potential misclassification of “possible users”, we applied exact (primary) matching between “users” and “unclassified” patients. Differences between “users” and “unclassified” patients were meta-analytically pooled with the differences between “users” and “possible users” to illustrate the effect of exposure to PPIs. We used random-effects pooling in package *meta* ^26^ in R.

**Table 2.** Covariates used for exact matching in primary and sensitivity analysis (see Appendix 1 for details: Table A1 for ATC codes of pharmacological treatments, Tables A2-A4 for ICD-10 codes of comorbidities, Section 3 and Section 4 on identification of immunocompromised patients and patients with diabetes).

| Matching variables common for primary and sensitivity analysis |  |  |
| --- | --- | --- |
| Age | As 5-year bins between 16 and 111 years |  |
| Sex | Male or female |  |
| Vaccination status | Not vaccinated; received a single-dose vaccine (i) <14 days before diagnosis; ii) 14-90 days before or (iii) >90 days before; received the 1<sup>st</sup> dose of a two-dose vaccine (i) <14 days before; (ii) 14-90 days before or (iii) >90 days before; received the 2<sup>nd</sup> dose of a two-dose vaccine (i) <14 days before; (ii) 14-90 days before or (iii) >90 days before. |  |
| Time period | Up to January 10 2020 – still no vaccination, Alpha strain(s) prevailing; January 10 – July 15 2021 – Alpha strain(s) prevailing, mass vaccination in progress; after July 15 2021 – Delta strain starts to prevail, mass vaccination in progress. |  |
| Comorbidities | Congestive heart failure; malignant disease; renal disease<sup>1</sup> |  |
| Matching variables specific for primary and sensitivity analysis |  |  |
|  | Primary analysis | Sensitivity analysis |
| Comorbidities | Charlson comorbidity index with 4 levels: 0, 1-2, 3-4 or ≥5; and additionally: atrial fibrillation; autoimmune disease; chronic obstructive pulmonary disease; ischemic heart disease or cerebrovascular disease; immunocompromised status (by disease or pharmacologically). | Acute myocardial infarction; peripheral vascular disease; cerebrovascular disease; Alzheimer dementia; other dementia; mild liver disease; moderate to severe liver disease; diabetes without complications; diabetes with complications; metastatic cancer; human immunodeficiency virus infection/AIDS. |
| Co-treatments | RAAS inhibitors (includes angiotensin converting enzyme inhibitors, AT1 receptor antagonists, aliskiren, mineralocorticoid receptor antagonists) | RAAS inhibitors, beta-blockers, diuretics, systemic corticosteroids, other immunosuppressants, antineoplastic drugs, antivirals. |

ATC – anatomical, therapeutic, chemical classification; ICD – international classification of disease;
RAAS – renin angiotensin aldosterone system
<sup>1</sup> Renal disease is included in Charlson comorbidity index (CCI) with extensive codes. In the primary analysis, matching based on CCI levels was supplemented also by exact matching in respect to chronic kidney disease (ICD-10 code N18) and dependence on renal dialysis (ICD-10 code Z99.2). In the sensitivity analysis, renal disease included all individual ICD-10 codes included in CCI

### Nomenclature of targets and ligands

Key protein targets and ligands in this article are hyperlinked to corresponding entries in http://www.guidetopharmacology.org, and are permanently archived in the Concise Guide to PHARMACOLOGY 2019/20 ^27^.

### Results Patients

Raw data (Figure 1A) pertained to 526806 COVID-19 diagnoses (U07.1/U07.2) verified during the index period, eventually resulting in a dataset comprising first COVID-19 episodes in 433609 adults diagnosed up to August 15, 2021 (Figure 1B): 332389 qualified as PPI *non-users*, 18170 were *possible users* and 55098 qualified as *users*, while 27952 (6.4%) could not be classified. The vast majority of the subjects were not even partially vaccinated (Table 3). Age, Charlson comorbidity index and prevalence of all individual comorbidities and treatments used for matching in the primary and sensitivity analysis were the lowest in *non-users* (no PPI exposure, no respective conditions) and increased in *possible users* and *users* (overall) (Table 3), as well as across subsets of *users* with increasing number of issued PPI prescriptions (Table 3). Identical pattern was seen regarding all outcomes (Table 4): mortality and hospitalizations were the lowest in *non-users*, increased in *possible users* and *users*, and across subsets of *users* with increasing number of issued PPI prescriptions (Table 4).

**Table 3.** Subject characteristics (n, %) across subsets based on exposure to PPIs shortly preceding the index COVID-19 diagnosis (see Table 1 for definitions). Shown are all covariates used in primary and sensitivity matching (see Table 2). Vaccination status refers to number of doses received/number for full vaccination and time since the last vaccine dose.

|  | Non users | Possible users | Users (all) | Users, 1-3 | Users, 4-7 | Users, ≥8 |
| --- | --- | --- | --- | --- | --- | --- |
| N | 332389 | 181770 | 55098 | 18996 | 17730 | 18372 |
| Age (years), mean±SD (range) | 44.7±17.1 (16-108) | 53.2±17.9 (16-101) | 61.6±16.4 (16-103) | 55.7±16.8 (16-98) | 62.8±15.6 (16-100) | 66.6±14.8 (16-103) |
| Men | 162983 (49.0) | 7582 (41.7) | 22645 (41.1) | 7707 (40.6) | 7472 (42.1) | 7466 (40.6) |
| Vaccination status |  |  |  |  |  |  |
| 1/1, >90 days | 2 (0.0) | 0 (0.0) | 2 (0.0) | 1 (0.0) | 1 (0.0) | 0 (0.0) |
| 1/1, 14-90 days | 47 (0.0) | 1 (0.0) | 12 (0.0) | 2 (0.0) | 2 (0.0) | 8 (0.0) |
| 1/1, <14 days | 48 (0.0) | 0 (0.0) | 5 (0.0) | 2 (0.0) | 2 (0.0) | 1 (0.0) |
| No vaccination | 322899 (97.1) | 17347 (95.5) | 51111 (92.8) | 18000 (94.8) | 16370 (92.3) | 16741 (91.1) |
| 1/2, >90 days | 35 (0.0) | 7 (0.0) | 22 (0.0) | 7 (0.0) | 8 (0.0) | 7 (0.0) |
| 1/2, 14-90 days | 3549 (1.1) | 295 (1.6) | 1121 (2.0) | 332 (1.7) | 367 (2.1) | 422 (2.3) |
| 1/2, <14 days | 3880 (1.2) | 355 (2.0) | 1791 (3.3) | 431 (2.3) | 621 (3.5) | 739 (4.0) |
| 2/2, >90 days | 363 (0.1) | 30 (0.2) | 239 (0.4) | 36 (0.2) | 76 (0.4) | 127 (0.7) |
| 2/2, 14-90 days | 420 (0.1) | 44 (0.2) | 242 (0.4) | 56 (0.3) | 89 (0.5) | 97 (0.5) |
| 2/2, <14 days | 1146 (0.3) | 91 (0.5) | 553 (1.0) | 129 (0.7) | 194 (1.1) | 230 (1.3) |
| Before January 10, 2021 | 192599 (57.9) | 10566 (58.2) | 32317 (58.7) | 10627 (55.9) | 10501 (59.2) | 11189 (60.9) |
| January 10–July 15, 2021 | 132866 (40.0) | 7257 (39.9) | 22102 (40.1) | 8105 (42.7) | 7015 (39.6) | 6982 (38.0) |
| After July 15, 2021 | 6924 (2.1) | 347 (1.9) | 679 (1.2) | 264 (1.4) | 214 (1.2) | 201 (1.1) |
| Weighted Charlson index |  |  |  |  |  |  |
| 0 | 270536 (81.4) | 11868 (65.3) | 23477 (42.6) | 10660 (56.1) | 7204 (40.6) | 5613 (30.6) |
| 1-2 | 53673 (16.1) | 5248 (28.9) | 21575 (39.2) | 6259 (32.9) | 7253 (40.9) | 8063 (43.9) |
| 3-4 | 6652 (2.0) | 845 (4.7) | 7228 (13.1) | 1527 (8.0) | 2393 (13.5) | 3308 (18.0) |
| ≥5 | 1528 (0.5) | 209 (1.2) | 2818 (5.1) | 550 (2.9) | 880 (5.0) | 1388 (7.6) |
| Atrial fibrillation | 6300 (1.9) | 661 (3.6) | 5687 (10.3) | 1163 (6.1) | 1854 (10.5) | 2670 (14.5) |
| Autoimmune diseases | 19580 (5.9) | 2270 (12.5) | 10034 (18.2) | 2987 (15.7) | 3420 (19.3) | 3627 (19.7) |
| Malignant diseases | 10142 (3.1) | 978 (5.4) | 6253 (11.3) | 1873 (9.9) | 2040 (11.5) | 2340 (12.7) |
| Metastatic cancer | 375 (0.1) | 52 (0.3) | 611 (1.1) | 183 (1.0) | 208 (1.2) | 220 (1.2) |
| Congestive heart failure | 3380 (1.0) | 462 (2.5) | 4234 (7.7) | 783 (4.1) | 1353 (7.6) | 2098 (11.4) |
| COPD | 21520 (6.5) | 1979 (10.9) | 9700 (17.6) | 2626 (13.8) | 3152 (17.8) | 3922 (21.3) |
| IHD or CVD | 12849 (3.9) | 1518 (8.4) | 11683 (21.2) | 2437 (12.8) | 3861 (21.8) | 5385 (29.3) |
| Acute myocardial infarct. | 198 (1.1) | 2103 (0.6) | 2830 (5.1) | 515 (2.7) | 934 (5.3) | 1381 (7.5) |
| Cerebrovascular disease | 5189 (1.6) | 604 (3.3) | 4552 (8.3) | 980 (5.2) | 1451 (8.2) | 2121 (11.5) |
| Renal disease (primary)<sup>1</sup> | 1710 (0.5) | 230 (1.3) | 2580 (4.7) | 409 (2.2) | 864 (4.9) | 1307 (7.1) |
| Renal disease (sensitivity)<sup>1</sup> | 1904 (0.6) | 230 (1.3) | 2580 (4.7) | 409 (2.2) | 864 (4.9) | 1307 (7.1) |
| Immunocompromised | 1463 (0.4) | 132 (0.7) | 1873 (3.4) | 266 (1.4) | 620 (3.5) | 987 (5.4) |
| Peripheral vasc. disease | 2579 (0.8) | 301 (1.7) | 2238 (4.1) | 476 (2.5) | 748 (4.2) | 1014 (5.5) |
| Alzheimer's dementia | 2294 (0.7) | 249 (1.4) | 1540 (2.8) | 266 (1.4) | 461 (2.6) | 813 (4.4) |
| Other dementia | 21520 (6.5) | 1979 (10.9) | 9700 (17.6) | 2626 (13.8) | 3152 (17.8) | 3922 (21.3) |
| Mild liver disease | 3363 (1.0) | 340 (1.9) | 1724 (3.1) | 502 (2.6) | 578 (3.3) | 644 (3.5) |
| Moderate/severe liv. dis. | 66 (0.0) | 7 (0.0) | 127 (0.2) | 33 (0.2) | 43 (0.2) | 51 (0.3) |
| Diabetes w/o complic. | 21896 (6.6) | 2449 (13.5) | 11803 (21.4) | 2729 (14.4) | 3888 (21.9) | 5186 (28.2) |
| Diabetes with complic. | 1735 (0.5) | 202 (1.1) | 1499 (2.7) | 304 (1.6) | 450 (2.5) | 745 (4.1) |
| HIV/AIDS | 21 (0.0) | 1 (0.0) | 8 (0.0) | 2 (0.0) | 4 (0.0) | 2 (0.0) |
| RAAS inhibitors | 43193 (13.0) | 4841 (26.6) | 23376 (42.4) | 5450 (28.7) | 7861 (44.3) | 10065 (54.8) |
| Beta blockers | 22235 (6.7) | 2450 (13.5) | 16969 (30.8) | 3309 (17.4) | 5842 (32.9) | 7818 (42.6) |
| Diuretics | 14387 (4.3) | 1872 (10.3) | 13757 (25.0) | 2567 (13.5) | 4586 (25.9) | 6604 (35.9) |
| Systemic corticosteroids | 617 (0.2) | 72 (0.4) | 2535 (4.6) | 539 (2.8) | 920 (5.2) | 1076 (5.9) |
| Other immunosuppress. | 800 (0.2) | 75 (0.4) | 1454 (2.6) | 203 (1.1) | 560 (3.2) | 691 (3.8) |
| Antineoplastic drugs | 1241 (0.4) | 118 (0.6) | 975 (1.8) | 233 (1.2) | 337 (1.9) | 405 (2.2) |
| Antiviral agents | 215 (0.1) | 16 (0.1) | 417 (0.8) | 80 (0.4) | 150 (0.8) | 187 (1.0) |

<sup>1</sup>See footnote to Table 2 for “renal disease” in primary and sensitivity analysis
COPD – chronic obstructive pulmonary disease, CVD – cerebrovascular disease; IHD – ischemic heart disease; RAAS – any of the inhibitors of the renin angiotensin aldosterone system

**Table 4.** Crude incidence (n, %) of primary and secondary outcomes across patient subsets based on exposure to PPIs shortly preceding the index COVID-19 diagnosis (see Table 1 for definitions).

|  | Non users | Possible users | Users (all) | Users, 1-3 | Users, 4-7 | Users, ≥8 |
| --- | --- | --- | --- | --- | --- | --- |
| N | 332389 | 18170 | 55098 | 18996 | 17730 | 18372 |
| Death (composite) | 5594 (1.7) | 801 (4.4) | 4375 (7.9) | 1064 (5.6) | 1392 (7.9) | 1889 (10.3) |
| Hospitalization 1 | 10905 (3.3) | 1160 (6.4) | 5824 (10.6) | 1646 (8.8) | 1830 (10.3) | 2348 (12.8) |
| Death 1 | 4321 (1.3) | 581 (3.2) | 3196 (5.8) | 760 (4.0) | 1028 (5.8) | 1415 (7.7) |
| Death 2 | 3656 (1.1) | 527 (2.9) | 2865 (5.2) | 703 (3.7) | 922 (5.2) | 1213 (5.2) |
| Death 3 | 4653 (1.4) | 672 (3.7) | 3692 (6.7) | 893 (4.7) | 1170 (6.6) | 1598 (8.7) |
| Hospitalization 2 | 26924 (8.1) | 2671 (14.7) | 12728 (23.1) | 3628 (19.1) | 4096 (23.1) | 5016 (27.3) |

Death (composite) – composite of Death 1 (death occurring after the index COVID-19 diagnosis with U07.1 or U07.2 as a cause of death, regardless of elapsed time), Death 2 (death in hospital, where hospitalization followed within 45 days since the index COVID-19 diagnosis with U07.1/U07.2 as the leading diagnosis, or within 30 days since the index COVID-19 diagnosis and U07.1/U07.2 are listed among diagnoses (regardless of the immediate cause of death or elapsed time) and Death 3 (all-cause death within 14 day since the index COVID-19 diagnosis); Hospitalization 1 – hospitalization within 45 days since the index COVID-19 diagnosis with U07.1/U07.2 as the leading diagnosis, or within 30 days since the index COVID-19 diagnosis and U07.1/U07.2 are listed among diagnoses; Hospitalization 2 – all-cause hospitalization within 14 days since the index COVID-19 diagnosis.

Appendix 2 provides tabulated data on all covariates used in matching in all comparisons – before and after matching, as well as data on all outcomes (Table A5-Table A13 for primary analysis; Table A14-Table A22 for sensitivity analysis).

### Primary outcomes – analysis in matched sets

Relative (RR) and absolute risks (RD) of *death (composite)* were mildly higher in *possible users* and *users* vs. *non-users* (Figure 2A), but in the comparison of *users (all)* to possible users RR and RD were tightly around 1.0/0.0 or were < 1.0/0.0 (Figure 2A), and there was no indication of a “dose”-effect (Figure 2A).

**Figure 2.**
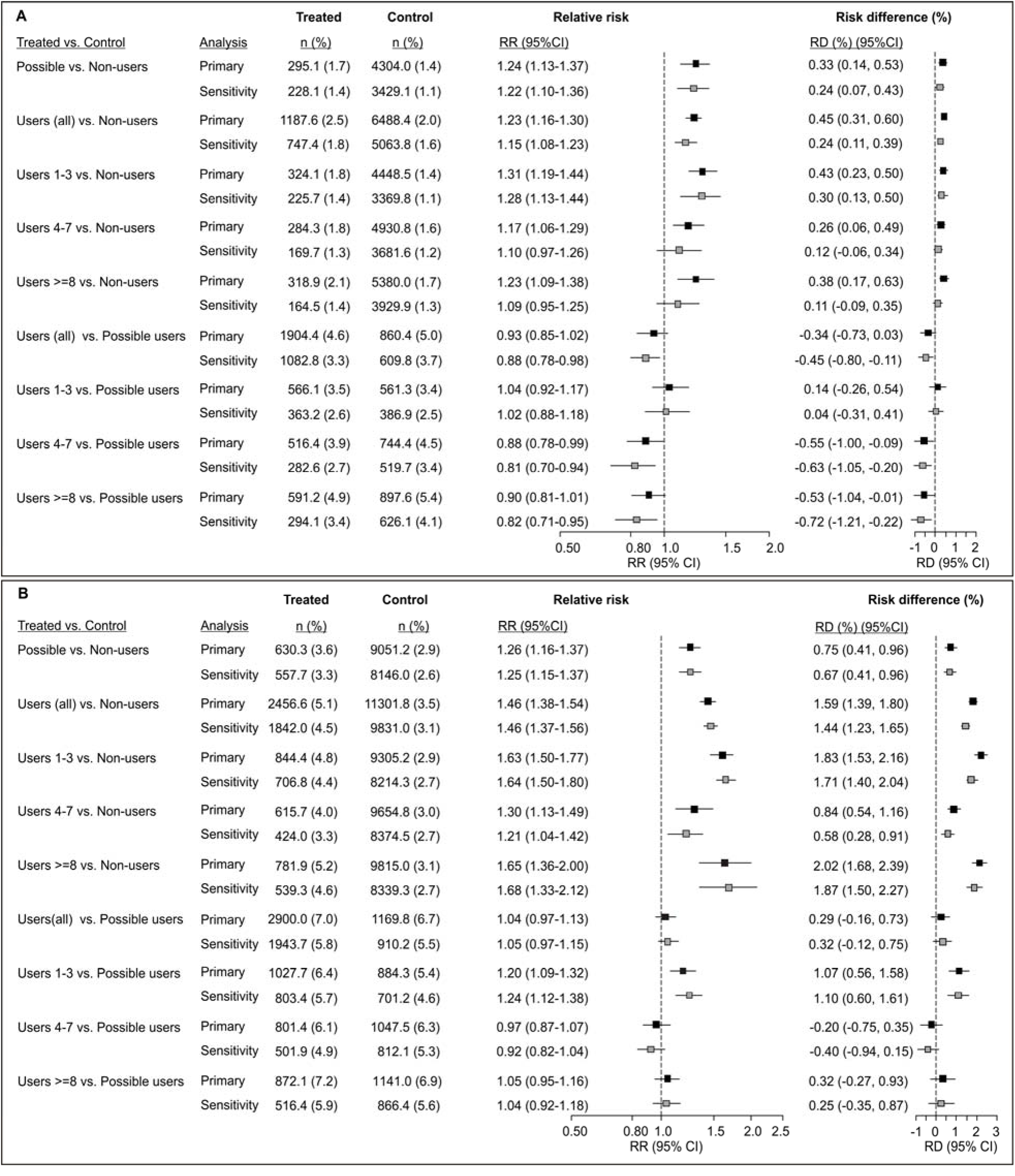
Analysis of the primary outcomes in matched sets: **A**. Death (composite) and **B**. Hospitalization 1 (see Methods for outcome definitions). Contrasts between different patient subsets defined in respect to exposure to proton pump inhibitors (PPIs) and burden of diseases requiring anti-secretory treatments [“non-users”, “possible users”, and “users” with different numbers of issued prescriptions (1-3, 4-7 or ≥8)] inform on different effects (elaborated in Table 1) – the contrast between “users” and “possible users” is the one most informative about the potential effect of exposure to PPIs. Data are weighted counts (%) from matched sets in primary and sensitivity analysis (matching variables are elaborated in Table 2). The numbers of exactly matched patients were (for contrasts displayed from top to bottom): “possible users” vs. “non-users” 17466 to 316168 in the primary and 16795 to 307785 in the sensitivity analysis; “users” (all) vs. “non-users” 48453 to 325005 in the primary and 40653 to 317678 in the sensitivity analysis; “users with 1-3 prescriptions” vs. “non-users” 171734 to 318312 in primary and 16157 to 308555 in the sensitivity analysis; “users with 4-7 prescriptions” vs. “non-users” 15569 to 316577 in primary and 12812 to 306627 in the sensitivity analysis; “users with ≥8 prescriptions” vs. “non-users” 15150 to 313399 in the primary and 11684 to 303530 in the sensitivity analysis; “users (all)” vs. “possible users” 41195 to 17334 in the primary and 33272 to 16343 in the sensitivity analysis; “users with 1-3 prescriptions” vs. “possible users” 15998 to 16517 in the primary and 14205 to 15405 in the sensitivity analysis; “users with 4-7 prescriptions” vs. “possible users” 13115 to 16594 in the primary and 10315 to 15414 in the sensitivity analysis; “users with ≥8 prescriptions” vs. “possible users” 12082 to 16564 in the primary and 8752 to 15337 in the sensitivity analysis. Differences are expressed as relative risks (RR) and as absolute risk (percentage) differences (RD). Estimates (confidence intervals, CI) based on matched data were generated in generalized linear models with robust sandwich variance estimation. Confidence intervals were not adjusted for multiplicity.

Relative (RR) and absolute risks (RD) of *hospitalization 1* were mildly higher in *possible users* and *users* vs. *non-users* (Figure 2B), but in the comparison of *users (all)* to *possible users* RR and RD were tightly around 1.0/0.0 (Figure 2B), and there was no indication of a “dose”-effect (Figure 2B).

### Secondary outcomes – analysis in matched sets

Relative (RR) and absolute risks (RD) of all secondary outcomes were mildly higher in *possible users* and *users* vs. *non-users* (Figure 3A), but in the comparison of *users* to *possible users* all RRs and RDs were tightly around 1.0/0.0 or were <1.0/0.0 (Figure 3A).

**Figure 3.**
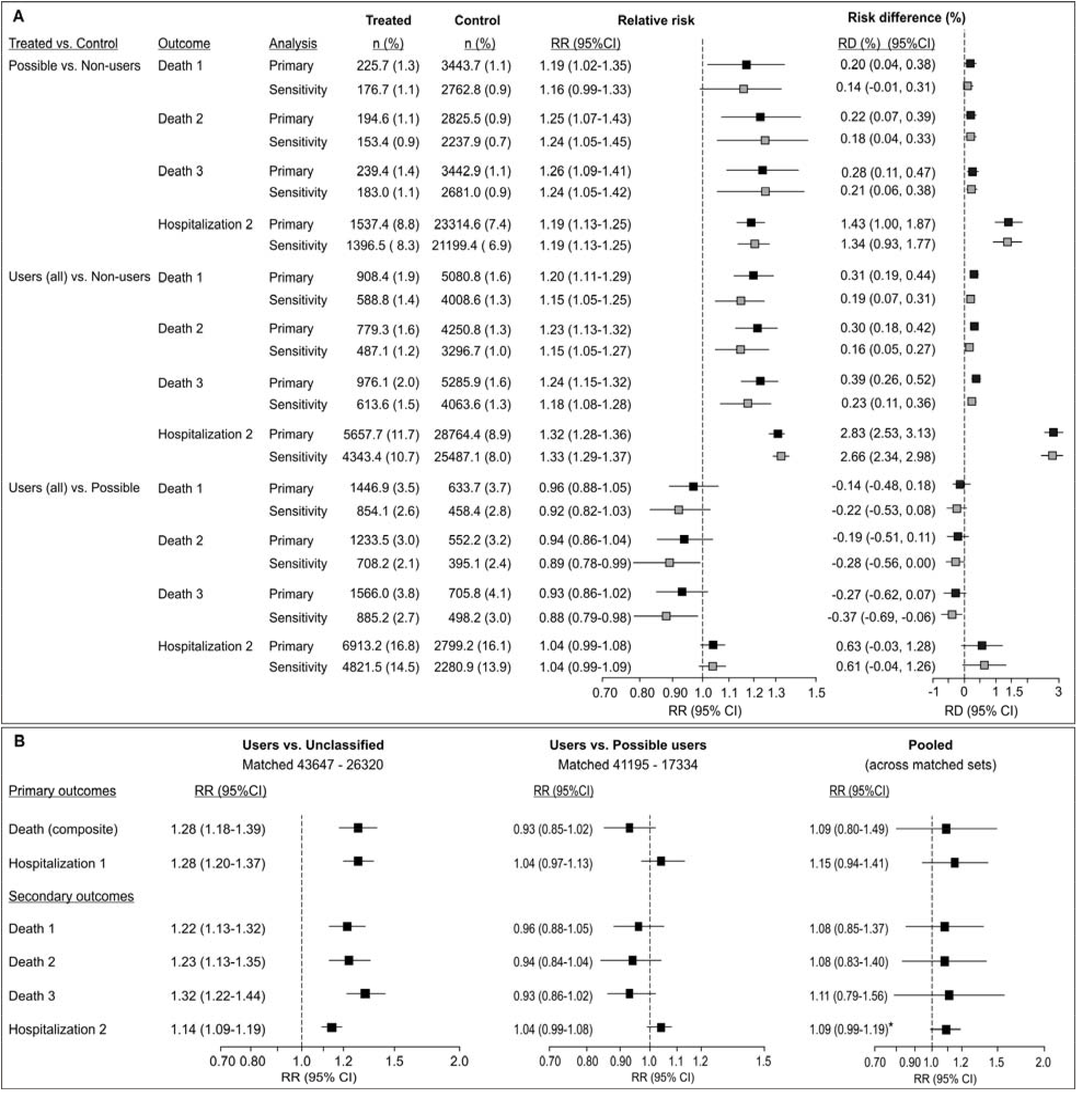
**A**. Analysis of the secondary outcomes in matched sets (see Methods for outcome definitions). Contrasts between different patient subsets defined in respect to exposure to proton pump inhibitors (PPIs) and burden of diseases requiring anti-secretory treatments (“non-users”, “possible users”, and “users”) inform on different effects (elaborated in Table 1) – the contrast between “users” and “possible users” is the one most informative about the potential effect of exposure to PPIs. Data are weighted counts (%) from matched sets in primary and sensitivity analysis (matching variables are elaborated in Table 2). The numbers of exactly matched patients in subset-to-subset comparisons are depicted in footnote to Figure 2d. Differences are expressed as relative risks (RR) and as absolute risk (percentage) differences (RD). Estimates (confidence intervals, CI) based on matched data were generated in generalized linear models with robust sandwich variance estimation. **B**. Differences (as relative risks) between “users” and “unclassified” patients in exactly matched subsets (as for the primary analysis, see Table 2 for covariates) along with differences between “users” and “possible users” in respect to primary and secondary outcomes (see Methods for outcome definitions). Pooled estimates were generated by random-effects pooling (restricted maximum likelihood variance estimator) of the two estimates. Confidence intervals were not adjusted for multiplicity * Numerically, this estimate indicated a trend towards a mildly higher risk in “users”, but already a mild effect (RR=1.25) of unmeasured (and, actually, unknown) confounding would suffice to push the point estimate to 1.04 and lower limit of the CI to 0.95.

### Sensitivity analysis to account for a potential misclassification of possible users

Appendix 2, Table A23, provides tabulated data before and after matching for *users* (all) vs. unclassified patients. In this comparison, RRs for the primary and secondary outcomes somewhat differed from those in the *users* vs. *possible users* comparison (Figure 3B). Due to this heterogeneity, pooled estimates of the *users* vs. unclassified and *users* vs. *possible users* RRs were somewhat imprecise (wide CIs), but also did not indicate any obvious/relevant effect of exposure to PPIs regarding any of the primary or secondary outcomes (Figure 3B).

## Discussion

As recently reviewed, ^12–15^ three out of many studies addressing potential effect of pre-morbid exposure to PPIs on the risk of unfavorable outcomes in COVID-19 patients were population-based ^12,17-19^ and thus devoid of biases common to studies restricted to hospitalized patients.^16^ The present population-based analysis was motivated primarily by their somewhat discrepant results, i.e., increased risk of poorer outcomes (Korean study^18,19^) or no increased risk (Danish study^12^, UK study^17^) in people issued prescriptions for PPIs shortly before the index COVID-19 diagnosis. Next, all three studies^12,17-19^ pertained to the period before vaccine availability, and vaccination could modify the relationship between any potential risk factor and course of the COVID-19 disease. We deemed that observations from different cultural and healthcare settings should contribute to the overall body of evidence on the topic.^28^ In respect to the Danish^12^ and Korean^18-19^ studies, the present analysis is limited in that we focused on relatively “crude” outcomes and did not pursue in detail the issue of a possibly dose-dependent effect of exposure to PPIs (suggested in the Korean study^19^). On the other hand, we believe that the strength of the present analysis is in the use of combined criteria of prescription (non)issuance and presence/history of conditions requiring PPI treatment to define patients “exposed” and “non-exposed” to the presumed risk factor, i.e., treatment with PPIs shortly preceding the COVID-19 diagnosis. Consequently, naming of the patient subsets somewhat differed from the common classification into “current” and “never” users, and, although this might not be immediately intuitive, the comparison of primary interest was that between *users* (at least one PPI prescription within 90 days prior to the index COVID-19 episode ^12^, suffer peptic acid disorders/exposure to NSAIDs) and *possible users* (no PPI/H_2_ antagonist prescriptions within 12 months prior to the index COVID-19 episode, suffer peptic acid disorders/exposure to NSAIDs). We adopted this approach for several reasons. First, the addressed question is not relevant for all COVID-19 patients – focus is on those who, at this critical time preceding the COVID-19 diagnosis, suffer from conditions that need PPI treatment (should it be withheld/postponed?). Thus, both “exposed” (treated) and “non-exposed” (control) patients should come from this target population. In the present setting, this cannot be achieved when the two subsets are identified solely on prescription (non)issuance. Next, if latter is the case, confounding (by indication) is likely to occur and might not be controllable. “Current users” would be suffering from acid peptic disorders, while all or most of their “non-user” controls would not: no exact matching in this respect would be feasible; there would be insufficient overlap between the subsets for any sensible regression modelling, including that for calculation of a propensity score – weighted or matched (on propensity score) “exposed” and “non-exposed” subjects would still differ in this respect.^20,29^ It is not medically implausible to consider that peptic acid disorders could be proxies (or descendants) of unmeasured confounders, or that they could on their own impact the course of the COVID-19 disease. Present data support such a view: crude incidence of all outcomes was higher in older and more comorbid *possible users* than in *non-users* (no PPI/H_2_ prescriptions and no conditions requiring antisecretory treatment within 14 months prior to the index COVID-19 episode) (Table 3, Table 4), and the difference was still present after matching (primary, sensitivity) on a range of covariates (Figure 2, Figure 3). Similarly, the suggested dose-dependent effect of PPI exposure with propensity-score matched “current” and “never” PPI users^19^ might be largely due to confounding by indication. Odds ratio for a composite of hypoxemia, intensive care admission, invasive ventilation or death for “current users” prescribed with once-daily PPIs within 30 days prior to the index COVID-19 diagnosis vs. “never users” was 1.49 (0.87-2.50), and it was 2.36 (1.08-5.10) if “current users” were prescribed with twice daily PPIs.^19^ It is not unreasonable to assume that the observed could have been largely due to the fact that “twice daily users” suffered more severe peptic acid disorders than those with once-daily prescriptions (while “never users” suffered no such conditions). A small confounder effect (e.g., due to bias by indication) of only RR=1.33 would suffice to largely explain away the observed effect (e.g., “move” the lower limit of CI to 0.95).^25^ To the extent to which “intensity” of issued prescriptions over a 12-month period preceding the COVID-19 diagnosis is indicative of a “PPI burden”, present data support such a view: a) crude incidence of all outcomes was progressively higher in *users* with increasing number of issued prescriptions vs. *non-users*, and less so vs. *possible users* (Table 4); b) after matching, the risk of death (composite) or hospitalization was mildly higher in *users* vs. *non-users*, but similarly across subsets with increasing numbers of issued prescriptions (Figure 3), while all RRs for *users* vs. *possible users* were closely around 1.0 (Figure 3). Finally, when “exposed” and “non-exposed” are (in the present setting) identified only based on prescription issuance, an attempt to control for (underlying) peptic acid disorders (e.g., by their inclusion in propensity score calculation) might not only fail to reduce bias, but might worsen it: peptic acid disorders are the main driver for PPI prescription issuance; if they have no impact (or have a very weak impact) on the outcome (apart from that exerted “through” the impact on PPI treatment), then they qualify as instrumental (or near-instrumental) variables – conditioning on them introduces new or amplifies existing bias.^30,31^ Overall, with the method used for patient subsets definition and with exact matching on two comprehensive sets of relevant covariates (taking care to avoid post-baseline variables), we believe that we achieved a reasonable control of confounding. Under these conditions, all comparisons between *users* and *possible users* yielded relative risks for all outcomes that were consistently closely around or lower than unity thus strongly indicating no untoward effect of exposure to PPIs shortly preceding COVID-19 diagnosis on subsequent mortality and hospitalizations. We used routinely collected administrative data and not a dedicated preplanned database, hence some inaccuracies in identification of exposures, comorbidities and outcomes cannot be excluded. In this respect, the following is of notion: a) although data originated from databases regularly maintained by professional organizations (CIPH), we re-checked them using entries on key variables such as dates of birth, sex, vaccination status, dates of COVID-19 test/test results or diagnosis as proxies - only 0.47% of the original entries had missing/erroneous entries, suggesting that inaccuracies/chance errors were sporadic; b) in the national system, prescriptions are issued exclusively within the primary healthcare network, and each prescription bears an ATC code/ICD-10 code. Moreover, for specialist consultations and work-up, patients need to be referred by the primary healthcare physicians who need to record the feedback information. All such acitivities are automatically entered into the Central Health Information System (CEZIH). We left a period of a minimum 14 months (from January 1 2019 to the first COVID-19 case in February 2020) to precede the index COVID-19 diagnosis not to miss entries related to comorbidities that did not require recent prescriptions or other medical procedures. Hence, likely, no relevant comorbidity or treatment was missed; c) having in mind clinical effectiveness of the on-demand PPI use,^32^ we considered that at least one prescription for a 30 tablet pack within 90 days prior to the index COVID-19 diagnosis indicated exposure^12^ in the sense of a reduced gastric acidity at the time of Sars-Cov2 virus infection; d) outcomes were defined in respect to medical relevance, but also in respect to probability of not remaining unrecorded/erroneously recorded, and we left a reasonable period of observation for outcomes to occur (a minimum of 77 days for subjects diagnosed with COVID-19 on August 15, 2021). The observed crude incidence of hospitalizations subsequent to the index COVID-19 diagnosis across the patient subsets were in line with published expected probabilities of hospitalization for people of similar age and comorbidity diagnosed with COVID-19,^33,34^ thus providing a form of external validation of the present data. The overall crude mortality (all subsets and unclassified patients) of 3.0% is in line with the ratio of cumulative COVID-19-confirmed deaths and COVID-19 confirmed cases in Croatia up to October 31, 2021.^35^ Overall, it seems plausible to consider that we reasonably accurately captured relevant exposures, confounders and outcomes, and that inaccuracies/missing data were minor and not prejudiced in respect to the exposure and outcomes of interest.

In conclusion, in the present population-based matched cohort study we observed no signal that people prescribed with PPIs shortly preceding the COVID-19 diagnosis are at increased risk of subsequent mortality or hospitalizations compared to their non-prescribed peers.

## Data Availability

Data can be obtained upon a reasonable request directly from the Croatian institute for public health.

## Disclosures and Declarations

### Ethics

This is an observational study that used anonymized administrative data standardly collected on routine procedures, hence ethical approval was waived by the Ethics Committee of the Zagreb University School of Medicine and Croatian Institute for Public Healthy.

### Author contributions

Ivan Kodvanj, Jan Homolak and Vladimir Trkulja conceived and designed the study. Material preparation, data collection and analysis were performed by Ivan Kodvanj and Vladimir Trkulja. The first draft was written by Ivan Kodvanj and Vladimir Trkulja. All authors commented on previous versions of the manuscripts, read and approved the final version.

## Acknowledgments

Computing was done on high-throughput computing resources (HTC Cloud) provided by the University of Zagreb Computing Centre (SRCE). We are thankful to all the personnel at the Croatian Institute for Public Health (CIPH) for preparing the raw data, and especially for the kind support of Tamara Poljičanin MD, PhD, who supervised the data preparation process at CIPH.

## Data availability

Data can be obtained upon a reasonable request directly from the CIPH.

## Appendix 1

Lists of ATC codes and ICD-10 codes used to identify exposure to pharmacological treatments and comorbidities

## Appendix 2

Tabulated data on covariates used for matching in comparisons between patient subsets – before matching [counts, (%)] and after matching [weighted counts (%)], with standardized mean differences (d)

