## Appendix 1 for "People exposed to proton pump inhibitors shortly preceding COVID-19 diagnosis are not at an increased risk of subsequent hospitalizations and mortality: a nation-wide matched cohort study"

1. Identification of drug prescriptions: **Table A1** – ATC codes used for identification of prescribed drugs
2. Identification of comorbidities: **Table A2** –Charlson comorbidity index (CCI) ICD-10 codes; **Table A3** – weights used for CCI calculation; **Table A4** – Additional comorbidities (ICD-10 codes) used for matching.
3. Identification of immunocompromised patients
4. Identification of patients suffering from diabetes
1. **Identification of drug prescriptions**

**Table A1**. ATC codes used for identification of prescribed drugs (issued prescriptions)

| **Drug** | **ATC^1^** |
| --- | --- |
| Proton pump inhibitors | A02BC |
| H2 receptor inhibitors | A02BA |
| Oral antidiabetic drugs | A10B |
| Insulin | A10A |
| Anticoagulants | BO1AA, B01AB, B01AE, B01AF |
| RAAS inhibitors | C09 |
| Beta blocking agents | C07 |
| Diuretics | C03, C07B, C07C, C09DA, C09DX01, C09DX03 |
| Systemic corticosteroids | H02 |
| Antivirals | J05A |
| Antineoplastic agents | L01, L02, L03 |
| Immunosuppressants | L04 |
| NSAID | M01A |

^1^ Includes all children codes

2. **Identification of comorbidities**

**Table A2.** Charlson comorbidity index codes

| **Charlson comorbidity components** | **ICD10 codes**^1^ |
| --- | --- |
| Myocardial infarction | I21, I22, I25.2 |
| Congestive heart failure | I09.9, I11.0, I13.0, I13.2, I25.5, I42.0, I42.5, I42.6, I42.7, I42.8, I42.9, I43, I50, P29.0 |
| Peripheral vascular disease | I70, I71, I73.1, I73.8, I73.9, I77.1, I79.0, I79.2, K55.1, K55.8, K55.9, Z95.8, Z95.9 |
| Cerebrovascular disease | G45, G46, H34.0, I60, I61, I62, I63, I64, I65, I66, I67, I68, I69 |
| Dementia | F00, F01, F02, F03, F05.1, G30, G31.1 |
| Chronic pulmonary disease | I27.8, I27.9, J40, J41, J42, J43, J44, J45, J46, J47, J60, J61, J62, J63, J64, J65, J66, J67, J68.4, J70.1, J70.3 |
| Rheumatic disease | M05, M06, M31.5, M32, M33, M34, M35.1, M35.3, M36.0 |
| Peptic ulcer disease | K25, K26, K27, K28 |
| Mild liver disease | B18, K70.0, K70.1, K70.2, K70.3, K70.9, K71.3, K71.4, K71.5, K71.7, K73, K74, K76.0, K76.2, K76.3, K76.4, K76.8, K76.9, Z94.4 |
| Diabetes without chronic complications | E10.0, E10.1, E10.6, E10.8, E10.9, E11.0, E11.1, E11.6, E11.8, E11.9, E12.0, E12.1, E12.6, E12.8, E12.9, E13.0, E13.1, E13.6, E13.8, E13.9, E14.0, E14.1, E14.6, E14.8, E14.9 |
| Diabetes with chronic complications | E10.2, E10.3, E10.4, E10.5, E10.7, E11.2, E11.3, E11.4, E11.5, E11.7, E12.2, E12.3, E12.4, E12.5, E12.7, E13.2, E13.3, E13.4, E13.5, E13.7, E14.2, E14.3, E14.4, E14.5, E14.7 |
| Hemiplegia or paraplegia | G04.1, G11.4, G80.1, G80.2, G81, G82, G83.0, G83.1, G83.2, G83.3, G83.4, G83.9 |
| Renal disease | I12.0, I13.1, N03.2, N03.3, N03.4, N03.5, N03.6, N03.7, N05.2, N05.3, N05.4, N05.5, N05.6, N05.7, N18, N19, N25.0, Z49.0, Z49.1, Z49.2, Z94.0, Z99.2 |
| Malignancy | C00, C01, C02, C03, C04, C05, C06, C07, C08, C09, C10, C11, C12, C13, C14, C15, C16, C17, C18, C19, C20, C21, C22, C23, C24, C25, C26, C30, C31, C32, C33, C34, C37, C38, C39, C40, C41, C43, C45, C46, C47, C48, C49, C50, C51, C52, C53, C54, C55, C56, C57, C58, C60, C61, C62, C63, C64, C65, C66, C67, C68, C69, C70, C71, C72, C73, C74, C75, C76, C81, C82, C83, C84, C85, C88, C90, C91, C92, C93, C94, C95, C96, C97 |
| Moderate or severe liver disease | I85.0, I85.9, I86.4, I98.2, K70.4, K71.1, K72.1, K72.9, K76.5, K76.6, K76.7 |
| Metastatic solid tumor | C77, C78, C79, C80 |
| AIDS | B20, B21, B22, B24 |

^1^ All subcodes are included when subcode is not specified

**Table A3.** Weights used for calculation of the Charlson comorbidity index

| **Charlson comorbidity components** | **Weights** |
| --- | --- |
| Myocardial infarction | 1 |
| Congestive heart failure | 1 |
| Peripheral vascular disease | 1 |
| Cerebrovascular disease | 1 |
| Dementia | 1 |
| Chronic pulmonary disease | 1 |
| Rheumatic disease | 1 |
| Peptic ulcer disease | 1 |
| Mild liver disease | 1 |
| Diabetes without chronic complications | 1 |
| Diabetes with chronic complications | 2 |
| Hemiplegia or paraplegia | 2 |
| Renal disease | 2 |
| Malignancy | 2 |
| Moderate or severe liver disease | 3 |
| Metastatic solid tumor | 6 |
| AIDS | 6 |

**Table A4**. Additional comorbidities (ICD10 codes) used for matching

| Covariates for matching | ICD10 codes^1^ |
| --- | --- |
| Atrial fibrilation or undulation | I48 |
| Autoimmune diseases | D86, G35, K50, K52, K73, K74.3, K90.0, L10, L40, L93, L94, M05, M06, M07, M08, M09, M30, M32, M33, M34, M35, M36, M45, M51, N00, N01, N03, N04, N05 |
| Cancer | C00, C01, C02, C03, C04, C05, C06, C07, C08, C09, C10, C11, C12, C13, C14, C15, C16, C17, C18, C19, C20, C21, C22, C23, C24, C25, C26, C30, C31, C32, C33, C34, C37, C38, C39, C40, C41, C43, C45, C46, C47, C48, C49, C50, C51, C52, C53, C54, C55, C56, C57, C58, C60, C61, C62, C63, C64, C65, C66, C67, C68, C69, C70, C71, C72, C73, C74, C75, C76, C77, C78, C79, C80, C81, C82, C83, C84, C85, C88, C90, C91, C92, C93, C94, C95, C96, C97 |
| Congestive heart failure | I09.9, I11.0, I13.0, I13.2, I25.5, I42.0, I42.5, I42.6, I42.7, I42.8, I42.9, I43, I50, P29.0 |
| COPD, asthma, pneumoconioses | I27.8, I27.9, J40, J41, J42, J43, J44, J45, J46, J47, J60, J61, J62, J63, J64, J65, J66, J67, J68.4, J70.1, J70.3 |
| Ishemic or cerebrovascular disease | G45, G46, H34.0, I20, I21, I22, I23, I24, I25, I60, I61, I62, I63, I64, I65, I66, I67, I68, I69 |
| Renal disease | N18, Z99.2 |

^1^ All subcodes are included when subcode is not specified.

3. **Identification of immunocompromised patients**

Patients where considered immunocompromised when meeting any of the following: 1) were diagnosed with HIV/AIDS (ICD10 codes: B20, B21, B22, B24); 2) within 12 months before COVID-19 diagnoses were issued (i) at least 6 prescriptions for antineoplastic drugs; or (ii) at least 9 prescriptions for immunosuppressive drugs; or (iii) at least 9 prescriptions for systemic corticosteroids; 3) received solid organ or stem cells transplants (ICD codes: Z94.0, Z94.1, Z94.2, Z94.3, Z94.4, Z94.6, Z94.8); 4) were maintained on dialysis (ICD10 codes found in primary care records or in hospital records: N18.5, Z99.2); 5) where hospitalized due to cancer (ICD10 codes for cancer used for Charlson comorbidity index in hospital records); 6) had any of the following diagnoses: D45, D46, D47 . Exact codes used to detect prescriptions of antineoplastic, immunosuppressive and systemic corticosteroid drugs are displayed in Table 1.

4. **Identifications of patients suffering from diabetes**

Due to specific rules related to diagnosing diabetes mellitus (DM), patients were assigned diagnosis of diabetes (only) when undergoing measurement of HbA1c. We considered that a patient suffered from diabetes any of the following was met: i) at least two entries of DM diagnosis could be identified between January 1 2019 and the index COVID-19 diagnosis; or ii) one entry of DM diagnosis was identified over the designated period and at least one prescription was issued for either insulin preparations or for oral antidiabetics (for ATC codes, see Table 1).
