## Appendix 2 for "People exposed to proton pump inhibitors shortly preceding COVID-19 diagnosis are not at an increased risk of subsequent hospitalizations and mortality: a nation-wide matched cohort study"

Tabulated data on covariates (listed in Table 2 in the main text) used for matching in comparisons between patient subsets – before matching [counts, (%)] and after matching [weighted counts (%)], with standardized mean differences (d)

**Primary analysis**

Table A5 – possible users vs. non-users

Table A6 – users (all) vs. non-users

Table A7 – users (1-3 prescriptions issued) vs. non-users

Table A8 – users (4-7 prescriptions issued) vs. non-users

Table A9 – users (≥8 prescription issued) vs. non-users

Table A10 – users (all) vs. possible users

Table A11 – users (1-3 prescription issued) vs. possible users

Table A12 – users (4-7 prescriptions issued) vs. possible users

Table A13 –users (≥8 prescriptions issued) vs. possible users

**Sensitivity analysis**

Table A14 – possible users vs. non-users

Table A15 – users (all) vs. non-users

Table A16 – users (1-3 prescriptions issued) vs. non-users

Table A17 – users (4-7 prescriptions issued) vs. non-users

Table A18 – users (≥8 prescription issued) vs. non-users

Table A19 – users (all) vs. possible users

Table A20 – users (1-3 prescription issued) vs. possible users

Table A21 – users (4-7 prescriptions issued) vs. possible users

Table A22 –users (≥8 prescriptions issued) vs. possible users

**Sensitivity analysis to account for potential misclassification of “possible users”**

Table A23 – users (all) vs. unclassified patients

**Table A5**. Primary analysis: possible users vs. non-users (counts/weighted counts, %)

|  | Before matching | | | After matching | | |
| --- | --- | --- | --- | --- | --- | --- |
| Characteristics | Treated | Control | d | Treated | Control | d |
| N | 18170 | 332389 |  | 17466 | 316168 |  |
| Age (5-year bins) |  |  |  |  |  |  |
| [16,21) | 540 ( 3.0) | 22054 ( 6.6) | -0.172 | 1166.6 ( 6.7) | 21117.4 ( 6.7) | 0.000 |
| [21,26) | 759 ( 4.2) | 27711 ( 8.3) | -0.172 | 1468.3 ( 8.4) | 26578.7 ( 8.4) | 0.000 |
| [26,31) | 981 ( 5.4) | 31607 ( 9.5) | -0.157 | 1673.8 ( 9.6) | 30298.2 ( 9.6) | 0.000 |
| [31,36) | 1142 ( 6.3) | 34524 (10.4) | -0.149 | 1839.2 (10.5) | 33292.8 (10.5) | 0.000 |
| [36,41) | 1425 ( 7.8) | 37047 (11.1) | -0.113 | 1973.8 (11.3) | 35729.2 (11.3) | 0.000 |
| [41,46) | 1644 ( 9.0) | 35359 (10.6) | -0.053 | 1899.7 (10.9) | 34388.3 (10.9) | 0.000 |
| [46,51) | 1775 ( 9.8) | 30711 ( 9.2) | 0.018 | 1656.3 ( 9.5) | 29982.7 ( 9.5) | 0.000 |
| [51,56) | 1833 (10.1) | 28177 ( 8.5) | 0.056 | 1517.0 ( 8.7) | 27461.0 ( 8.7) | 0.000 |
| [56,61) | 1886 (10.4) | 25009 ( 7.5) | 0.100 | 1344.4 ( 7.7) | 24335.6 ( 7.7) | 0.000 |
| [61,66) | 1664 ( 9.2) | 19809 ( 6.0) | 0.121 | 1046.0 ( 6.0) | 18934.0 ( 6.0) | 0.000 |
| [66,71) | 1332 ( 7.3) | 14082 ( 4.2) | 0.133 | 708.9 ( 4.1) | 12833.1 ( 4.1) | 0.000 |
| [71,76) | 1010 ( 5.6) | 9476 ( 2.9) | 0.135 | 449.6 ( 2.6) | 8139.4 ( 2.6) | 0.000 |
| [76,81) | 831 ( 4.6) | 6813 ( 2.0) | 0.141 | 308.2 ( 1.8) | 5578.8 ( 1.8) | 0.000 |
| [81,86) | 755 ( 4.2) | 5529 ( 1.7) | 0.149 | 246.5 ( 1.4) | 4462.5 ( 1.4) | 0.000 |
| [86,91) | 430 ( 2.4) | 3158 ( 1.0) | 0.111 | 126.6 ( 0.7) | 2292.4 ( 0.7) | 0.000 |
| [91,96) | 135 ( 0.7) | 1101 ( 0.3) | 0.056 | 36.1 ( 0.2) | 652.9 ( 0.2) | 0.000 |
| [96,101) | 28 ( 0.2) | 214 ( 0.1) | 0.027 | 5.0 ( 0.0) | 91.0 ( 0.0) | 0.000 |
| [101,106) | 0 ( 0.0) | 7 ( 0.0) | -0.006 | 0.0 ( 0.0) | 0.0 ( 0.0) | 0.000 |
| [106,111) | 0 ( 0.0) | 1 ( 0.0) | -0.002 | 0.0 ( 0.0) | 0.0 ( 0.0) | 0.000 |
| Sex |  |  |  |  |  |  |
| M | 7582 (41.7) | 162983 (49.0) | -0.147 | 8457.4 (48.4) | 153095.6 (48.4) | 0.000 |
| F | 10588 (58.3) | 169406 (51.0) | 0.147 | 9008.6 (51.6) | 163072.4 (51.6) | 0.000 |
| Vaccination status |  |  |  |  |  |  |
| 1/1, >90d | 0 ( 0.0) | 2 ( 0.0) | -0.003 | 0.0 ( 0.0) | 0.0 ( 0.0) | 0.000 |
| 1/1, <14d | 1 ( 0.0) | 47 ( 0.0) | -0.009 | 0.1 ( 0.0) | 1.9 ( 0.0) | 0.000 |
| 1/1, 14-90d | 0 ( 0.0) | 48 ( 0.0) | -0.017 | 0.0 ( 0.0) | 0.0 ( 0.0) | 0.000 |
| Not vaccinated | 17347 (95.5) | 322899 (97.1) | -0.089 | 17189.8 (98.4) | 311168.2 (98.4) | 0.000 |
| 1/2, >90d | 7 ( 0.0) | 35 ( 0.0) | 0.018 | 0.0 ( 0.0) | 0.0 ( 0.0) | 0.000 |
| 1/2, <14d | 295 ( 1.6) | 3549 ( 1.1) | 0.048 | 129.1 ( 0.7) | 2337.9 ( 0.7) | 0.000 |
| 1/2, 14-90d | 355 ( 2.0) | 3880 ( 1.2) | 0.063 | 124.1 ( 0.7) | 2246.9 ( 0.7) | 0.000 |
| 2/2, >90d | 30 ( 0.2) | 363 ( 0.1) | 0.015 | 1.2 ( 0.0) | 21.8 ( 0.0) | 0.000 |
| 2/2, <14d | 44 ( 0.2) | 420 ( 0.1) | 0.027 | 5.1 ( 0.0) | 91.9 ( 0.0) | 0.000 |
| 2/2, 14-90d | 91 ( 0.5) | 1146 ( 0.3) | 0.024 | 16.5 ( 0.1) | 299.5 ( 0.1) | 0.000 |
| Period |  |  |  |  |  |  |
| before 10.01.2021. | 10566 (58.2) | 192599 (57.9) | 0.004 | 10304.3 (59.0) | 186527.7 (59.0) | 0.000 |
| 10.01.-15.07.2021. | 7257 (39.9) | 132866 (40.0) | -0.001 | 6866.2 (39.3) | 124290.8 (39.3) | 0.000 |
| after 15.07.2021. | 347 ( 1.9) | 6924 ( 2.1) | -0.012 | 295.5 ( 1.7) | 5349.5 ( 1.7) | 0.000 |
| CC index |  |  |  |  |  |  |
| 0 | 11868 (65.3) | 270536 (81.4) | -0.370 | 14588.9 (83.5) | 264086.1 (83.5) | 0.000 |
| 1-2 | 5248 (28.9) | 53673 (16.1) | 0.309 | 2703.1 (15.5) | 48931.9 (15.5) | 0.000 |
| 3-4 | 845 ( 4.7) | 6652 ( 2.0) | 0.148 | 162.2 ( 0.9) | 2935.8 ( 0.9) | 0.000 |
| >=5 | 209 ( 1.2) | 1528 ( 0.5) | 0.077 | 11.8 ( 0.1) | 214.2 ( 0.1) | 0.000 |
| Comorbidities |  |  |  |  |  |  |
| Atrial fibrillation | 661 ( 3.6) | 6300 ( 1.9) | 0.106 | 160.6 ( 0.9) | 2906.4 ( 0.9) | 0.000 |
| Autoimmune | 2270 (12.5) | 19580 ( 5.9) | 0.230 | 943.2 ( 5.4) | 17073.8 ( 5.4) | 0.000 |
| Cancer | 978 ( 5.4) | 10142 ( 3.1) | 0.116 | 376.0 ( 2.2) | 6806.0 ( 2.2) | 0.000 |
| Congestive heart failure | 462 ( 2.5) | 3380 ( 1.0) | 0.116 | 59.1 ( 0.3) | 1069.9 ( 0.3) | 0.000 |
| COPD | 1979 (10.9) | 21520 ( 6.5) | 0.157 | 1025.0 ( 5.9) | 18554.0 ( 5.9) | 0.000 |
| IHD or CVD | 1518 ( 8.4) | 12849 ( 3.9) | 0.188 | 476.8 ( 2.7) | 8631.2 ( 2.7) | 0.000 |
| Renal disease | 230 ( 1.3) | 1710 ( 0.5) | 0.080 | 19.2 ( 0.1) | 347.8 ( 0.1) | 0.000 |
| Immunocompromised | 132 ( 0.7) | 1463 ( 0.4) | 0.038 | 19.3 ( 0.1) | 349.7 ( 0.1) | 0.000 |
| RAAS inhibitors | 4841 (26.6) | 43193 (13.0) | 0.348 | 2144.4 (12.3) | 38818.6 (12.3) | 0.000 |
| Death (composite) | 803 ( 4.4) | 5599 ( 1.7) | 0.160 | 295.1 ( 1.7) | 4304.0 ( 1.4) | 0.019 |
| Death 1 | 585 ( 3.2) | 4366 ( 1.3) | 0.128 | 225.7 ( 1.3) | 3443.7 ( 1.1) | 0.014 |
| Death 2 | 519 ( 2.9) | 3698 ( 1.1) | 0.125 | 194.6 ( 1.1) | 2825.5 ( 0.9) | 0.016 |
| Death 3 | 666 ( 3.7) | 4519 ( 1.4) | 0.148 | 239.4 ( 1.4) | 3442.9 ( 1.1) | 0.018 |
| Hospitalization 1 | 1160 ( 6.4) | 10905 ( 3.3) | 0.145 | 630.3 ( 3.6) | 9051.2 ( 2.9) | 0.035 |
| Hospitalization 2 | 2680 (14.7) | 26965 ( 8.1) | 0.210 | 1537.4 ( 8.8) | 23314.6 ( 7.4) | 0.045 |

**Table A6**. Primary analysis: users (all) vs. non-users (counts/weighted counts, %)

|  | Before matching | | | After matching | | |
| --- | --- | --- | --- | --- | --- | --- |
| Characteristics | Treated | Control | d | Treated | Control | d |
| N | 55098 | 332389 |  | 48453 | 325005 |  |
| Age |  |  |  |  |  |  |
| [16,21) | 401 ( 0.7) | 22054 ( 6.6) | -0.318 | 2873.1 ( 5.9) | 19271.9 ( 5.9) | 0.000 |
| [21,26) | 782 ( 1.4) | 27711 ( 8.3) | -0.325 | 3655.7 ( 7.5) | 24521.3 ( 7.5) | 0.000 |
| [26,31) | 1137 ( 2.1) | 31607 ( 9.5) | -0.323 | 4193.2 ( 8.7) | 28126.8 ( 8.7) | 0.000 |
| [31,36) | 1703 ( 3.1) | 34524 (10.4) | -0.294 | 4647.1 ( 9.6) | 31170.9 ( 9.6) | 0.000 |
| [36,41) | 2571 ( 4.7) | 37047 (11.1) | -0.242 | 5085.9 (10.5) | 34114.1 (10.5) | 0.000 |
| [41,46) | 3335 ( 6.1) | 35359 (10.6) | -0.166 | 4952.6 (10.2) | 33220.4 (10.2) | 0.000 |
| [46,51) | 4243 ( 7.7) | 30711 ( 9.2) | -0.055 | 4453.1 ( 9.2) | 29869.9 ( 9.2) | 0.000 |
| [51,56) | 5301 ( 9.6) | 28177 ( 8.5) | 0.040 | 4244.9 ( 8.8) | 28473.1 ( 8.8) | 0.000 |
| [56,61) | 6107 (11.1) | 25009 ( 7.5) | 0.123 | 3909.8 ( 8.1) | 26225.2 ( 8.1) | 0.000 |
| [61,66) | 6560 (11.9) | 19809 ( 6.0) | 0.210 | 3256.6 ( 6.7) | 21844.4 ( 6.7) | 0.000 |
| [66,71) | 6134 (11.1) | 14082 ( 4.2) | 0.261 | 2429.8 ( 5.0) | 16298.2 ( 5.0) | 0.000 |
| [71,76) | 5050 ( 9.2) | 9476 ( 2.9) | 0.268 | 1677.2 ( 3.5) | 11249.8 ( 3.5) | 0.000 |
| [76,81) | 4480 ( 8.1) | 6813 ( 2.0) | 0.279 | 1261.9 ( 2.6) | 8464.1 ( 2.6) | 0.000 |
| [81,86) | 4104 ( 7.4) | 5529 ( 1.7) | 0.280 | 1046.6 ( 2.2) | 7020.4 ( 2.2) | 0.000 |
| [86,91) | 2400 ( 4.4) | 3158 ( 1.0) | 0.213 | 579.9 ( 1.2) | 3890.1 ( 1.2) | 0.000 |
| [91,96) | 693 ( 1.3) | 1101 ( 0.3) | 0.105 | 163.1 ( 0.3) | 1093.9 ( 0.3) | 0.000 |
| [96,101) | 95 ( 0.2) | 214 ( 0.1) | 0.031 | 22.4 ( 0.0) | 150.6 ( 0.0) | 0.000 |
| [101,106) | 2 ( 0.0) | 7 ( 0.0) | 0.003 | 0.0 ( 0.0) | 0.0 ( 0.0) | 0.000 |
| [106,111) | 0 ( 0.0) | 1 ( 0.0) | -0.002 | 0.0 ( 0.0) | 0.0 ( 0.0) | 0.000 |
| Sex |  |  |  |  |  |  |
| M | 22645 (41.1) | 162983 (49.0) | -0.160 | 23137.4 (47.8) | 155197.6 (47.8) | 0.000 |
| F | 32453 (58.9) | 169406 (51.0) | 0.160 | 25315.6 (52.2) | 169807.4 (52.2) | 0.000 |
| Vaccination status |  |  |  |  |  |  |
| 1/1, >90d | 2 ( 0.0) | 2 ( 0.0) | 0.007 | 0.0 ( 0.0) | 0.0 ( 0.0) | 0.000 |
| 1/1, <14d | 12 ( 0.0) | 47 ( 0.0) | 0.006 | 0.4 ( 0.0) | 2.6 ( 0.0) | 0.000 |
| 1/1, 14-90d | 5 ( 0.0) | 48 ( 0.0) | -0.005 | 0.0 ( 0.0) | 0.0 ( 0.0) | 0.000 |
| Not vaccinated | 51111 (92.8) | 322899 (97.1) | -0.201 | 47362.7 (97.7) | 317691.3 (97.7) | 0.000 |
| 1/2, >90d | 22 ( 0.0) | 35 ( 0.0) | 0.019 | 0.3 ( 0.0) | 1.7 ( 0.0) | 0.000 |
| 1/2, <14d | 1121 ( 2.0) | 3549 ( 1.1) | 0.078 | 446.7 ( 0.9) | 2996.3 ( 0.9) | 0.000 |
| 1/2, 14-90d | 1791 ( 3.3) | 3880 ( 1.2) | 0.142 | 527.3 ( 1.1) | 3536.7 ( 1.1) | 0.000 |
| 2/2, >90d | 239 ( 0.4) | 363 ( 0.1) | 0.062 | 14.4 ( 0.0) | 96.6 ( 0.0) | 0.000 |
| 2/2, <14d | 242 ( 0.4) | 420 ( 0.1) | 0.059 | 25.9 ( 0.1) | 174.1 ( 0.1) | 0.000 |
| 2/2, 14-90d | 553 ( 1.0) | 1146 ( 0.3) | 0.081 | 75.4 ( 0.2) | 505.6 ( 0.2) | 0.000 |
| Period |  |  |  |  |  |  |
| before 10.01.2021. | 32317 (58.7) | 192599 (57.9) | 0.014 | 28551.6 (58.9) | 191513.4 (58.9) | 0.000 |
| 10.01.-15.07.2021. | 22102 (40.1) | 132866 (40.0) | 0.003 | 19136.9 (39.5) | 128363.1 (39.5) | 0.000 |
| after 15.07.2021. | 679 ( 1.2) | 6924 ( 2.1) | -0.067 | 764.6 ( 1.6) | 5128.4 ( 1.6) | 0.000 |
| CC index |  |  |  |  |  |  |
| 0 | 23477 (42.6) | 270536 (81.4) | -0.872 | 37855.7 (78.1) | 253922.3 (78.1) | 0.000 |
| 1-2 | 21575 (39.2) | 53673 (16.1) | 0.532 | 9159.4 (18.9) | 61437.6 (18.9) | 0.000 |
| 3-4 | 7228 (13.1) | 6652 ( 2.0) | 0.430 | 1237.2 ( 2.6) | 8298.8 ( 2.6) | 0.000 |
| >=5 | 2818 ( 5.1) | 1528 ( 0.5) | 0.286 | 200.7 ( 0.4) | 1346.3 ( 0.4) | 0.000 |
| Comorbidities |  |  |  |  |  |  |
| Atrial fibrillation | 5687 (10.3) | 6300 ( 1.9) | 0.357 | 1056.1 ( 2.2) | 7083.9 ( 2.2) | 0.000 |
| Autoimmune | 10034 (18.2) | 19580 ( 5.9) | 0.385 | 3301.9 ( 6.8) | 22148.1 ( 6.8) | 0.000 |
| Cancer | 6253 (11.3) | 10142 ( 3.1) | 0.325 | 1648.1 ( 3.4) | 11054.9 ( 3.4) | 0.000 |
| Congestive heart failure | 4234 ( 7.7) | 3380 ( 1.0) | 0.331 | 559.1 ( 1.2) | 3749.9 ( 1.2) | 0.000 |
| COPD | 9700 (17.6) | 21520 ( 6.5) | 0.347 | 3502.0 ( 7.2) | 23490.0 ( 7.2) | 0.000 |
| IHD or CVD | 11683 (21.2) | 12849 ( 3.9) | 0.543 | 2520.1 ( 5.2) | 16903.9 ( 5.2) | 0.000 |
| Renal disease | 2580 ( 4.7) | 1710 ( 0.5) | 0.264 | 209.3 ( 0.4) | 1403.7 ( 0.4) | 0.000 |
| Immunocompromised | 1873 ( 3.4) | 1463 ( 0.4) | 0.217 | 194.1 ( 0.4) | 1301.9 ( 0.4) | 0.000 |
| RAAS inhibitors | 23376 (42.4) | 43193 (13.0) | 0.696 | 7816.4 (16.1) | 52429.6 (16.1) | 0.000 |
| Death (composite) | 4356 ( 7.9) | 5599 ( 1.7) | 0.294 | 1187.6 ( 2.5) | 6488.4 ( 2.0) | 0.022 |
| Death 1 | 3195 ( 5.8) | 4366 ( 1.3) | 0.244 | 908.4 ( 1.9) | 5080.8 ( 1.6) | 0.017 |
| Death 2 | 2839 ( 5.2) | 3698 ( 1.1) | 0.234 | 779.3 ( 1.6) | 4250.8 ( 1.3) | 0.017 |
| Death 3 | 3670 ( 6.7) | 4519 ( 1.4) | 0.273 | 976.1 ( 2.0) | 5285.9 ( 1.6) | 0.020 |
| Hospitalization 1 | 5824 (10.6) | 10905 ( 3.3) | 0.290 | 2456.6 ( 5.1) | 11301.8 ( 3.5) | 0.063 |
| Hospitalization 2 | 12735 (23.1) | 26965 ( 8.1) | 0.422 | 5657.7 (11.7) | 28764.4 ( 8.9) | 0.080 |

**Table A7**. Primary analysis: users (1-3 prescriptions) vs. non-users (counts/weighted counts, %)

|  | Before matching | | | After matching | | |
| --- | --- | --- | --- | --- | --- | --- |
| Characteristics | Treated | Control | d | Treated | Control | d |
| N | 18996 | 332389 |  | 17734 | 318312 |  |
| Age (5-year bins) |  |  |  |  |  |  |
| [16,21) | 305 ( 1.6) | 22054 ( 6.6) | -0.255 | 1162.6 ( 6.6) | 20868.4 ( 6.6) | 0.000 |
| [21,26) | 515 ( 2.7) | 27711 ( 8.3) | -0.248 | 1469.2 ( 8.3) | 26371.8 ( 8.3) | 0.000 |
| [26,31) | 726 ( 3.8) | 31607 ( 9.5) | -0.230 | 1678.0 ( 9.5) | 30119.0 ( 9.5) | 0.000 |
| [31,36) | 1002 ( 5.3) | 34524 (10.4) | -0.191 | 1849.8 (10.4) | 33202.2 (10.4) | 0.000 |
| [36,41) | 1378 ( 7.3) | 37047 (11.1) | -0.135 | 2000.5 (11.3) | 35907.5 (11.3) | 0.000 |
| [41,46) | 1596 ( 8.4) | 35359 (10.6) | -0.076 | 1915.7 (10.8) | 34385.3 (10.8) | 0.000 |
| [46,51) | 1825 ( 9.6) | 30711 ( 9.2) | 0.013 | 1677.6 ( 9.5) | 30111.4 ( 9.5) | 0.000 |
| [51,56) | 2101 (11.1) | 28177 ( 8.5) | 0.087 | 1548.4 ( 8.7) | 27792.6 ( 8.7) | 0.000 |
| [56,61) | 2099 (11.0) | 25009 ( 7.5) | 0.122 | 1370.9 ( 7.7) | 24606.1 ( 7.7) | 0.000 |
| [61,66) | 2054 (10.8) | 19809 ( 6.0) | 0.176 | 1082.7 ( 6.1) | 19434.3 ( 6.1) | 0.000 |
| [66,71) | 1680 ( 8.8) | 14082 ( 4.2) | 0.187 | 754.0 ( 4.3) | 13534.0 ( 4.3) | 0.000 |
| [71,76) | 1310 ( 6.9) | 9476 ( 2.9) | 0.189 | 481.3 ( 2.7) | 8639.7 ( 2.7) | 0.000 |
| [76,81) | 1012 ( 5.3) | 6813 ( 2.0) | 0.175 | 327.7 ( 1.8) | 5881.3 ( 1.8) | 0.000 |
| [81,86) | 826 ( 4.3) | 5529 ( 1.7) | 0.158 | 253.8 ( 1.4) | 4555.2 ( 1.4) | 0.000 |
| [86,91) | 433 ( 2.3) | 3158 ( 1.0) | 0.106 | 126.4 ( 0.7) | 2269.6 ( 0.7) | 0.000 |
| [91,96) | 118 ( 0.6) | 1101 ( 0.3) | 0.042 | 33.1 ( 0.2) | 593.9 ( 0.2) | 0.000 |
| [96,101) | 16 ( 0.1) | 214 ( 0.1) | 0.007 | 2.2 ( 0.0) | 39.8 ( 0.0) | 0.000 |
| [101,106) | 0 ( 0.0) | 7 ( 0.0) | -0.006 | 0.0 ( 0.0) | 0.0 ( 0.0) | 0.000 |
| [106,111) | 0 ( 0.0) | 1 ( 0.0) | -0.002 | 0.0 ( 0.0) | 0.0 ( 0.0) | 0.000 |
| Sex |  |  |  |  |  |  |
| M | 7707 (40.6) | 162983 (49.0) | -0.171 | 8565.6 (48.3) | 153746.4 (48.3) | 0.000 |
| F | 11289 (59.4) | 169406 (51.0) | 0.171 | 9168.4 (51.7) | 164565.6 (51.7) | 0.000 |
| Vaccination status |  |  |  |  |  |  |
| 1/1, >90d | 1 ( 0.0) | 2 ( 0.0) | 0.009 | 0.0 ( 0.0) | 0.0 ( 0.0) | 0.000 |
| 1/1, <14d | 2 ( 0.0) | 47 ( 0.0) | -0.003 | 0.2 ( 0.0) | 2.8 ( 0.0) | 0.000 |
| 1/1, 14-90d | 2 ( 0.0) | 48 ( 0.0) | -0.004 | 0.0 ( 0.0) | 0.0 ( 0.0) | 0.000 |
| Not vaccinated | 18000 (94.8) | 322899 (97.1) | -0.121 | 17438.7 (98.3) | 313011.3 (98.3) | 0.000 |
| 1/2, >90d | 7 ( 0.0) | 35 ( 0.0) | 0.017 | 0.0 ( 0.0) | 0.0 ( 0.0) | 0.000 |
| 1/2, <14d | 332 ( 1.7) | 3549 ( 1.1) | 0.058 | 136.8 ( 0.8) | 2456.2 ( 0.8) | 0.000 |
| 1/2, 14-90d | 431 ( 2.3) | 3880 ( 1.2) | 0.085 | 134.2 ( 0.8) | 2408.8 ( 0.8) | 0.000 |
| 2/2, >90d | 36 ( 0.2) | 363 ( 0.1) | 0.021 | 2.0 ( 0.0) | 36.0 ( 0.0) | 0.000 |
| 2/2, <14d | 56 ( 0.3) | 420 ( 0.1) | 0.037 | 4.9 ( 0.0) | 88.1 ( 0.0) | 0.000 |
| 2/2, 14-90d | 129 ( 0.7) | 1146 ( 0.3) | 0.047 | 17.2 ( 0.1) | 308.8 ( 0.1) | 0.000 |
| Period |  |  |  |  |  |  |
| before 10.01.2021. | 10627 (55.9) | 192599 (57.9) | -0.040 | 10424.6 (58.8) | 187114.4 (58.8) | 0.000 |
| 10.01.-15.07.2021. | 8105 (42.7) | 132866 (40.0) | 0.055 | 7019.8 (39.6) | 126000.2 (39.6) | 0.000 |
| after 15.07.2021. | 264 ( 1.4) | 6924 ( 2.1) | -0.053 | 289.6 ( 1.6) | 5197.4 ( 1.6) | 0.000 |
| CC index |  |  |  |  |  |  |
| 0 | 10660 (56.1) | 270536 (81.4) | -0.567 | 14658.9 (82.7) | 263117.1 (82.7) | 0.000 |
| 1-2 | 6259 (32.9) | 53673 (16.1) | 0.398 | 2820.1 (15.9) | 50617.9 (15.9) | 0.000 |
| 3-4 | 1527 ( 8.0) | 6652 ( 2.0) | 0.279 | 226.9 ( 1.3) | 4072.1 ( 1.3) | 0.000 |
| >=5 | 550 ( 2.9) | 1528 ( 0.5) | 0.191 | 28.1 ( 0.2) | 504.9 ( 0.2) | 0.000 |
| Comorbidities |  |  |  |  |  |  |
| Atrial fibrillation | 1163 ( 6.1) | 6300 ( 1.9) | 0.217 | 205.3 ( 1.2) | 3685.7 ( 1.2) | 0.000 |
| Autoimmune | 2987 (15.7) | 19580 ( 5.9) | 0.321 | 997.8 ( 5.6) | 17910.2 ( 5.6) | 0.000 |
| Cancer | 1873 ( 9.9) | 10142 ( 3.1) | 0.280 | 456.9 ( 2.6) | 8201.1 ( 2.6) | 0.000 |
| Congestive heart failure | 783 ( 4.1) | 3380 ( 1.0) | 0.197 | 84.2 ( 0.5) | 1510.8 ( 0.5) | 0.000 |
| COPD | 2626 (13.8) | 21520 ( 6.5) | 0.245 | 1077.1 ( 6.1) | 19333.9 ( 6.1) | 0.000 |
| IHD or CVD | 2437 (12.8) | 12849 ( 3.9) | 0.328 | 565.8 ( 3.2) | 10155.2 ( 3.2) | 0.000 |
| Renal disease | 409 ( 2.2) | 1710 ( 0.5) | 0.143 | 21.2 ( 0.1) | 380.8 ( 0.1) | 0.000 |
| Immunocompromised | 266 ( 1.4) | 1463 ( 0.4) | 0.101 | 27.2 ( 0.2) | 487.8 ( 0.2) | 0.000 |
| RAAS inhibitors | 5450 (28.7) | 43193 (13.0) | 0.394 | 2228.0 (12.6) | 39991.0 (12.6) | 0.000 |
| Death (composite) | 1070 ( 5.6) | 5599 ( 1.7) | 0.211 | 324.1 ( 1.8) | 4448.5 ( 1.4) | 0.023 |
| Death 1 | 761 ( 4.0) | 4366 ( 1.3) | 0.168 | 249.8 ( 1.4) | 3526.4 ( 1.1) | 0.019 |
| Death 2 | 704 ( 3.7) | 3698 ( 1.1) | 0.170 | 211.6 ( 1.2) | 2912.6 ( 0.9) | 0.018 |
| Death 3 | 900 ( 4.7) | 4519 ( 1.4) | 0.197 | 267.6 ( 1.5) | 3558.5 ( 1.1) | 0.023 |
| Hospitalization 1 | 1646 ( 8.7) | 10905 ( 3.3) | 0.229 | 844.4 ( 4.8) | 9305.2 ( 2.9) | 0.078 |
| Hospitalization 2 | 3624 (19.1) | 26965 ( 8.1) | 0.324 | 1931.2 (10.9) | 23947.3 ( 7.5) | 0.100 |

**Table A8**. Primary analysis: users (4-7 prescriptions) vs. non-users (counts/weighted counts, %)

|  | Before matching | | | After matching | | |
| --- | --- | --- | --- | --- | --- | --- |
| Characteristics | Treated | Control | d | Treated | Control | d |
| N | 17730 | 332389 |  | 15569 | 316577 |  |
| Age (5-year bins) |  |  |  |  |  |  |
| [16,21) | 74 ( 0.4) | 22054 ( 6.6) | -0.342 | 973.2 ( 6.3) | 19787.8 ( 6.3) | 0.000 |
| [21,26) | 168 ( 0.9) | 27711 ( 8.3) | -0.357 | 1252.1 ( 8.0) | 25460.9 ( 8.0) | 0.000 |
| [26,31) | 251 ( 1.4) | 31607 ( 9.5) | -0.362 | 1454.5 ( 9.3) | 29576.5 ( 9.3) | 0.000 |
| [31,36) | 442 ( 2.5) | 34524 (10.4) | -0.326 | 1608.6 (10.3) | 32709.4 (10.3) | 0.000 |
| [36,41) | 707 ( 4.0) | 37047 (11.1) | -0.273 | 1724.7 (11.1) | 35069.3 (11.1) | 0.000 |
| [41,46) | 1006 ( 5.7) | 35359 (10.6) | -0.182 | 1663.1 (10.7) | 33817.9 (10.7) | 0.000 |
| [46,51) | 1352 ( 7.6) | 30711 ( 9.2) | -0.058 | 1460.9 ( 9.4) | 29705.1 ( 9.4) | 0.000 |
| [51,56) | 1727 ( 9.7) | 28177 ( 8.5) | 0.044 | 1353.2 ( 8.7) | 27514.8 ( 8.7) | 0.000 |
| [56,61) | 2096 (11.8) | 25009 ( 7.5) | 0.146 | 1214.2 ( 7.8) | 24688.8 ( 7.8) | 0.000 |
| [61,66) | 2205 (12.4) | 19809 ( 6.0) | 0.226 | 969.6 ( 6.2) | 19716.4 ( 6.2) | 0.000 |
| [66,71) | 2108 (11.9) | 14082 ( 4.2) | 0.284 | 692.0 ( 4.4) | 14072.0 ( 4.4) | 0.000 |
| [71,76) | 1709 ( 9.6) | 9476 ( 2.9) | 0.283 | 454.6 ( 2.9) | 9244.4 ( 2.9) | 0.000 |
| [76,81) | 1469 ( 8.3) | 6813 ( 2.0) | 0.285 | 316.0 ( 2.0) | 6426.0 ( 2.0) | 0.000 |
| [81,86) | 1359 ( 7.7) | 5529 ( 1.7) | 0.288 | 255.8 ( 1.6) | 5201.2 ( 1.6) | 0.000 |
| [86,91) | 770 ( 4.3) | 3158 ( 1.0) | 0.213 | 133.7 ( 0.9) | 2719.3 ( 0.9) | 0.000 |
| [91,96) | 247 ( 1.4) | 1101 ( 0.3) | 0.115 | 37.3 ( 0.2) | 757.7 ( 0.2) | 0.000 |
| [96,101) | 40 ( 0.2) | 214 ( 0.1) | 0.042 | 5.4 ( 0.0) | 109.6 ( 0.0) | 0.000 |
| [101,106) | 0 ( 0.0) | 7 ( 0.0) | -0.006 | 0.0 ( 0.0) | 0.0 ( 0.0) | 0.000 |
| [106,111) | 0 ( 0.0) | 1 ( 0.0) | -0.002 | 0.0 ( 0.0) | 0.0 ( 0.0) | 0.000 |
| Sex |  |  |  |  |  |  |
| M | 7472 (42.1) | 162983 (49.0) | -0.139 | 7536.1 (48.4) | 153236.9 (48.4) | 0.000 |
| F | 10258 (57.9) | 169406 (51.0) | 0.139 | 8032.9 (51.6) | 163340.1 (51.6) | 0.000 |
| Vaccination status |  |  |  |  |  |  |
| 1/1, >90d | 1 ( 0.0) | 2 ( 0.0) | 0.009 | 0.0 ( 0.0) | 0.0 ( 0.0) | 0.000 |
| 1/1, <14d | 2 ( 0.0) | 47 ( 0.0) | -0.003 | 0.0 ( 0.0) | 0.0 ( 0.0) | 0.000 |
| 1/1, 14-90d | 2 ( 0.0) | 48 ( 0.0) | -0.003 | 0.0 ( 0.0) | 0.0 ( 0.0) | 0.000 |
| Not vaccinated | 16370 (92.3) | 322899 (97.1) | -0.217 | 15319.1 (98.4) | 311494.9 (98.4) | 0.000 |
| 1/2, >90d | 8 ( 0.0) | 35 ( 0.0) | 0.021 | 0.1 ( 0.0) | 1.9 ( 0.0) | 0.000 |
| 1/2, <14d | 367 ( 2.1) | 3549 ( 1.1) | 0.081 | 107.8 ( 0.7) | 2191.2 ( 0.7) | 0.000 |
| 1/2, 14-90d | 621 ( 3.5) | 3880 ( 1.2) | 0.155 | 126.6 ( 0.8) | 2573.4 ( 0.8) | 0.000 |
| 2/2, >90d | 76 ( 0.4) | 363 ( 0.1) | 0.062 | 2.2 ( 0.0) | 44.8 ( 0.0) | 0.000 |
| 2/2, <14d | 89 ( 0.5) | 420 ( 0.1) | 0.067 | 3.0 ( 0.0) | 62.0 ( 0.0) | 0.000 |
| 2/2, 14-90d | 194 ( 1.1) | 1146 ( 0.3) | 0.089 | 10.3 ( 0.1) | 208.7 ( 0.1) | 0.000 |
| Period |  |  |  |  |  |  |
| before 10.01.2021. | 10501 (59.2) | 192599 (57.9) | 0.026 | 9278.7 (59.6) | 188671.3 (59.6) | 0.000 |
| 10.01.-15.07.2021. | 7015 (39.6) | 132866 (40.0) | -0.008 | 6113.8 (39.3) | 124317.2 (39.3) | 0.000 |
| after 15.07.2021. | 214 ( 1.2) | 6924 ( 2.1) | -0.069 | 176.5 ( 1.1) | 3588.5 ( 1.1) | 0.000 |
| CC index |  |  |  |  |  |  |
| 0 | 7204 (40.6) | 270536 (81.4) | -0.920 | 12760.7 (82.0) | 259474.3 (82.0) | 0.000 |
| 1-2 | 7253 (40.9) | 53673 (16.1) | 0.570 | 2531.9 (16.3) | 51483.1 (16.3) | 0.000 |
| 3-4 | 2393 (13.5) | 6652 ( 2.0) | 0.440 | 243.9 ( 1.6) | 4960.1 ( 1.6) | 0.000 |
| >=5 | 880 ( 5.0) | 1528 ( 0.5) | 0.280 | 32.4 ( 0.2) | 659.6 ( 0.2) | 0.000 |
| Comorbidities |  |  |  |  |  |  |
| Atrial fibrillation | 1854 (10.5) | 6300 ( 1.9) | 0.361 | 216.0 ( 1.4) | 4393.0 ( 1.4) | 0.000 |
| Autoimmune | 3420 (19.3) | 19580 ( 5.9) | 0.412 | 895.2 ( 5.7) | 18202.8 ( 5.7) | 0.000 |
| Cancer | 2040 (11.5) | 10142 ( 3.1) | 0.330 | 403.5 ( 2.6) | 8205.5 ( 2.6) | 0.000 |
| Congestive heart failure | 1353 ( 7.6) | 3380 ( 1.0) | 0.330 | 99.1 ( 0.6) | 2015.9 ( 0.6) | 0.000 |
| COPD | 3152 (17.8) | 21520 ( 6.5) | 0.352 | 959.0 ( 6.2) | 19501.0 ( 6.2) | 0.000 |
| IHD or CVD | 3861 (21.8) | 12849 ( 3.9) | 0.556 | 570.6 ( 3.7) | 11602.4 ( 3.7) | 0.000 |
| Renal disease | 864 ( 4.9) | 1710 ( 0.5) | 0.272 | 34.7 ( 0.2) | 706.3 ( 0.2) | 0.000 |
| Immunocompromised | 620 ( 3.5) | 1463 ( 0.4) | 0.221 | 31.9 ( 0.2) | 649.1 ( 0.2) | 0.000 |
| RAAS inhibitors | 7861 (44.3) | 43193 (13.0) | 0.739 | 2108.7 (13.5) | 42877.3 (13.5) | 0.000 |
| Death (composite) | 1397 ( 7.9) | 5599 ( 1.7) | 0.293 | 284.3 ( 1.8) | 4930.8 ( 1.6) | 0.013 |
| Death 1 | 1022 ( 5.8) | 4366 ( 1.3) | 0.243 | 213.3 ( 1.4) | 3908.5 ( 1.2) | 0.007 |
| Death 2 | 920 ( 5.2) | 3698 ( 1.1) | 0.235 | 193.2 ( 1.2) | 3241.1 ( 1.0) | 0.013 |
| Death 3 | 1170 ( 6.6) | 4519 ( 1.4) | 0.270 | 230.0 ( 1.5) | 3973.6 ( 1.3) | 0.011 |
| Hospitalization 1 | 1830 (10.3) | 10905 ( 3.3) | 0.282 | 615.7 ( 4.0) | 9654.8 ( 3.0) | 0.036 |
| Hospitalization 2 | 4096 (23.1) | 26965 ( 8.1) | 0.422 | 1539.5 ( 9.9) | 24851.9 ( 7.9) | 0.057 |

**Table A9**. Primary analysis: users (≥8 prescriptions) vs. non-users (counts/weighted counts, %)

|  | Before matching | | | After matching | | |
| --- | --- | --- | --- | --- | --- | --- |
| Characteristics | Treated | Control | d | Treated | Control | d |
| N | 18372 | 332389 |  | 15150 | 313399 |  |
| Age (5-year bins) |  |  |  |  |  |  |
| [16,21) | 22 ( 0.1) | 22054 ( 6.6) | -0.367 | 922.9 ( 6.1) | 19092.1 ( 6.1) | 0.000 |
| [21,26) | 99 ( 0.5) | 27711 ( 8.3) | -0.386 | 1190.7 ( 7.9) | 24632.3 ( 7.9) | 0.000 |
| [26,31) | 160 ( 0.9) | 31607 ( 9.5) | -0.397 | 1392.7 ( 9.2) | 28809.3 ( 9.2) | 0.000 |
| [31,36) | 259 ( 1.4) | 34524 (10.4) | -0.388 | 1539.3 (10.2) | 31842.7 (10.2) | 0.000 |
| [36,41) | 486 ( 2.6) | 37047 (11.1) | -0.340 | 1692.1 (11.2) | 35002.9 (11.2) | 0.000 |
| [41,46) | 733 ( 4.0) | 35359 (10.6) | -0.257 | 1615.9 (10.7) | 33426.1 (10.7) | 0.000 |
| [46,51) | 1066 ( 5.8) | 30711 ( 9.2) | -0.131 | 1422.2 ( 9.4) | 29419.8 ( 9.4) | 0.000 |
| [51,56) | 1473 ( 8.0) | 28177 ( 8.5) | -0.017 | 1304.6 ( 8.6) | 26987.4 ( 8.6) | 0.000 |
| [56,61) | 1912 (10.4) | 25009 ( 7.5) | 0.101 | 1171.1 ( 7.7) | 24226.9 ( 7.7) | 0.000 |
| [61,66) | 2301 (12.5) | 19809 ( 6.0) | 0.228 | 955.0 ( 6.3) | 19756.0 ( 6.3) | 0.000 |
| [66,71) | 2346 (12.8) | 14082 ( 4.2) | 0.310 | 679.8 ( 4.5) | 14063.2 ( 4.5) | 0.000 |
| [71,76) | 2031 (11.1) | 9476 ( 2.9) | 0.327 | 450.4 ( 3.0) | 9316.6 ( 3.0) | 0.000 |
| [76,81) | 1999 (10.9) | 6813 ( 2.0) | 0.365 | 340.0 ( 2.2) | 7034.0 ( 2.2) | 0.000 |
| [81,86) | 1919 (10.4) | 5529 ( 1.7) | 0.375 | 272.9 ( 1.8) | 5645.1 ( 1.8) | 0.000 |
| [86,91) | 1197 ( 6.5) | 3158 ( 1.0) | 0.297 | 152.8 ( 1.0) | 3161.2 ( 1.0) | 0.000 |
| [91,96) | 328 ( 1.8) | 1101 ( 0.3) | 0.142 | 42.6 ( 0.3) | 880.4 ( 0.3) | 0.000 |
| [96,101) | 39 ( 0.2) | 214 ( 0.1) | 0.040 | 5.0 ( 0.0) | 103.0 ( 0.0) | 0.000 |
| [101,106) | 2 ( 0.0) | 7 ( 0.0) | 0.011 | 0.0 ( 0.0) | 0.0 ( 0.0) | 0.000 |
| [106,111) | 0 ( 0.0) | 1 ( 0.0) | -0.002 | 0.0 ( 0.0) | 0.0 ( 0.0) | 0.000 |
| Sex |  |  |  |  |  |  |
| M | 7466 (40.6) | 162983 (49.0) | -0.169 | 7301.8 (48.2) | 151048.2 (48.2) | 0.000 |
| F | 10906 (59.4) | 169406 (51.0) | 0.169 | 7848.2 (51.8) | 162350.8 (51.8) | 0.000 |
| Vaccination status |  |  |  |  |  |  |
| 1/1, >90d | 0 ( 0.0) | 2 ( 0.0) | -0.003 | 0.0 ( 0.0) | 0.0 ( 0.0) | 0.000 |
| 1/1, <14d | 8 ( 0.0) | 47 ( 0.0) | 0.017 | 0.0 ( 0.0) | 0.0 ( 0.0) | 0.000 |
| 1/1, 14-90d | 1 ( 0.0) | 48 ( 0.0) | -0.009 | 0.0 ( 0.0) | 0.0 ( 0.0) | 0.000 |
| Not vaccinated | 16741 (91.1) | 322899 (97.1) | -0.258 | 14931.3 (98.6) | 308875.7 (98.6) | 0.000 |
| 1/2, >90d | 7 ( 0.0) | 35 ( 0.0) | 0.018 | 0.0 ( 0.0) | 0.0 ( 0.0) | 0.000 |
| 1/2, <14d | 422 ( 2.3) | 3549 ( 1.1) | 0.096 | 93.7 ( 0.6) | 1939.3 ( 0.6) | 0.000 |
| 1/2, 14-90d | 739 ( 4.0) | 3880 ( 1.2) | 0.180 | 112.4 ( 0.7) | 2325.6 ( 0.7) | 0.000 |
| 2/2, >90d | 127 ( 0.7) | 363 ( 0.1) | 0.092 | 1.7 ( 0.0) | 34.3 ( 0.0) | 0.000 |
| 2/2, <14d | 97 ( 0.5) | 420 ( 0.1) | 0.070 | 3.5 ( 0.0) | 72.5 ( 0.0) | 0.000 |
| 2/2, 14-90d | 230 ( 1.3) | 1146 ( 0.3) | 0.102 | 7.3 ( 0.0) | 151.7 ( 0.0) | 0.000 |
| Period |  |  |  |  |  |  |
| before 10.01.2021. | 11189 (60.9) | 192599 (57.9) | 0.060 | 9110.5 (60.1) | 188463.5 (60.1) | 0.000 |
| 10.01.-15.07.2021. | 6982 (38.0) | 132866 (40.0) | -0.040 | 5955.5 (39.3) | 123197.5 (39.3) | 0.000 |
| after 15.07.2021. | 201 ( 1.1) | 6924 ( 2.1) | -0.079 | 84.0 ( 0.6) | 1738.0 ( 0.6) | 0.000 |
| CC index |  |  |  |  |  |  |
| 0 | 5613 (30.6) | 270536 (81.4) | -1.192 | 12354.2 (81.5) | 255564.8 (81.5) | 0.000 |
| 1-2 | 8063 (43.9) | 53673 (16.1) | 0.635 | 2480.7 (16.4) | 51317.3 (16.4) | 0.000 |
| 3-4 | 3308 (18.0) | 6652 ( 2.0) | 0.553 | 276.3 ( 1.8) | 5715.7 ( 1.8) | 0.000 |
| >=5 | 1388 ( 7.6) | 1528 ( 0.5) | 0.368 | 38.7 ( 0.3) | 801.3 ( 0.3) | 0.000 |
| Comorbidities |  |  |  |  |  |  |
| Atrial fibrillation | 2670 (14.5) | 6300 ( 1.9) | 0.473 | 240.2 ( 1.6) | 4969.8 ( 1.6) | 0.000 |
| Autoimmune | 3627 (19.7) | 19580 ( 5.9) | 0.424 | 853.0 ( 5.6) | 17645.0 ( 5.6) | 0.000 |
| Cancer | 2340 (12.7) | 10142 ( 3.1) | 0.365 | 382.2 ( 2.5) | 7905.8 ( 2.5) | 0.000 |
| Congestive heart failure | 2098 (11.4) | 3380 ( 1.0) | 0.441 | 120.5 ( 0.8) | 2492.5 ( 0.8) | 0.000 |
| COPD | 3922 (21.3) | 21520 ( 6.5) | 0.440 | 943.4 ( 6.2) | 19516.6 ( 6.2) | 0.000 |
| IHD or CVD | 5385 (29.3) | 12849 ( 3.9) | 0.728 | 617.3 ( 4.1) | 12769.7 ( 4.1) | 0.000 |
| Renal disease | 1307 ( 7.1) | 1710 ( 0.5) | 0.350 | 42.8 ( 0.3) | 886.2 ( 0.3) | 0.000 |
| Immunocompromised | 987 ( 5.4) | 1463 ( 0.4) | 0.297 | 42.9 ( 0.3) | 888.1 ( 0.3) | 0.000 |
| RAAS inhibitors | 10065 (54.8) | 43193 (13.0) | 0.984 | 2173.2 (14.3) | 44955.8 (14.3) | 0.000 |
| Death (composite) | 1889 (10.3) | 5599 ( 1.7) | 0.369 | 318.9 ( 2.1) | 5380.0 ( 1.7) | 0.017 |
| Death 1 | 1412 ( 7.7) | 4366 ( 1.3) | 0.311 | 244.9 ( 1.6) | 4232.1 ( 1.4) | 0.013 |
| Death 2 | 1215 ( 6.6) | 3698 ( 1.1) | 0.288 | 206.5 ( 1.4) | 3529.6 ( 1.1) | 0.012 |
| Death 3 | 1600 ( 8.7) | 4519 ( 1.4) | 0.341 | 265.8 ( 1.8) | 4348.9 ( 1.4) | 0.017 |
| Hospitalization 1 | 2348 (12.8) | 10905 ( 3.3) | 0.355 | 781.9 ( 5.2) | 9815.0 ( 3.1) | 0.076 |
| Hospitalization 2 | 5015 (27.3) | 26965 ( 8.1) | 0.519 | 1646.2 (10.9) | 25255.3 ( 8.1) | 0.076 |

**Table A10**. Primary analysis: users (all) vs. possible users (counts/weighted counts, %)

|  | Before matching | | | After matching | | |
| --- | --- | --- | --- | --- | --- | --- |
| Characteristics | Treated | Control | d | Treated | Control | d |
| N | 55098 | 18170 |  | 41195 | 17344 |  |
| Age (5-year bins) |  |  |  |  |  |  |
| [16,21) | 401 ( 0.7) | 540 ( 3.0) | -0.167 | 639.7 ( 1.6) | 269.3 ( 1.6) | 0.000 |
| [21,26) | 782 ( 1.4) | 759 ( 4.2) | -0.168 | 1037.3 ( 2.5) | 436.7 ( 2.5) | 0.000 |
| [26,31) | 1137 ( 2.1) | 981 ( 5.4) | -0.177 | 1400.4 ( 3.4) | 589.6 ( 3.4) | 0.000 |
| [31,36) | 1703 ( 3.1) | 1142 ( 6.3) | -0.152 | 1896.5 ( 4.6) | 798.5 ( 4.6) | 0.000 |
| [36,41) | 2571 ( 4.7) | 1425 ( 7.8) | -0.131 | 2622.8 ( 6.4) | 1104.2 ( 6.4) | 0.000 |
| [41,46) | 3335 ( 6.1) | 1644 ( 9.0) | -0.114 | 3264.6 ( 7.9) | 1374.4 ( 7.9) | 0.000 |
| [46,51) | 4243 ( 7.7) | 1775 ( 9.8) | -0.073 | 3836.0 ( 9.3) | 1615.0 ( 9.3) | 0.000 |
| [51,56) | 5301 ( 9.6) | 1833 (10.1) | -0.016 | 4481.3 (10.9) | 1886.7 (10.9) | 0.000 |
| [56,61) | 6107 (11.1) | 1886 (10.4) | 0.023 | 4904.2 (11.9) | 2064.8 (11.9) | 0.000 |
| [61,66) | 6560 (11.9) | 1664 ( 9.2) | 0.090 | 4726.2 (11.5) | 1989.8 (11.5) | 0.000 |
| [66,71) | 6134 (11.1) | 1332 ( 7.3) | 0.132 | 3810.6 ( 9.3) | 1604.4 ( 9.3) | 0.000 |
| [71,76) | 5050 ( 9.2) | 1010 ( 5.6) | 0.138 | 2819.8 ( 6.8) | 1187.2 ( 6.8) | 0.000 |
| [76,81) | 4480 ( 8.1) | 831 ( 4.6) | 0.146 | 2290.6 ( 5.6) | 964.4 ( 5.6) | 0.000 |
| [81,86) | 4104 ( 7.4) | 755 ( 4.2) | 0.141 | 2054.2 ( 5.0) | 864.8 ( 5.0) | 0.000 |
| [86,91) | 2400 ( 4.4) | 430 ( 2.4) | 0.111 | 1134.4 ( 2.8) | 477.6 ( 2.8) | 0.000 |
| [91,96) | 693 ( 1.3) | 135 ( 0.7) | 0.052 | 247.7 ( 0.6) | 104.3 ( 0.6) | 0.000 |
| [96,101) | 95 ( 0.2) | 28 ( 0.2) | 0.005 | 28.9 ( 0.1) | 12.1 ( 0.1) | 0.000 |
| [101,106) | 2 ( 0.0) | 0 ( 0.0) | 0.009 | 0.0 ( 0.0) | 0.0 ( 0.0) | 0.000 |
| [106,111) | 0 ( 0.0) | 0 ( 0.0) | 0.000 | 0.0 ( 0.0) | 0.0 ( 0.0) | 0.000 |
| Sex |  |  |  |  |  |  |
| M | 22645 (41.1) | 7582 (41.7) | -0.013 | 16425.5 (39.9) | 6915.5 (39.9) | 0.000 |
| F | 32453 (58.9) | 10588 (58.3) | 0.013 | 24769.5 (60.1) | 10428.5 (60.1) | 0.000 |
| Vaccination status |  |  |  |  |  |  |
| 1/1, >90d | 2 ( 0.0) | 0 ( 0.0) | 0.009 | 0.0 ( 0.0) | 0.0 ( 0.0) | 0.000 |
| 1/1, <14d | 12 ( 0.0) | 1 ( 0.0) | 0.014 | 0.0 ( 0.0) | 0.0 ( 0.0) | 0.000 |
| 1/1, 14-90d | 5 ( 0.0) | 0 ( 0.0) | 0.013 | 0.0 ( 0.0) | 0.0 ( 0.0) | 0.000 |
| Not vaccinated | 51111 (92.8) | 17347 (95.5) | -0.115 | 40128.9 (97.4) | 16895.1 (97.4) | 0.000 |
| 1/2, >90d | 22 ( 0.0) | 7 ( 0.0) | 0.001 | 0.0 ( 0.0) | 0.0 ( 0.0) | 0.000 |
| 1/2, <14d | 1121 ( 2.0) | 295 ( 1.6) | 0.031 | 429.3 ( 1.0) | 180.7 ( 1.0) | 0.000 |
| 1/2, 14-90d | 1791 ( 3.3) | 355 ( 2.0) | 0.082 | 570.7 ( 1.4) | 240.3 ( 1.4) | 0.000 |
| 2/2, >90d | 239 ( 0.4) | 30 ( 0.2) | 0.049 | 4.2 ( 0.0) | 1.8 ( 0.0) | 0.000 |
| 2/2, <14d | 242 ( 0.4) | 44 ( 0.2) | 0.034 | 18.3 ( 0.0) | 7.7 ( 0.0) | 0.000 |
| 2/2, 14-90d | 553 ( 1.0) | 91 ( 0.5) | 0.058 | 43.6 ( 0.1) | 18.4 ( 0.1) | 0.000 |
| Period |  |  |  |  |  |  |
| before 10.01.2021. | 32317 (58.7) | 10566 (58.2) | 0.010 | 25364.8 (61.6) | 10679.2 (61.6) | 0.000 |
| 10.01.-15.07.2021. | 22102 (40.1) | 7257 (39.9) | 0.004 | 15540.9 (37.7) | 6543.1 (37.7) | 0.000 |
| after 15.07.2021. | 679 ( 1.2) | 347 ( 1.9) | -0.054 | 289.2 ( 0.7) | 121.8 ( 0.7) | 0.000 |
| CC index |  |  |  |  |  |  |
| 0 | 23477 (42.6) | 11868 (65.3) | -0.468 | 23950.4 (58.1) | 10083.6 (58.1) | 0.000 |
| 1-2 | 21575 (39.2) | 5248 (28.9) | 0.218 | 15047.6 (36.5) | 6335.4 (36.5) | 0.000 |
| 3-4 | 7228 (13.1) | 845 ( 4.7) | 0.301 | 1962.0 ( 4.8) | 826.0 ( 4.8) | 0.000 |
| >=5 | 2818 ( 5.1) | 209 ( 1.2) | 0.229 | 235.0 ( 0.6) | 99.0 ( 0.6) | 0.000 |
| Comorbidities |  |  |  |  |  |  |
| Atrial fibrillation | 5687 (10.3) | 661 ( 3.6) | 0.265 | 1465.1 ( 3.6) | 616.9 ( 3.6) | 0.000 |
| Autoimmune | 10034 (18.2) | 2270 (12.5) | 0.159 | 5704.3 (13.8) | 2401.7 (13.8) | 0.000 |
| Cancer | 6253 (11.3) | 978 ( 5.4) | 0.217 | 2348.3 ( 5.7) | 988.7 ( 5.7) | 0.000 |
| Congestive heart failure | 4234 ( 7.7) | 462 ( 2.5) | 0.235 | 764.9 ( 1.9) | 322.1 ( 1.9) | 0.000 |
| COPD | 9700 (17.6) | 1979 (10.9) | 0.193 | 4953.5 (12.0) | 2085.5 (12.0) | 0.000 |
| IHD or CVD | 11683 (21.2) | 1518 ( 8.4) | 0.368 | 4733.2 (11.5) | 1992.8 (11.5) | 0.000 |
| Renal disease | 2580 ( 4.7) | 230 ( 1.3) | 0.202 | 225.9 ( 0.5) | 95.1 ( 0.5) | 0.000 |
| Immunocompromised | 1873 ( 3.4) | 132 ( 0.7) | 0.189 | 194.9 ( 0.5) | 82.1 ( 0.5) | 0.000 |
| RAAS inhibitors | 23376 (42.4) | 4841 (26.6) | 0.337 | 14109.6 (34.3) | 5940.4 (34.3) | 0.000 |
| Death (composite) | 4356 ( 7.9) | 803 ( 4.4) | 0.145 | 1904.4 ( 4.6) | 860.4 ( 5.0) | -0.014 |
| Death 1 | 3195 ( 5.8) | 585 ( 3.2) | 0.125 | 1446.9 ( 3.5) | 633.7 ( 3.7) | -0.007 |
| Death 2 | 2839 ( 5.2) | 519 ( 2.9) | 0.117 | 1233.5 ( 3.0) | 552.2 ( 3.2) | -0.010 |
| Death 3 | 3670 ( 6.7) | 666 ( 3.7) | 0.136 | 1566.0 ( 3.8) | 705.8 ( 4.1) | -0.012 |
| Hospitalization 1 | 5824 (10.6) | 1160 ( 6.4) | 0.151 | 2900.0 ( 7.0) | 1169.8 ( 6.7) | 0.011 |
| Hospitalization 2 | 12735 (23.1) | 2680 (14.7) | 0.215 | 6913.2 (16.8) | 2799.2 (16.1) | 0.016 |

**Table A11**. Primary analysis: users (1-3 prescriptions) vs. possible users (counts/weighted counts, %)

|  | Before matching | | | After matching | | |
| --- | --- | --- | --- | --- | --- | --- |
| Characteristics | Treated | Control | d | Treated | Control | d |
| N | 18996 | 18170 |  | 15998 | 16517 |  |
| Age (5-year bins) |  |  |  |  |  |  |
| [16,21) | 305 ( 1.6) | 540 ( 3.0) | -0.091 | 404.4 ( 2.5) | 417.6 ( 2.5) | 0.000 |
| [21,26) | 515 ( 2.7) | 759 ( 4.2) | -0.080 | 604.2 ( 3.8) | 623.8 ( 3.8) | 0.000 |
| [26,31) | 726 ( 3.8) | 981 ( 5.4) | -0.075 | 802.0 ( 5.0) | 828.0 ( 5.0) | 0.000 |
| [31,36) | 1002 ( 5.3) | 1142 ( 6.3) | -0.043 | 1020.4 ( 6.4) | 1053.6 ( 6.4) | 0.000 |
| [36,41) | 1378 ( 7.3) | 1425 ( 7.8) | -0.022 | 1316.6 ( 8.2) | 1359.4 ( 8.2) | 0.000 |
| [41,46) | 1596 ( 8.4) | 1644 ( 9.0) | -0.023 | 1523.3 ( 9.5) | 1572.7 ( 9.5) | 0.000 |
| [46,51) | 1825 ( 9.6) | 1775 ( 9.8) | -0.005 | 1672.4 (10.5) | 1726.6 (10.5) | 0.000 |
| [51,56) | 2101 (11.1) | 1833 (10.1) | 0.032 | 1791.9 (11.2) | 1850.1 (11.2) | 0.000 |
| [56,61) | 2099 (11.0) | 1886 (10.4) | 0.022 | 1795.4 (11.2) | 1853.6 (11.2) | 0.000 |
| [61,66) | 2054 (10.8) | 1664 ( 9.2) | 0.055 | 1599.1 (10.0) | 1650.9 (10.0) | 0.000 |
| [66,71) | 1680 ( 8.8) | 1332 ( 7.3) | 0.056 | 1201.0 ( 7.5) | 1240.0 ( 7.5) | 0.000 |
| [71,76) | 1310 ( 6.9) | 1010 ( 5.6) | 0.055 | 836.4 ( 5.2) | 863.6 ( 5.2) | 0.000 |
| [76,81) | 1012 ( 5.3) | 831 ( 4.6) | 0.035 | 599.8 ( 3.7) | 619.2 ( 3.7) | 0.000 |
| [81,86) | 826 ( 4.3) | 755 ( 4.2) | 0.010 | 514.2 ( 3.2) | 530.8 ( 3.2) | 0.000 |
| [86,91) | 433 ( 2.3) | 430 ( 2.4) | -0.006 | 258.8 ( 1.6) | 267.2 ( 1.6) | 0.000 |
| [91,96) | 118 ( 0.6) | 135 ( 0.7) | -0.015 | 54.6 ( 0.3) | 56.4 ( 0.3) | 0.000 |
| [96,101) | 16 ( 0.1) | 28 ( 0.2) | -0.020 | 3.4 ( 0.0) | 3.6 ( 0.0) | 0.000 |
| [101,106) | 0 ( 0.0) | 0 ( 0.0) | 0.000 | 0.0 ( 0.0) | 0.0 ( 0.0) | 0.000 |
| [106,111) | 0 ( 0.0) | 0 ( 0.0) | 0.000 | 0.0 ( 0.0) | 0.0 ( 0.0) | 0.000 |
| Sex |  |  |  |  |  |  |
| M | 7707 (40.6) | 7582 (41.7) | -0.024 | 6375.6 (39.9) | 6582.4 (39.9) | 0.000 |
| F | 11289 (59.4) | 10588 (58.3) | 0.024 | 9622.4 (60.1) | 9934.6 (60.1) | 0.000 |
| Vaccination status |  |  |  |  |  |  |
| 1/1, >90d | 1 ( 0.0) | 0 ( 0.0) | 0.010 | 0.0 ( 0.0) | 0.0 ( 0.0) | 0.000 |
| 1/1, <14d | 2 ( 0.0) | 1 ( 0.0) | 0.006 | 0.0 ( 0.0) | 0.0 ( 0.0) | 0.000 |
| 1/1, 14-90d | 2 ( 0.0) | 0 ( 0.0) | 0.015 | 0.0 ( 0.0) | 0.0 ( 0.0) | 0.000 |
| Not vaccinated | 18000 (94.8) | 17347 (95.5) | -0.033 | 15649.7 (97.8) | 16157.3 (97.8) | 0.000 |
| 1/2, >90d | 7 ( 0.0) | 7 ( 0.0) | -0.001 | 0.0 ( 0.0) | 0.0 ( 0.0) | 0.000 |
| 1/2, <14d | 332 ( 1.7) | 295 ( 1.6) | 0.010 | 155.5 ( 1.0) | 160.5 ( 1.0) | 0.000 |
| 1/2, 14-90d | 431 ( 2.3) | 355 ( 2.0) | 0.022 | 171.2 ( 1.1) | 176.8 ( 1.1) | 0.000 |
| 2/2, >90d | 36 ( 0.2) | 30 ( 0.2) | 0.006 | 0.0 ( 0.0) | 0.0 ( 0.0) | 0.000 |
| 2/2, <14d | 56 ( 0.3) | 44 ( 0.2) | 0.010 | 4.9 ( 0.0) | 5.1 ( 0.0) | 0.000 |
| 2/2, 14-90d | 129 ( 0.7) | 91 ( 0.5) | 0.023 | 16.7 ( 0.1) | 17.3 ( 0.1) | 0.000 |
| Period |  |  |  |  |  |  |
| before 10.01.2021. | 10627 (55.9) | 10566 (58.2) | -0.045 | 9477.8 (59.2) | 9785.2 (59.2) | 0.000 |
| 10.01.-15.07.2021. | 8105 (42.7) | 7257 (39.9) | 0.055 | 6362.8 (39.8) | 6569.2 (39.8) | 0.000 |
| after 15.07.2021. | 264 ( 1.4) | 347 ( 1.9) | -0.041 | 157.4 ( 1.0) | 162.6 ( 1.0) | 0.000 |
| CC index |  |  |  |  |  |  |
| 0 | 10660 (56.1) | 11868 (65.3) | -0.189 | 10760.5 (67.3) | 11109.5 (67.3) | 0.000 |
| 1-2 | 6259 (32.9) | 5248 (28.9) | 0.088 | 4724.9 (29.5) | 4878.1 (29.5) | 0.000 |
| 3-4 | 1527 ( 8.0) | 845 ( 4.7) | 0.139 | 454.6 ( 2.8) | 469.4 ( 2.8) | 0.000 |
| >=5 | 550 ( 2.9) | 209 ( 1.2) | 0.124 | 58.1 ( 0.4) | 59.9 ( 0.4) | 0.000 |
| Comorbidities |  |  |  |  |  |  |
| Atrial fibrillation | 1163 ( 6.1) | 661 ( 3.6) | 0.116 | 313.4 ( 2.0) | 323.6 ( 2.0) | 0.000 |
| Autoimmune | 2987 (15.7) | 2270 (12.5) | 0.093 | 1935.6 (12.1) | 1998.4 (12.1) | 0.000 |
| Cancer | 1873 ( 9.9) | 978 ( 5.4) | 0.169 | 768.0 ( 4.8) | 793.0 ( 4.8) | 0.000 |
| Congestive heart failure | 783 ( 4.1) | 462 ( 2.5) | 0.088 | 158.4 ( 1.0) | 163.6 ( 1.0) | 0.000 |
| COPD | 2626 (13.8) | 1979 (10.9) | 0.089 | 1603.0 (10.0) | 1655.0 (10.0) | 0.000 |
| IHD or CVD | 2437 (12.8) | 1518 ( 8.4) | 0.146 | 1112.5 ( 7.0) | 1148.5 ( 7.0) | 0.000 |
| Renal disease | 409 ( 2.2) | 230 ( 1.3) | 0.068 | 36.4 ( 0.2) | 37.6 ( 0.2) | 0.000 |
| Immunocompromised | 266 ( 1.4) | 132 ( 0.7) | 0.066 | 33.5 ( 0.2) | 34.5 ( 0.2) | 0.000 |
| RAAS inhibitors | 5450 (28.7) | 4841 (26.6) | 0.046 | 3998.1 (25.0) | 4127.9 (25.0) | 0.000 |
| Death (composite) | 1070 ( 5.6) | 803 ( 4.4) | 0.056 | 566.1 ( 3.5) | 561.3 ( 3.4) | 0.006 |
| Death 1 | 761 ( 4.0) | 585 ( 3.2) | 0.042 | 430.1 ( 2.7) | 423.1 ( 2.6) | 0.007 |
| Death 2 | 704 ( 3.7) | 519 ( 2.9) | 0.048 | 370.4 ( 2.3) | 375.6 ( 2.3) | 0.002 |
| Death 3 | 900 ( 4.7) | 666 ( 3.7) | 0.053 | 466.1 ( 2.9) | 456.5 ( 2.8) | 0.007 |
| Hospitalization 1 | 1646 ( 8.7) | 1160 ( 6.4) | 0.087 | 1027.7 ( 6.4) | 884.3 ( 5.4) | 0.041 |
| Hospitalization 2 | 3624 (19.1) | 2680 (14.7) | 0.116 | 2383.4 (14.9) | 2167.9 (13.1) | 0.047 |

**Table A12**. Primary analysis: users (4-7 prescriptions) vs. possible users (counts/weighted counts, %)

|  | Before matching | | | After matching | | |
| --- | --- | --- | --- | --- | --- | --- |
| Characteristics | Treated | Control | d | Treated | Control | d |
| N | 17730 | 18170 |  | 13115 | 16594 |  |
| Age (5-year bins) |  |  |  |  |  |  |
| [16,21) | 74 ( 0.4) | 540 ( 3.0) | -0.199 | 248.5 ( 1.9) | 314.5 ( 1.9) | 0.000 |
| [21,26) | 168 ( 0.9) | 759 ( 4.2) | -0.205 | 385.4 ( 2.9) | 487.6 ( 2.9) | 0.000 |
| [26,31) | 251 ( 1.4) | 981 ( 5.4) | -0.221 | 514.3 ( 3.9) | 650.7 ( 3.9) | 0.000 |
| [31,36) | 442 ( 2.5) | 1142 ( 6.3) | -0.186 | 661.3 ( 5.0) | 836.7 ( 5.0) | 0.000 |
| [36,41) | 707 ( 4.0) | 1425 ( 7.8) | -0.164 | 885.5 ( 6.8) | 1120.5 ( 6.8) | 0.000 |
| [41,46) | 1006 ( 5.7) | 1644 ( 9.0) | -0.129 | 1092.6 ( 8.3) | 1382.4 ( 8.3) | 0.000 |
| [46,51) | 1352 ( 7.6) | 1775 ( 9.8) | -0.076 | 1257.7 ( 9.6) | 1591.3 ( 9.6) | 0.000 |
| [51,56) | 1727 ( 9.7) | 1833 (10.1) | -0.012 | 1419.7 (10.8) | 1796.3 (10.8) | 0.000 |
| [56,61) | 2096 (11.8) | 1886 (10.4) | 0.046 | 1548.6 (11.8) | 1959.4 (11.8) | 0.000 |
| [61,66) | 2205 (12.4) | 1664 ( 9.2) | 0.106 | 1435.6 (10.9) | 1816.4 (10.9) | 0.000 |
| [66,71) | 2108 (11.9) | 1332 ( 7.3) | 0.155 | 1165.0 ( 8.9) | 1474.0 ( 8.9) | 0.000 |
| [71,76) | 1709 ( 9.6) | 1010 ( 5.6) | 0.154 | 848.5 ( 6.5) | 1073.5 ( 6.5) | 0.000 |
| [76,81) | 1469 ( 8.3) | 831 ( 4.6) | 0.152 | 665.3 ( 5.1) | 841.7 ( 5.1) | 0.000 |
| [81,86) | 1359 ( 7.7) | 755 ( 4.2) | 0.149 | 595.5 ( 4.5) | 753.5 ( 4.5) | 0.000 |
| [86,91) | 770 ( 4.3) | 430 ( 2.4) | 0.110 | 311.2 ( 2.4) | 393.8 ( 2.4) | 0.000 |
| [91,96) | 247 ( 1.4) | 135 ( 0.7) | 0.063 | 68.0 ( 0.5) | 86.0 ( 0.5) | 0.000 |
| [96,101) | 40 ( 0.2) | 28 ( 0.2) | 0.016 | 12.4 ( 0.1) | 15.6 ( 0.1) | 0.000 |
| [101,106) | 0 ( 0.0) | 0 ( 0.0) | 0.000 | 0.0 ( 0.0) | 0.0 ( 0.0) | 0.000 |
| [106,111) | 0 ( 0.0) | 0 ( 0.0) | 0.000 | 0.0 ( 0.0) | 0.0 ( 0.0) | 0.000 |
| Sex |  |  |  |  |  |  |
| M | 7472 (42.1) | 7582 (41.7) | 0.008 | 5331.8 (40.7) | 6746.2 (40.7) | 0.000 |
| F | 10258 (57.9) | 10588 (58.3) | -0.008 | 7783.2 (59.3) | 9847.8 (59.3) | 0.000 |
| Vaccination status |  |  |  |  |  |  |
| 1/1, >90d | 1 ( 0.0) | 0 ( 0.0) | 0.011 | 0.0 ( 0.0) | 0.0 ( 0.0) | 0.000 |
| 1/1, <14d | 2 ( 0.0) | 1 ( 0.0) | 0.006 | 0.0 ( 0.0) | 0.0 ( 0.0) | 0.000 |
| 1/1, 14-90d | 2 ( 0.0) | 0 ( 0.0) | 0.015 | 0.0 ( 0.0) | 0.0 ( 0.0) | 0.000 |
| Not vaccinated | 16370 (92.3) | 17347 (95.5) | -0.132 | 12798.5 (97.6) | 16193.5 (97.6) | 0.000 |
| 1/2, >90d | 8 ( 0.0) | 7 ( 0.0) | 0.003 | 0.0 ( 0.0) | 0.0 ( 0.0) | 0.000 |
| 1/2, <14d | 367 ( 2.1) | 295 ( 1.6) | 0.033 | 124.5 ( 0.9) | 157.5 ( 0.9) | 0.000 |
| 1/2, 14-90d | 621 ( 3.5) | 355 ( 2.0) | 0.095 | 173.5 ( 1.3) | 219.5 ( 1.3) | 0.000 |
| 2/2, >90d | 76 ( 0.4) | 30 ( 0.2) | 0.048 | 2.2 ( 0.0) | 2.8 ( 0.0) | 0.000 |
| 2/2, <14d | 89 ( 0.5) | 44 ( 0.2) | 0.043 | 5.7 ( 0.0) | 7.3 ( 0.0) | 0.000 |
| 2/2, 14-90d | 194 ( 1.1) | 91 ( 0.5) | 0.067 | 10.6 ( 0.1) | 13.4 ( 0.1) | 0.000 |
| Period |  |  |  |  |  |  |
| before 10.01.2021. | 10501 (59.2) | 10566 (58.2) | 0.022 | 8111.2 (61.8) | 10262.8 (61.8) | 0.000 |
| 10.01.-15.07.2021. | 7015 (39.6) | 7257 (39.9) | -0.008 | 4915.1 (37.5) | 6218.9 (37.5) | 0.000 |
| after 15.07.2021. | 214 ( 1.2) | 347 ( 1.9) | -0.057 | 88.7 ( 0.7) | 112.3 ( 0.7) | 0.000 |
| CC index |  |  |  |  |  |  |
| 0 | 7204 (40.6) | 11868 (65.3) | -0.510 | 8116.9 (61.9) | 10270.1 (61.9) | 0.000 |
| 1-2 | 7253 (40.9) | 5248 (28.9) | 0.254 | 4430.8 (33.8) | 5606.2 (33.8) | 0.000 |
| 3-4 | 2393 (13.5) | 845 ( 4.7) | 0.312 | 508.1 ( 3.9) | 642.9 ( 3.9) | 0.000 |
| >=5 | 880 ( 5.0) | 209 ( 1.2) | 0.223 | 59.2 ( 0.5) | 74.8 ( 0.5) | 0.000 |
| Comorbidities |  |  |  |  |  |  |
| Atrial fibrillation | 1854 (10.5) | 661 ( 3.6) | 0.269 | 386.7 ( 2.9) | 489.3 ( 2.9) | 0.000 |
| Autoimmune | 3420 (19.3) | 2270 (12.5) | 0.187 | 1715.5 (13.1) | 2170.5 (13.1) | 0.000 |
| Cancer | 2040 (11.5) | 978 ( 5.4) | 0.222 | 637.5 ( 4.9) | 806.5 ( 4.9) | 0.000 |
| Congestive heart failure | 1353 ( 7.6) | 462 ( 2.5) | 0.233 | 194.2 ( 1.5) | 245.8 ( 1.5) | 0.000 |
| COPD | 3152 (17.8) | 1979 (10.9) | 0.197 | 1447.5 (11.0) | 1831.5 (11.0) | 0.000 |
| IHD or CVD | 3861 (21.8) | 1518 ( 8.4) | 0.382 | 1265.6 ( 9.7) | 1601.4 ( 9.7) | 0.000 |
| Renal disease | 864 ( 4.9) | 230 ( 1.3) | 0.210 | 60.9 ( 0.5) | 77.1 ( 0.5) | 0.000 |
| Immunocompromised | 620 ( 3.5) | 132 ( 0.7) | 0.194 | 48.1 ( 0.4) | 60.9 ( 0.4) | 0.000 |
| RAAS inhibitors | 7861 (44.3) | 4841 (26.6) | 0.376 | 4181.8 (31.9) | 5291.2 (31.9) | 0.000 |
| Death (composite) | 1397 ( 7.9) | 803 ( 4.4) | 0.144 | 516.4 ( 3.9) | 744.4 ( 4.5) | -0.023 |
| Death 1 | 1022 ( 5.8) | 585 ( 3.2) | 0.123 | 387.8 ( 3.0) | 552.0 ( 3.3) | -0.018 |
| Death 2 | 920 ( 5.2) | 519 ( 2.9) | 0.119 | 331.8 ( 2.5) | 477.8 ( 2.9) | -0.018 |
| Death 3 | 1170 ( 6.6) | 666 ( 3.7) | 0.133 | 423.4 ( 3.2) | 609.1 ( 3.7) | -0.020 |
| Hospitalization 1 | 1830 (10.3) | 1160 ( 6.4) | 0.143 | 801.4 ( 6.1) | 1047.5 ( 6.3) | -0.007 |
| Hospitalization 2 | 4096 (23.1) | 2680 (14.7) | 0.214 | 1987.0 (15.2) | 2536.9 (15.3) | -0.004 |

**Table A13**. Primary analysis: users (≥8 prescriptions) vs. possible users (counts/weighted counts, %)

|  | Before matching | | | After matching | | |
| --- | --- | --- | --- | --- | --- | --- |
| Characteristics | Treated | Control | d | Treated | Control | d |
| N | 18372 | 18170 |  | 12082 | 16564 |  |
| Age (5-year bins) |  |  |  |  |  |  |
| [16,21) | 22 ( 0.1) | 540 ( 3.0) | -0.233 | 205.8 ( 1.7) | 282.2 ( 1.7) | 0.000 |
| [21,26) | 99 ( 0.5) | 759 ( 4.2) | -0.242 | 326.0 ( 2.7) | 447.0 ( 2.7) | 0.000 |
| [26,31) | 160 ( 0.9) | 981 ( 5.4) | -0.262 | 442.4 ( 3.7) | 606.6 ( 3.7) | 0.000 |
| [31,36) | 259 ( 1.4) | 1142 ( 6.3) | -0.256 | 539.0 ( 4.5) | 739.0 ( 4.5) | 0.000 |
| [36,41) | 486 ( 2.6) | 1425 ( 7.8) | -0.235 | 757.9 ( 6.3) | 1039.1 ( 6.3) | 0.000 |
| [41,46) | 733 ( 4.0) | 1644 ( 9.0) | -0.206 | 924.5 ( 7.7) | 1267.5 ( 7.7) | 0.000 |
| [46,51) | 1066 ( 5.8) | 1775 ( 9.8) | -0.148 | 1079.3 ( 8.9) | 1479.7 ( 8.9) | 0.000 |
| [51,56) | 1473 ( 8.0) | 1833 (10.1) | -0.072 | 1219.3 (10.1) | 1671.7 (10.1) | 0.000 |
| [56,61) | 1912 (10.4) | 1886 (10.4) | 0.001 | 1377.5 (11.4) | 1888.5 (11.4) | 0.000 |
| [61,66) | 2301 (12.5) | 1664 ( 9.2) | 0.108 | 1337.4 (11.1) | 1833.6 (11.1) | 0.000 |
| [66,71) | 2346 (12.8) | 1332 ( 7.3) | 0.182 | 1108.8 ( 9.2) | 1520.2 ( 9.2) | 0.000 |
| [71,76) | 2031 (11.1) | 1010 ( 5.6) | 0.200 | 851.6 ( 7.0) | 1167.4 ( 7.0) | 0.000 |
| [76,81) | 1999 (10.9) | 831 ( 4.6) | 0.238 | 748.2 ( 6.2) | 1025.8 ( 6.2) | 0.000 |
| [81,86) | 1919 (10.4) | 755 ( 4.2) | 0.244 | 672.7 ( 5.6) | 922.3 ( 5.6) | 0.000 |
| [86,91) | 1197 ( 6.5) | 430 ( 2.4) | 0.202 | 395.2 ( 3.3) | 541.8 ( 3.3) | 0.000 |
| [91,96) | 328 ( 1.8) | 135 ( 0.7) | 0.093 | 88.6 ( 0.7) | 121.4 ( 0.7) | 0.000 |
| [96,101) | 39 ( 0.2) | 28 ( 0.2) | 0.014 | 7.6 ( 0.1) | 10.4 ( 0.1) | 0.000 |
| [101,106) | 2 ( 0.0) | 0 ( 0.0) | 0.015 | 0.0 ( 0.0) | 0.0 ( 0.0) | 0.000 |
| [106,111) | 0 ( 0.0) | 0 ( 0.0) | 0.000 | 0.0 ( 0.0) | 0.0 ( 0.0) | 0.000 |
| Sex |  |  |  |  |  |  |
| M | 7466 (40.6) | 7582 (41.7) | -0.022 | 4793.0 (39.7) | 6571.0 (39.7) | 0.000 |
| F | 10906 (59.4) | 10588 (58.3) | 0.022 | 7289.0 (60.3) | 9993.0 (60.3) | 0.000 |
| Vaccination status |  |  |  |  |  |  |
| 1/1, >90d | 0 ( 0.0) | 0 ( 0.0) | 0.000 | 0.0 ( 0.0) | 0.0 ( 0.0) | 0.000 |
| 1/1, <14d | 8 ( 0.0) | 1 ( 0.0) | 0.024 | 0.0 ( 0.0) | 0.0 ( 0.0) | 0.000 |
| 1/1, 14-90d | 1 ( 0.0) | 0 ( 0.0) | 0.010 | 0.0 ( 0.0) | 0.0 ( 0.0) | 0.000 |
| Not vaccinated | 16741 (91.1) | 17347 (95.5) | -0.175 | 11810.0 (97.7) | 16191.0 (97.7) | 0.000 |
| 1/2, >90d | 7 ( 0.0) | 7 ( 0.0) | 0.000 | 0.0 ( 0.0) | 0.0 ( 0.0) | 0.000 |
| 1/2, <14d | 422 ( 2.3) | 295 ( 1.6) | 0.049 | 105.4 ( 0.9) | 144.6 ( 0.9) | 0.000 |
| 1/2, 14-90d | 739 ( 4.0) | 355 ( 2.0) | 0.122 | 157.7 ( 1.3) | 216.3 ( 1.3) | 0.000 |
| 2/2, >90d | 127 ( 0.7) | 30 ( 0.2) | 0.081 | 0.8 ( 0.0) | 1.2 ( 0.0) | 0.000 |
| 2/2, <14d | 97 ( 0.5) | 44 ( 0.2) | 0.046 | 2.5 ( 0.0) | 3.5 ( 0.0) | 0.000 |
| 2/2, 14-90d | 230 ( 1.3) | 91 ( 0.5) | 0.081 | 5.5 ( 0.0) | 7.5 ( 0.0) | 0.000 |
| Period |  |  |  |  |  |  |
| before 10.01.2021. | 11189 (60.9) | 10566 (58.2) | 0.056 | 7623.5 (63.1) | 10451.5 (63.1) | 0.000 |
| 10.01.-15.07.2021. | 6982 (38.0) | 7257 (39.9) | -0.040 | 4415.1 (36.5) | 6052.9 (36.5) | 0.000 |
| after 15.07.2021. | 201 ( 1.1) | 347 ( 1.9) | -0.067 | 43.4 ( 0.4) | 59.6 ( 0.4) | 0.000 |
| CC index |  |  |  |  |  |  |
| 0 | 5613 (30.6) | 11868 (65.3) | -0.742 | 6993.8 (57.9) | 9588.2 (57.9) | 0.000 |
| 1-2 | 8063 (43.9) | 5248 (28.9) | 0.316 | 4411.3 (36.5) | 6047.7 (36.5) | 0.000 |
| 3-4 | 3308 (18.0) | 845 ( 4.7) | 0.431 | 612.8 ( 5.1) | 840.2 ( 5.1) | 0.000 |
| >=5 | 1388 ( 7.6) | 209 ( 1.2) | 0.318 | 64.1 ( 0.5) | 87.9 ( 0.5) | 0.000 |
| Comorbidities |  |  |  |  |  |  |
| Atrial fibrillation | 2670 (14.5) | 661 ( 3.6) | 0.386 | 463.5 ( 3.8) | 635.5 ( 3.8) | 0.000 |
| Autoimmune | 3627 (19.7) | 2270 (12.5) | 0.198 | 1552.1 (12.8) | 2127.9 (12.8) | 0.000 |
| Cancer | 2340 (12.7) | 978 ( 5.4) | 0.258 | 609.0 ( 5.0) | 835.0 ( 5.0) | 0.000 |
| Congestive heart failure | 2098 (11.4) | 462 ( 2.5) | 0.354 | 249.7 ( 2.1) | 342.3 ( 2.1) | 0.000 |
| COPD | 3922 (21.3) | 1979 (10.9) | 0.287 | 1446.2 (12.0) | 1982.8 (12.0) | 0.000 |
| IHD or CVD | 5385 (29.3) | 1518 ( 8.4) | 0.556 | 1469.9 (12.2) | 2015.1 (12.2) | 0.000 |
| Renal disease | 1307 ( 7.1) | 230 ( 1.3) | 0.295 | 80.1 ( 0.7) | 109.9 ( 0.7) | 0.000 |
| Immunocompromised | 987 ( 5.4) | 132 ( 0.7) | 0.273 | 65.4 ( 0.5) | 89.6 ( 0.5) | 0.000 |
| RAAS inhibitors | 10065 (54.8) | 4841 (26.6) | 0.598 | 4462.7 (36.9) | 6118.3 (36.9) | 0.000 |
| Death (composite) | 1889 (10.3) | 803 ( 4.4) | 0.226 | 591.2 ( 4.9) | 897.6 ( 5.4) | -0.020 |
| Death 1 | 1412 ( 7.7) | 585 ( 3.2) | 0.198 | 453.1 ( 3.8) | 662.4 ( 4.0) | -0.011 |
| Death 2 | 1215 ( 6.6) | 519 ( 2.9) | 0.178 | 380.0 ( 3.1) | 560.3 ( 3.4) | -0.011 |
| Death 3 | 1600 ( 8.7) | 666 ( 3.7) | 0.210 | 485.6 ( 4.0) | 736.1 ( 4.4) | -0.018 |
| Hospitalization 1 | 2348 (12.8) | 1160 ( 6.4) | 0.219 | 872.1 ( 7.2) | 1141.0 ( 6.9) | 0.011 |
| Hospitalization 2 | 5015 (27.3) | 2680 (14.7) | 0.312 | 2035.9 (16.9) | 2725.3 (16.5) | 0.010 |

**Table A14**. Sensitivity analysis: possible users vs. non-users (counts/weighted counts, %)

|  | Before matching | | | After matching | | |
| --- | --- | --- | --- | --- | --- | --- |
| Characteristics | Treated | Control | d | Treated | Control | d |
| N | 18170 | 332389 |  | 16795 | 307785 |  |
| Age (5-year bins) |  |  |  |  |  |  |
| [16,21) | 540 ( 3.0) | 22054 ( 6.6) | -0.172 | 1147.6 ( 6.8) | 21031.4 ( 6.8) | 0.000 |
| [21,26) | 759 ( 4.2) | 27711 ( 8.3) | -0.172 | 1447.7 ( 8.6) | 26530.3 ( 8.6) | 0.000 |
| [26,31) | 981 ( 5.4) | 31607 ( 9.5) | -0.157 | 1645.6 ( 9.8) | 30156.4 ( 9.8) | 0.000 |
| [31,36) | 1142 ( 6.3) | 34524 ( 10.4) | -0.149 | 1804.1 ( 10.7) | 33061.9 ( 10.7) | 0.000 |
| [36,41) | 1425 ( 7.8) | 37047 ( 11.1) | -0.113 | 1941.8 ( 11.6) | 35586.2 ( 11.6) | 0.000 |
| [41,46) | 1644 ( 9.0) | 35359 ( 10.6) | -0.053 | 1858.2 ( 11.1) | 34052.8 ( 11.1) | 0.000 |
| [46,51) | 1775 ( 9.8) | 30711 ( 9.2) | 0.018 | 1615.1 ( 9.6) | 29597.9 ( 9.6) | 0.000 |
| [51,56) | 1833 ( 10.1) | 28177 ( 8.5) | 0.056 | 1469.3 ( 8.7) | 26925.7 ( 8.7) | 0.000 |
| [56,61) | 1886 ( 10.4) | 25009 ( 7.5) | 0.100 | 1275.4 ( 7.6) | 23372.6 ( 7.6) | 0.000 |
| [61,66) | 1664 ( 9.2) | 19809 ( 6.0) | 0.121 | 981.2 ( 5.8) | 17981.8 ( 5.8) | 0.000 |
| [66,71) | 1332 ( 7.3) | 14082 ( 4.2) | 0.133 | 641.4 ( 3.8) | 11754.6 ( 3.8) | 0.000 |
| [71,76) | 1010 ( 5.6) | 9476 ( 2.9) | 0.135 | 396.7 ( 2.4) | 7269.3 ( 2.4) | 0.000 |
| [76,81) | 831 ( 4.6) | 6813 ( 2.0) | 0.141 | 249.7 ( 1.5) | 4576.3 ( 1.5) | 0.000 |
| [81,86) | 755 ( 4.2) | 5529 ( 1.7) | 0.149 | 194.9 ( 1.2) | 3571.1 ( 1.2) | 0.000 |
| [86,91) | 430 ( 2.4) | 3158 ( 1.0) | 0.111 | 94.8 ( 0.6) | 1738.2 ( 0.6) | 0.000 |
| [91,96) | 135 ( 0.7) | 1101 ( 0.3) | 0.056 | 28.2 ( 0.2) | 516.8 ( 0.2) | 0.000 |
| [96,101) | 28 ( 0.2) | 214 ( 0.1) | 0.027 | 3.4 ( 0.0) | 61.6 ( 0.0) | 0.000 |
| [101,106) | 0 ( 0.0) | 7 ( 0.0) | -0.006 | 0.0 ( 0.0) | 0.0 ( 0.0) | 0.000 |
| [106,111) | 0 ( 0.0) | 1 ( 0.0) | -0.002 | 0.0 ( 0.0) | 0.0 ( 0.0) | 0.000 |
| Sex |  |  |  |  |  |  |
| M | 7582 ( 41.7) | 162983 ( 49.0) | -0.147 | 8099.7 ( 48.2) | 148434.3 ( 48.2) | 0.000 |
| F | 10588 ( 58.3) | 169406 ( 51.0) | 0.147 | 8695.3 ( 51.8) | 159350.7 ( 51.8) | 0.000 |
| Vaccination status |  |  |  |  |  |  |
| 1/1, >90d | 0 ( 0.0) | 2 ( 0.0) | -0.003 | 0.0 ( 0.0) | 0.0 ( 0.0) | 0.000 |
| 1/1, <14d | 1 ( 0.0) | 47 ( 0.0) | -0.009 | 0.1 ( 0.0) | 1.9 ( 0.0) | 0.000 |
| 1/1, 14-90d | 0 ( 0.0) | 48 ( 0.0) | -0.017 | 0.0 ( 0.0) | 0.0 ( 0.0) | 0.000 |
| Not vaccinated | 17347 ( 95.5) | 322899 ( 97.1) | -0.089 | 16541.6 ( 98.5) | 303140.4 ( 98.5) | 0.000 |
| 1/2, >90d | 7 ( 0.0) | 35 ( 0.0) | 0.018 | 0.0 ( 0.0) | 0.0 ( 0.0) | 0.000 |
| 1/2, <14d | 295 ( 1.6) | 3549 ( 1.1) | 0.048 | 117.4 ( 0.7) | 2150.6 ( 0.7) | 0.000 |
| 1/2, 14-90d | 355 ( 2.0) | 3880 ( 1.2) | 0.063 | 114.1 ( 0.7) | 2091.9 ( 0.7) | 0.000 |
| 2/2, >90d | 30 ( 0.2) | 363 ( 0.1) | 0.015 | 1.4 ( 0.0) | 25.6 ( 0.0) | 0.000 |
| 2/2, <14d | 44 ( 0.2) | 420 ( 0.1) | 0.027 | 4.3 ( 0.0) | 78.7 ( 0.0) | 0.000 |
| 2/2, 14-90d | 91 ( 0.5) | 1146 ( 0.3) | 0.024 | 16.1 ( 0.1) | 295.9 ( 0.1) | 0.000 |
| Period |  |  |  |  |  |  |
| before 10.01.2021. | 10566 ( 58.2) | 192599 ( 57.9) | 0.004 | 9891.2 ( 58.9) | 181265.8 ( 58.9) | 0.000 |
| 10.01.-15.07.2021. | 7257 ( 39.9) | 132866 ( 40.0) | -0.001 | 6607.6 ( 39.3) | 121090.4 ( 39.3) | 0.000 |
| after 15.07.2021. | 347 ( 1.9) | 6924 ( 2.1) | -0.012 | 296.2 ( 1.8) | 5428.8 ( 1.8) | 0.000 |
| Comorbidities |  |  |  |  |  |  |
| Acute myocardial infarction | 198 ( 1.1) | 2103 ( 0.6) | 0.049 | 24.9 ( 0.1) | 457.1 ( 0.1) | 0.000 |
| Congestive heart failure | 462 ( 2.5) | 3380 ( 1.0) | 0.116 | 31.8 ( 0.2) | 582.2 ( 0.2) | 0.000 |
| Peripheral vascular disease | 301 ( 1.7) | 2579 ( 0.8) | 0.080 | 30.6 ( 0.2) | 560.4 ( 0.2) | 0.000 |
| Cerebrovascular disease | 604 ( 3.3) | 5189 ( 1.6) | 0.114 | 104.7 ( 0.6) | 1918.3 ( 0.6) | 0.000 |
| Alzheimer’s dementia | 249 ( 1.4) | 2294 ( 0.7) | 0.067 | 41.8 ( 0.2) | 766.2 ( 0.2) | 0.000 |
| Other dementia | 1979 ( 10.9) | 21520 ( 6.5) | 0.157 | 916.1 ( 5.5) | 16787.9 ( 5.5) | 0.000 |
| Mild liver disease | 340 ( 1.9) | 3363 ( 1.0) | 0.072 | 109.4 ( 0.7) | 2004.6 ( 0.7) | 0.000 |
| Diabetes without complications | 2449 ( 13.5) | 21896 ( 6.6) | 0.231 | 829.0 ( 4.9) | 15193.0 ( 4.9) | 0.000 |
| Diabetes with complications | 202 ( 1.1) | 1735 ( 0.5) | 0.066 | 20.3 ( 0.1) | 371.7 ( 0.1) | 0.000 |
| Renal disease | 252 ( 1.4) | 1904 ( 0.6) | 0.083 | 14.9 ( 0.1) | 272.1 ( 0.1) | 0.000 |
| Cancer | 971 ( 5.3) | 10088 ( 3.0) | 0.115 | 322.7 ( 1.9) | 5913.3 ( 1.9) | 0.000 |
| Moderate to severe liver disease | 7 ( 0.0) | 66 ( 0.0) | 0.011 | 0.0 ( 0.0) | 0.0 ( 0.0) | 0.000 |
| Metastatic cancer | 52 ( 0.3) | 375 ( 0.1) | 0.039 | 2.0 ( 0.0) | 36.0 ( 0.0) | 0.000 |
| HIV/AIDS | 1 ( 0.0) | 21 ( 0.0) | -0.001 | 0.1 ( 0.0) | 1.9 ( 0.0) | 0.000 |
| RAAS inhibitors | 4841 ( 26.6) | 43193 ( 13.0) | 0.348 | 1875.0 ( 11.2) | 34362.0 ( 11.2) | 0.000 |
| Beta blockers | 2450 ( 13.5) | 22235 ( 6.7) | 0.227 | 796.3 ( 4.7) | 14592.7 ( 4.7) | 0.000 |
| Diuretics | 1872 ( 10.3) | 14387 ( 4.3) | 0.231 | 451.7 ( 2.7) | 8277.3 ( 2.7) | 0.000 |
| Immunosuppressant drugs | 75 ( 0.4) | 800 ( 0.2) | 0.030 | 6.9 ( 0.0) | 126.1 ( 0.0) | 0.000 |
| Systemic corticosteroids | 72 ( 0.4) | 617 ( 0.2) | 0.039 | 4.6 ( 0.0) | 83.4 ( 0.0) | 0.000 |
| Antineoplastic drugs | 118 ( 0.6) | 1241 ( 0.4) | 0.039 | 20.9 ( 0.1) | 382.1 ( 0.1) | 0.000 |
| Antiviral agents | 16 ( 0.1) | 215 ( 0.1) | 0.008 | 0.8 ( 0.0) | 14.2 ( 0.0) | 0.000 |
| Death (composite) | 803 ( 4.4) | 5599 ( 1.7) | 0.160 | 228.1 ( 1.4) | 3429.1 ( 1.1) | 0.014 |
| Death 1 | 585 ( 3.2) | 4366 ( 1.3) | 0.128 | 176.7 ( 1.1) | 2762.8 ( 0.9) | 0.010 |
| Death 2 | 519 ( 2.9) | 3698 ( 1.1) | 0.125 | 153.4 ( 0.9) | 2237.9 ( 0.7) | 0.013 |
| Death 3 | 666 ( 3.7) | 4519 ( 1.4) | 0.148 | 183.0 ( 1.1) | 2681.0 ( 0.9) | 0.014 |
| Hospitalization 1 | 1160 ( 6.4) | 10905 ( 3.3) | 0.145 | 557.7 ( 3.3) | 8146.0 ( 2.6) | 0.032 |
| Hospitalization 2 | 2680 ( 14.7) | 26965 ( 8.1) | 0.210 | 1396.5 ( 8.3) | 21199.4 ( 6.9) | 0.045 |

**Table A15**. Sensitivity analysis: users (all) vs. non-users (counts/weighted counts, %)

|  | Before matching | | | After matching | | |
| --- | --- | --- | --- | --- | --- | --- |
| Characteristics | Treated | Control | d | Treated | Control | d |
| N | 55098 | 332389 |  | 40653 | 317678 |  |
| Age (5-year bins) |  |  |  |  |  |  |
| [16,21) | 401 ( 0.7) | 22054 ( 6.6) | -0.318 | 2493.1 ( 6.1) | 19481.9 ( 6.1) | 0.000 |
| [21,26) | 782 ( 1.4) | 27711 ( 8.3) | -0.325 | 3172.5 ( 7.8) | 24791.5 ( 7.8) | 0.000 |
| [26,31) | 1137 ( 2.1) | 31607 ( 9.5) | -0.323 | 3648.6 ( 9.0) | 28511.4 ( 9.0) | 0.000 |
| [31,36) | 1703 ( 3.1) | 34524 ( 10.4) | -0.294 | 4047.0 ( 10.0) | 31625.0 ( 10.0) | 0.000 |
| [36,41) | 2571 ( 4.7) | 37047 ( 11.1) | -0.242 | 4416.0 ( 10.9) | 34508.0 ( 10.9) | 0.000 |
| [41,46) | 3335 ( 6.1) | 35359 ( 10.6) | -0.166 | 4279.5 ( 10.5) | 33441.5 ( 10.5) | 0.000 |
| [46,51) | 4243 ( 7.7) | 30711 ( 9.2) | -0.055 | 3831.9 ( 9.4) | 29944.1 ( 9.4) | 0.000 |
| [51,56) | 5301 ( 9.6) | 28177 ( 8.5) | 0.040 | 3612.5 ( 8.9) | 28229.5 ( 8.9) | 0.000 |
| [56,61) | 6107 ( 11.1) | 25009 ( 7.5) | 0.123 | 3267.2 ( 8.0) | 25530.8 ( 8.0) | 0.000 |
| [61,66) | 6560 ( 11.9) | 19809 ( 6.0) | 0.210 | 2649.6 ( 6.5) | 20705.4 ( 6.5) | 0.000 |
| [66,71) | 6134 ( 11.1) | 14082 ( 4.2) | 0.261 | 1895.1 ( 4.7) | 14808.9 ( 4.7) | 0.000 |
| [71,76) | 5050 ( 9.2) | 9476 ( 2.9) | 0.268 | 1243.8 ( 3.1) | 9719.2 ( 3.1) | 0.000 |
| [76,81) | 4480 ( 8.1) | 6813 ( 2.0) | 0.279 | 883.0 ( 2.2) | 6900.0 ( 2.2) | 0.000 |
| [81,86) | 4104 ( 7.4) | 5529 ( 1.7) | 0.280 | 712.4 ( 1.8) | 5566.6 ( 1.8) | 0.000 |
| [86,91) | 2400 ( 4.4) | 3158 ( 1.0) | 0.213 | 382.0 ( 0.9) | 2985.0 ( 0.9) | 0.000 |
| [91,96) | 693 ( 1.3) | 1101 ( 0.3) | 0.105 | 106.3 ( 0.3) | 830.7 ( 0.3) | 0.000 |
| [96,101) | 95 ( 0.2) | 214 ( 0.1) | 0.031 | 12.6 ( 0.0) | 98.4 ( 0.0) | 0.000 |
| [101,106) | 2 ( 0.0) | 7 ( 0.0) | 0.003 | 0.0 ( 0.0) | 0.0 ( 0.0) | 0.000 |
| [106,111) | 0 ( 0.0) | 1 ( 0.0) | -0.002 | 0.0 ( 0.0) | 0.0 ( 0.0) | 0.000 |
| Sex |  |  |  |  |  |  |
| M | 22645 ( 41.1) | 162983 ( 49.0) | -0.160 | 19383.8 ( 47.7) | 151472.2 ( 47.7) | 0.000 |
| F | 32453 ( 58.9) | 169406 ( 51.0) | 0.160 | 21269.2 ( 52.3) | 166205.8 ( 52.3) | 0.000 |
| Vaccination status |  |  |  |  |  |  |
| 1/1, >90d | 2 ( 0.0) | 2 ( 0.0) | 0.007 | 0.0 ( 0.0) | 0.0 ( 0.0) | 0.000 |
| 1/1, <14d | 12 ( 0.0) | 47 ( 0.0) | 0.006 | 0.3 ( 0.0) | 2.7 ( 0.0) | 0.000 |
| 1/1, 14-90d | 5 ( 0.0) | 48 ( 0.0) | -0.005 | 0.0 ( 0.0) | 0.0 ( 0.0) | 0.000 |
| Not vaccinated | 51111 ( 92.8) | 322899 ( 97.1) | -0.201 | 39861.2 ( 98.1) | 311490.8 ( 98.1) | 0.000 |
| 1/2, >90d | 22 ( 0.0) | 35 ( 0.0) | 0.019 | 0.0 ( 0.0) | 0.0 ( 0.0) | 0.000 |
| 1/2, <14d | 1121 ( 2.0) | 3549 ( 1.1) | 0.078 | 339.2 ( 0.8) | 2650.8 ( 0.8) | 0.000 |
| 1/2, 14-90d | 1791 ( 3.3) | 3880 ( 1.2) | 0.142 | 372.5 ( 0.9) | 2910.5 ( 0.9) | 0.000 |
| 2/2, >90d | 239 ( 0.4) | 363 ( 0.1) | 0.062 | 10.3 ( 0.0) | 80.7 ( 0.0) | 0.000 |
| 2/2, <14d | 242 ( 0.4) | 420 ( 0.1) | 0.059 | 13.7 ( 0.0) | 107.3 ( 0.0) | 0.000 |
| 2/2, 14-90d | 553 ( 1.0) | 1146 ( 0.3) | 0.081 | 55.7 ( 0.1) | 435.3 ( 0.1) | 0.000 |
| Period |  |  |  |  |  |  |
| before 10.01.2021. | 32317 ( 58.7) | 192599 ( 57.9) | 0.014 | 24002.4 ( 59.0) | 187563.6 ( 59.0) | 0.000 |
| 10.01.-15.07.2021. | 22102 ( 40.1) | 132866 ( 40.0) | 0.003 | 15979.3 ( 39.3) | 124868.7 ( 39.3) | 0.000 |
| after 15.07.2021. | 679 ( 1.2) | 6924 ( 2.1) | -0.067 | 671.3 ( 1.7) | 5245.7 ( 1.7) | 0.000 |
| Comorbidities |  |  |  |  |  |  |
| Acute myocardial infarction | 2830 ( 5.1) | 2103 ( 0.6) | 0.272 | 231.4 ( 0.6) | 1808.6 ( 0.6) | 0.000 |
| Congestive heart failure | 4234 ( 7.7) | 3380 ( 1.0) | 0.331 | 265.6 ( 0.7) | 2075.4 ( 0.7) | 0.000 |
| Peripheral vascular disease | 2238 ( 4.1) | 2579 ( 0.8) | 0.215 | 166.8 ( 0.4) | 1303.2 ( 0.4) | 0.000 |
| Cerebrovascular disease | 4552 ( 8.3) | 5189 ( 1.6) | 0.314 | 533.1 ( 1.3) | 4165.9 ( 1.3) | 0.000 |
| Alzheimer’s dementia | 1540 ( 2.8) | 2294 ( 0.7) | 0.161 | 187.6 ( 0.5) | 1466.4 ( 0.5) | 0.000 |
| Other dementia | 9700 ( 17.6) | 21520 ( 6.5) | 0.347 | 2676.2 ( 6.6) | 20912.8 ( 6.6) | 0.000 |
| Mild liver disease | 1724 ( 3.1) | 3363 ( 1.0) | 0.149 | 338.3 ( 0.8) | 2643.7 ( 0.8) | 0.000 |
| Diabetes without complications | 11803 ( 21.4) | 21896 ( 6.6) | 0.438 | 2611.1 ( 6.4) | 20403.9 ( 6.4) | 0.000 |
| Diabetes with complications | 1499 ( 2.7) | 1735 ( 0.5) | 0.175 | 99.5 ( 0.2) | 777.5 ( 0.2) | 0.000 |
| Renal disease | 2805 ( 5.1) | 1904 ( 0.6) | 0.275 | 112.0 ( 0.3) | 875.0 ( 0.3) | 0.000 |
| Cancer | 6173 ( 11.2) | 10088 ( 3.0) | 0.322 | 1116.0 ( 2.7) | 8721.0 ( 2.7) | 0.000 |
| Moderate to severe liver disease | 127 ( 0.2) | 66 ( 0.0) | 0.060 | 0.5 ( 0.0) | 3.5 ( 0.0) | 0.000 |
| Metastatic cancer | 611 ( 1.1) | 375 ( 0.1) | 0.128 | 25.3 ( 0.1) | 197.7 ( 0.1) | 0.000 |
| HIV/AIDS | 8 ( 0.0) | 21 ( 0.0) | 0.008 | 0.0 ( 0.0) | 0.0 ( 0.0) | 0.000 |
| RAAS inhibitors | 23376 ( 42.4) | 43193 ( 13.0) | 0.696 | 5933.6 ( 14.6) | 46367.4 ( 14.6) | 0.000 |
| Beta blockers | 16969 ( 30.8) | 22235 ( 6.7) | 0.650 | 3053.4 ( 7.5) | 23860.6 ( 7.5) | 0.000 |
| Diuretics | 13757 ( 25.0) | 14387 ( 4.3) | 0.610 | 1912.2 ( 4.7) | 14942.8 ( 4.7) | 0.000 |
| Immunosuppressant drugs | 1454 ( 2.6) | 800 ( 0.2) | 0.202 | 73.6 ( 0.2) | 575.4 ( 0.2) | 0.000 |
| Systemic corticosteroids | 2535 ( 4.6) | 617 ( 0.2) | 0.292 | 76.7 ( 0.2) | 599.3 ( 0.2) | 0.000 |
| Antineoplastic drugs | 975 ( 1.8) | 1241 ( 0.4) | 0.136 | 95.6 ( 0.2) | 747.4 ( 0.2) | 0.000 |
| Antiviral agents | 417 ( 0.8) | 215 ( 0.1) | 0.108 | 5.6 ( 0.0) | 43.4 ( 0.0) | 0.000 |
| Death (composite) | 4356 ( 7.9) | 5599 ( 1.7) | 0.294 | 747.4 ( 1.8) | 5063.8 ( 1.6) | 0.012 |
| Death 1 | 3195 ( 5.8) | 4366 ( 1.3) | 0.244 | 588.8 ( 1.4) | 4008.6 ( 1.3) | 0.010 |
| Death 2 | 2839 ( 5.2) | 3698 ( 1.1) | 0.234 | 487.1 ( 1.2) | 3296.7 ( 1.0) | 0.009 |
| Death 3 | 3670 ( 6.7) | 4519 ( 1.4) | 0.273 | 613.6 ( 1.5) | 4063.6 ( 1.3) | 0.012 |
| Hospitalization 1 | 5824 ( 10.6) | 10905 ( 3.3) | 0.290 | 1842.0 ( 4.5) | 9831.0 ( 3.1) | 0.057 |
| Hospitalization 2 | 12735 ( 23.1) | 26965 ( 8.1) | 0.422 | 4343.4 ( 10.7) | 25487.1 ( 8.0) | 0.075 |

**Table A16**. Sensitivity analysis: users (1-3 prescriptions) vs. non-users (counts/weighted counts, %)

|  | Before matching | | | After matching | | |
| --- | --- | --- | --- | --- | --- | --- |
| Characteristics | Treated | Control | d | Treated | Control | d |
| N | 18996 | 332389 |  | 16157 | 308555 |  |
| Age (5-year bins) |  |  |  |  |  |  |
| [16,21) | 305 ( 1.6) | 22054 ( 6.6) | -0.255 | 1084.9 ( 6.7) | 20719.1 ( 6.7) | 0.000 |
| [21,26) | 515 ( 2.7) | 27711 ( 8.3) | -0.248 | 1374.2 ( 8.5) | 26243.8 ( 8.5) | 0.000 |
| [26,31) | 726 ( 3.8) | 31607 ( 9.5) | -0.230 | 1570.9 ( 9.7) | 29999.1 ( 9.7) | 0.000 |
| [31,36) | 1002 ( 5.3) | 34524 ( 10.4) | -0.191 | 1734.6 ( 10.7) | 33125.4 ( 10.7) | 0.000 |
| [36,41) | 1378 ( 7.3) | 37047 ( 11.1) | -0.135 | 1873.1 ( 11.6) | 35770.9 ( 11.6) | 0.000 |
| [41,46) | 1596 ( 8.4) | 35359 ( 10.6) | -0.076 | 1783.4 ( 11.0) | 34058.6 ( 11.0) | 0.000 |
| [46,51) | 1825 ( 9.6) | 30711 ( 9.2) | 0.013 | 1557.6 ( 9.6) | 29745.4 ( 9.6) | 0.000 |
| [51,56) | 2101 ( 11.1) | 28177 ( 8.5) | 0.087 | 1428.9 ( 8.8) | 27288.1 ( 8.8) | 0.000 |
| [56,61) | 2099 ( 11.0) | 25009 ( 7.5) | 0.122 | 1238.1 ( 7.7) | 23644.9 ( 7.7) | 0.000 |
| [61,66) | 2054 ( 10.8) | 19809 ( 6.0) | 0.176 | 955.9 ( 5.9) | 18256.1 ( 5.9) | 0.000 |
| [66,71) | 1680 ( 8.8) | 14082 ( 4.2) | 0.187 | 637.2 ( 3.9) | 12167.8 ( 3.9) | 0.000 |
| [71,76) | 1310 ( 6.9) | 9476 ( 2.9) | 0.189 | 388.9 ( 2.4) | 7427.1 ( 2.4) | 0.000 |
| [76,81) | 1012 ( 5.3) | 6813 ( 2.0) | 0.175 | 239.9 ( 1.5) | 4582.1 ( 1.5) | 0.000 |
| [81,86) | 826 ( 4.3) | 5529 ( 1.7) | 0.158 | 178.1 ( 1.1) | 3400.9 ( 1.1) | 0.000 |
| [86,91) | 433 ( 2.3) | 3158 ( 1.0) | 0.106 | 90.0 ( 0.6) | 1718.0 ( 0.6) | 0.000 |
| [91,96) | 118 ( 0.6) | 1101 ( 0.3) | 0.042 | 20.6 ( 0.1) | 392.4 ( 0.1) | 0.000 |
| [96,101) | 16 ( 0.1) | 214 ( 0.1) | 0.007 | 0.8 ( 0.0) | 15.2 ( 0.0) | 0.000 |
| [101,106) | 0 ( 0.0) | 7 ( 0.0) | -0.006 | 0.0 ( 0.0) | 0.0 ( 0.0) | 0.000 |
| [106,111) | 0 ( 0.0) | 1 ( 0.0) | -0.002 | 0.0 ( 0.0) | 0.0 ( 0.0) | 0.000 |
| Sex |  |  |  |  |  |  |
| M | 7707 ( 40.6) | 162983 ( 49.0) | -0.171 | 7778.4 ( 48.1) | 148545.6 ( 48.1) | 0.000 |
| F | 11289 ( 59.4) | 169406 ( 51.0) | 0.171 | 8378.6 ( 51.9) | 160009.4 ( 51.9) | 0.000 |
| Vaccination status |  |  |  |  |  |  |
| 1/1, >90d | 1 ( 0.0) | 2 ( 0.0) | 0.009 | 0.0 ( 0.0) | 0.0 ( 0.0) | 0.000 |
| 1/1, <14d | 2 ( 0.0) | 47 ( 0.0) | -0.003 | 0.1 ( 0.0) | 2.9 ( 0.0) | 0.000 |
| 1/1, 14-90d | 2 ( 0.0) | 48 ( 0.0) | -0.004 | 0.0 ( 0.0) | 0.0 ( 0.0) | 0.000 |
| Not vaccinated | 18000 ( 94.8) | 322899 ( 97.1) | -0.121 | 15906.8 ( 98.5) | 303776.2 ( 98.5) | 0.000 |
| 1/2, >90d | 7 ( 0.0) | 35 ( 0.0) | 0.017 | 0.0 ( 0.0) | 0.0 ( 0.0) | 0.000 |
| 1/2, <14d | 332 ( 1.7) | 3549 ( 1.1) | 0.058 | 119.9 ( 0.7) | 2289.1 ( 0.7) | 0.000 |
| 1/2, 14-90d | 431 ( 2.3) | 3880 ( 1.2) | 0.085 | 108.5 ( 0.7) | 2071.5 ( 0.7) | 0.000 |
| 2/2, >90d | 36 ( 0.2) | 363 ( 0.1) | 0.021 | 2.3 ( 0.0) | 44.7 ( 0.0) | 0.000 |
| 2/2, <14d | 56 ( 0.3) | 420 ( 0.1) | 0.037 | 3.8 ( 0.0) | 73.2 ( 0.0) | 0.000 |
| 2/2, 14-90d | 129 ( 0.7) | 1146 ( 0.3) | 0.047 | 15.6 ( 0.1) | 297.4 ( 0.1) | 0.000 |
| Period |  |  |  |  |  |  |
| before 10.01.2021. | 10627 ( 55.9) | 192599 ( 57.9) | -0.040 | 9501.1 ( 58.8) | 181444.9 ( 58.8) | 0.000 |
| 10.01.-15.07.2021. | 8105 ( 42.7) | 132866 ( 40.0) | 0.055 | 6377.9 ( 39.5) | 121800.1 ( 39.5) | 0.000 |
| after 15.07.2021. | 264 ( 1.4) | 6924 ( 2.1) | -0.053 | 278.0 ( 1.7) | 5310.0 ( 1.7) | 0.000 |
| Comorbidities |  |  |  |  |  |  |
| Acute myocardial infarction | 515 ( 2.7) | 2103 ( 0.6) | 0.163 | 41.2 ( 0.3) | 786.8 ( 0.3) | 0.000 |
| Congestive heart failure | 783 ( 4.1) | 3380 ( 1.0) | 0.197 | 36.9 ( 0.2) | 704.1 ( 0.2) | 0.000 |
| Peripheral vascular disease | 476 ( 2.5) | 2579 ( 0.8) | 0.136 | 33.0 ( 0.2) | 630.0 ( 0.2) | 0.000 |
| Cerebrovascular disease | 980 ( 5.2) | 5189 ( 1.6) | 0.201 | 117.2 ( 0.7) | 2238.8 ( 0.7) | 0.000 |
| Alzheimer’s dementia | 266 ( 1.4) | 2294 ( 0.7) | 0.070 | 34.0 ( 0.2) | 649.0 ( 0.2) | 0.000 |
| Other dementia | 2626 ( 13.8) | 21520 ( 6.5) | 0.245 | 906.4 ( 5.6) | 17310.6 ( 5.6) | 0.000 |
| Mild liver disease | 502 ( 2.6) | 3363 ( 1.0) | 0.122 | 106.8 ( 0.7) | 2039.2 ( 0.7) | 0.000 |
| Diabetes without complications | 2729 ( 14.4) | 21896 ( 6.6) | 0.256 | 769.6 ( 4.8) | 14697.4 ( 4.8) | 0.000 |
| Diabetes with complications | 304 ( 1.6) | 1735 ( 0.5) | 0.105 | 18.1 ( 0.1) | 345.9 ( 0.1) | 0.000 |
| Renal disease | 442 ( 2.3) | 1904 ( 0.6) | 0.147 | 12.3 ( 0.1) | 234.7 ( 0.1) | 0.000 |
| Cancer | 1848 ( 9.7) | 10088 ( 3.0) | 0.276 | 348.3 ( 2.2) | 6651.7 ( 2.2) | 0.000 |
| Moderate to severe liver disease | 33 ( 0.2) | 66 ( 0.0) | 0.049 | 0.1 ( 0.0) | 1.9 ( 0.0) | 0.000 |
| Metastatic cancer | 183 ( 1.0) | 375 ( 0.1) | 0.116 | 6.0 ( 0.0) | 115.0 ( 0.0) | 0.000 |
| HIV/AIDS | 2 ( 0.0) | 21 ( 0.0) | 0.005 | 0.0 ( 0.0) | 0.0 ( 0.0) | 0.000 |
| RAAS inhibitors | 5450 ( 28.7) | 43193 ( 13.0) | 0.394 | 1807.7 ( 11.2) | 34522.3 ( 11.2) | 0.000 |
| Beta blockers | 3309 ( 17.4) | 22235 ( 6.7) | 0.334 | 797.6 ( 4.9) | 15232.4 ( 4.9) | 0.000 |
| Diuretics | 2567 ( 13.5) | 14387 ( 4.3) | 0.326 | 436.3 ( 2.7) | 8332.7 ( 2.7) | 0.000 |
| Immunosuppressant drugs | 203 ( 1.1) | 800 ( 0.2) | 0.103 | 13.3 ( 0.1) | 253.7 ( 0.1) | 0.000 |
| Systemic corticosteroids | 539 ( 2.8) | 617 ( 0.2) | 0.219 | 15.4 ( 0.1) | 294.6 ( 0.1) | 0.000 |
| Antineoplastic drugs | 233 ( 1.2) | 1241 ( 0.4) | 0.096 | 24.6 ( 0.2) | 470.4 ( 0.2) | 0.000 |
| Antiviral agents | 80 ( 0.4) | 215 ( 0.1) | 0.072 | 1.1 ( 0.0) | 20.9 ( 0.0) | 0.000 |
| Death (composite) | 1070 ( 5.6) | 5599 ( 1.7) | 0.211 | 225.7 ( 1.4) | 3369.8 ( 1.1) | 0.016 |
| Death 1 | 761 ( 4.0) | 4366 ( 1.3) | 0.168 | 183.2 ( 1.1) | 2703.0 ( 0.9) | 0.016 |
| Death 2 | 704 ( 3.7) | 3698 ( 1.1) | 0.170 | 151.8 ( 0.9) | 2222.7 ( 0.7) | 0.014 |
| Death 3 | 900 ( 4.7) | 4519 ( 1.4) | 0.197 | 183.2 ( 1.1) | 2626.1 ( 0.9) | 0.017 |
| Hospitalization 1 | 1646 ( 8.7) | 10905 ( 3.3) | 0.229 | 706.8 ( 4.4) | 8214.2 ( 2.7) | 0.073 |
| Hospitalization 2 | 3624 ( 19.1) | 26965 ( 8.1) | 0.324 | 1639.3 ( 10.1) | 21297.6 ( 6.9) | 0.096 |

**Table A17**. Sensitivity analysis: users (4-7 prescriptions) vs. non-users (counts/weighted counts, %)

|  | Before matching | | | After matching | | |
| --- | --- | --- | --- | --- | --- | --- |
| Characteristics | Treated | Control | d | Treated | Control | d |
| N | 17730 | 332389 |  | 12812 | 306627 |  |
| Age (5-year bins) |  |  |  |  |  |  |
| [16,21) | 74 ( 0.4) | 22054 ( 6.6) | -0.342 | 841.4 ( 6.6) | 20136.6 ( 6.6) | 0.000 |
| [21,26) | 168 ( 0.9) | 27711 ( 8.3) | -0.357 | 1060.5 ( 8.3) | 25381.5 ( 8.3) | 0.000 |
| [26,31) | 251 ( 1.4) | 31607 ( 9.5) | -0.362 | 1234.0 ( 9.6) | 29534.0 ( 9.6) | 0.000 |
| [31,36) | 442 ( 2.5) | 34524 ( 10.4) | -0.326 | 1367.6 ( 10.7) | 32729.4 ( 10.7) | 0.000 |
| [36,41) | 707 ( 4.0) | 37047 ( 11.1) | -0.273 | 1462.9 ( 11.4) | 35010.1 ( 11.4) | 0.000 |
| [41,46) | 1006 ( 5.7) | 35359 ( 10.6) | -0.182 | 1393.0 ( 10.9) | 33339.0 ( 10.9) | 0.000 |
| [46,51) | 1352 ( 7.6) | 30711 ( 9.2) | -0.058 | 1230.7 ( 9.6) | 29454.3 ( 9.6) | 0.000 |
| [51,56) | 1727 ( 9.7) | 28177 ( 8.5) | 0.044 | 1124.5 ( 8.8) | 26911.5 ( 8.8) | 0.000 |
| [56,61) | 2096 ( 11.8) | 25009 ( 7.5) | 0.146 | 996.0 ( 7.8) | 23836.0 ( 7.8) | 0.000 |
| [61,66) | 2205 ( 12.4) | 19809 ( 6.0) | 0.226 | 770.8 ( 6.0) | 18447.2 ( 6.0) | 0.000 |
| [66,71) | 2108 ( 11.9) | 14082 ( 4.2) | 0.284 | 526.5 ( 4.1) | 12600.5 ( 4.1) | 0.000 |
| [71,76) | 1709 ( 9.6) | 9476 ( 2.9) | 0.283 | 323.1 ( 2.5) | 7733.9 ( 2.5) | 0.000 |
| [76,81) | 1469 ( 8.3) | 6813 ( 2.0) | 0.285 | 209.0 ( 1.6) | 5002.0 ( 1.6) | 0.000 |
| [81,86) | 1359 ( 7.7) | 5529 ( 1.7) | 0.288 | 164.4 ( 1.3) | 3934.6 ( 1.3) | 0.000 |
| [86,91) | 770 ( 4.3) | 3158 ( 1.0) | 0.213 | 83.1 ( 0.6) | 1987.9 ( 0.6) | 0.000 |
| [91,96) | 247 ( 1.4) | 1101 ( 0.3) | 0.115 | 21.8 ( 0.2) | 521.2 ( 0.2) | 0.000 |
| [96,101) | 40 ( 0.2) | 214 ( 0.1) | 0.042 | 2.8 ( 0.0) | 67.2 ( 0.0) | 0.000 |
| [101,106) | 0 ( 0.0) | 7 ( 0.0) | -0.006 | 0.0 ( 0.0) | 0.0 ( 0.0) | 0.000 |
| [106,111) | 0 ( 0.0) | 1 ( 0.0) | -0.002 | 0.0 ( 0.0) | 0.0 ( 0.0) | 0.000 |
| Sex |  |  |  |  |  |  |
| M | 7472 ( 42.1) | 162983 ( 49.0) | -0.139 | 6195.7 ( 48.4) | 148280.3 ( 48.4) | 0.000 |
| F | 10258 ( 57.9) | 169406 ( 51.0) | 0.139 | 6616.3 ( 51.6) | 158346.7 ( 51.6) | 0.000 |
| Vaccination status |  |  |  |  |  |  |
| 1/1, >90d | 1 ( 0.0) | 2 ( 0.0) | 0.009 | 0.0 ( 0.0) | 0.0 ( 0.0) | 0.000 |
| 1/1, <14d | 2 ( 0.0) | 47 ( 0.0) | -0.003 | 0.0 ( 0.0) | 0.0 ( 0.0) | 0.000 |
| 1/1, 14-90d | 2 ( 0.0) | 48 ( 0.0) | -0.003 | 0.0 ( 0.0) | 0.0 ( 0.0) | 0.000 |
| Not vaccinated | 16370 ( 92.3) | 322899 ( 97.1) | -0.217 | 12637.4 ( 98.6) | 302447.6 ( 98.6) | 0.000 |
| 1/2, >90d | 8 ( 0.0) | 35 ( 0.0) | 0.021 | 0.0 ( 0.0) | 0.0 ( 0.0) | 0.000 |
| 1/2, <14d | 367 ( 2.1) | 3549 ( 1.1) | 0.081 | 75.9 ( 0.6) | 1817.1 ( 0.6) | 0.000 |
| 1/2, 14-90d | 621 ( 3.5) | 3880 ( 1.2) | 0.155 | 88.3 ( 0.7) | 2112.7 ( 0.7) | 0.000 |
| 2/2, >90d | 76 ( 0.4) | 363 ( 0.1) | 0.062 | 1.5 ( 0.0) | 36.5 ( 0.0) | 0.000 |
| 2/2, <14d | 89 ( 0.5) | 420 ( 0.1) | 0.067 | 1.2 ( 0.0) | 29.8 ( 0.0) | 0.000 |
| 2/2, 14-90d | 194 ( 1.1) | 1146 ( 0.3) | 0.089 | 7.7 ( 0.1) | 183.3 ( 0.1) | 0.000 |
| Period |  |  |  |  |  |  |
| before 10.01.2021. | 10501 ( 59.2) | 192599 ( 57.9) | 0.026 | 7642.2 ( 59.6) | 182899.8 ( 59.6) | 0.000 |
| 10.01.-15.07.2021. | 7015 ( 39.6) | 132866 ( 40.0) | -0.008 | 5026.0 ( 39.2) | 120285.0 ( 39.2) | 0.000 |
| after 15.07.2021. | 214 ( 1.2) | 6924 ( 2.1) | -0.069 | 143.8 ( 1.1) | 3442.2 ( 1.1) | 0.000 |
| Comorbidities |  |  |  |  |  |  |
| Acute myocardial infarction | 934 ( 5.3) | 2103 ( 0.6) | 0.277 | 42.3 ( 0.3) | 1012.7 ( 0.3) | 0.000 |
| Congestive heart failure | 1353 ( 7.6) | 3380 ( 1.0) | 0.330 | 39.8 ( 0.3) | 952.2 ( 0.3) | 0.000 |
| Peripheral vascular disease | 748 ( 4.2) | 2579 ( 0.8) | 0.222 | 28.2 ( 0.2) | 673.8 ( 0.2) | 0.000 |
| Cerebrovascular disease | 1451 ( 8.2) | 5189 ( 1.6) | 0.311 | 105.8 ( 0.8) | 2532.2 ( 0.8) | 0.000 |
| Alzheimer’s dementia | 461 ( 2.6) | 2294 ( 0.7) | 0.151 | 32.4 ( 0.3) | 775.6 ( 0.3) | 0.000 |
| Other dementia | 3152 ( 17.8) | 21520 ( 6.5) | 0.352 | 727.8 ( 5.7) | 17419.2 ( 5.7) | 0.000 |
| Mild liver disease | 578 ( 3.3) | 3363 ( 1.0) | 0.156 | 77.7 ( 0.6) | 1859.3 ( 0.6) | 0.000 |
| Diabetes without complications | 3888 ( 21.9) | 21896 ( 6.6) | 0.450 | 646.7 ( 5.0) | 15477.3 ( 5.0) | 0.000 |
| Diabetes with complications | 450 ( 2.5) | 1735 ( 0.5) | 0.165 | 12.8 ( 0.1) | 307.2 ( 0.1) | 0.000 |
| Renal disease | 939 ( 5.3) | 1904 ( 0.6) | 0.283 | 17.3 ( 0.1) | 413.7 ( 0.1) | 0.000 |
| Cancer | 2017 ( 11.4) | 10088 ( 3.0) | 0.327 | 263.3 ( 2.1) | 6301.7 ( 2.1) | 0.000 |
| Moderate to severe liver disease | 43 ( 0.2) | 66 ( 0.0) | 0.062 | 0.0 ( 0.0) | 0.0 ( 0.0) | 0.000 |
| Metastatic cancer | 208 ( 1.2) | 375 ( 0.1) | 0.133 | 3.6 ( 0.0) | 86.4 ( 0.0) | 0.000 |
| HIV/AIDS | 4 ( 0.0) | 21 ( 0.0) | 0.014 | 0.0 ( 0.0) | 0.0 ( 0.0) | 0.000 |
| RAAS inhibitors | 7861 ( 44.3) | 43193 ( 13.0) | 0.739 | 1548.8 ( 12.1) | 37066.2 ( 12.1) | 0.000 |
| Beta blockers | 5842 ( 32.9) | 22235 ( 6.7) | 0.698 | 742.2 ( 5.8) | 17762.8 ( 5.8) | 0.000 |
| Diuretics | 4586 ( 25.9) | 14387 ( 4.3) | 0.631 | 411.2 ( 3.2) | 9841.8 ( 3.2) | 0.000 |
| Immunosuppressant drugs | 560 ( 3.2) | 800 ( 0.2) | 0.227 | 15.8 ( 0.1) | 377.2 ( 0.1) | 0.000 |
| Systemic corticosteroids | 920 ( 5.2) | 617 ( 0.2) | 0.313 | 14.0 ( 0.1) | 334.0 ( 0.1) | 0.000 |
| Antineoplastic drugs | 337 ( 1.9) | 1241 ( 0.4) | 0.144 | 16.3 ( 0.1) | 390.7 ( 0.1) | 0.000 |
| Antiviral agents | 150 ( 0.8) | 215 ( 0.1) | 0.116 | 0.8 ( 0.0) | 18.2 ( 0.0) | 0.000 |
| Death (composite) | 1397 ( 7.9) | 5599 ( 1.7) | 0.293 | 169.7 ( 1.3) | 3681.6 ( 1.2) | 0.006 |
| Death 1 | 1022 ( 5.8) | 4366 ( 1.3) | 0.243 | 126.6 ( 1.0) | 2948.4 ( 1.0) | 0.001 |
| Death 2 | 920 ( 5.2) | 3698 ( 1.1) | 0.235 | 113.4 ( 0.9) | 2409.1 ( 0.8) | 0.006 |
| Death 3 | 1170 ( 6.6) | 4519 ( 1.4) | 0.270 | 139.5 ( 1.1) | 2925.9 ( 1.0) | 0.007 |
| Hospitalization 1 | 1830 ( 10.3) | 10905 ( 3.3) | 0.282 | 424.0 ( 3.3) | 8374.5 ( 2.7) | 0.023 |
| Hospitalization 2 | 4096 ( 23.1) | 26965 ( 8.1) | 0.422 | 1150.9 ( 9.0) | 21800.4 ( 7.1) | 0.053 |

**Table A18**. Sensitivity analysis: users (≥8 prescriptions) vs. non-users (counts/weighted counts, %)

|  | Before matching | | | After matching | | |
| --- | --- | --- | --- | --- | --- | --- |
| Characteristics | Treated | Control | d | Treated | Control | d |
| N | 18372 | 332389 |  | 11684 | 303530 |  |
| Age (5-year bins) |  |  |  |  |  |  |
| [16,21) | 22 ( 0.1) | 22054 ( 6.6) | -0.367 | 735.1 ( 6.3) | 19095.9 ( 6.3) | 0.000 |
| [21,26) | 99 ( 0.5) | 27711 ( 8.3) | -0.386 | 957.0 ( 8.2) | 24862.0 ( 8.2) | 0.000 |
| [26,31) | 160 ( 0.9) | 31607 ( 9.5) | -0.397 | 1116.9 ( 9.6) | 29014.1 ( 9.6) | 0.000 |
| [31,36) | 259 ( 1.4) | 34524 ( 10.4) | -0.388 | 1235.7 ( 10.6) | 32101.3 ( 10.6) | 0.000 |
| [36,41) | 486 ( 2.6) | 37047 ( 11.1) | -0.340 | 1337.4 ( 11.4) | 34743.6 ( 11.4) | 0.000 |
| [41,46) | 733 ( 4.0) | 35359 ( 10.6) | -0.257 | 1277.9 ( 10.9) | 33197.1 ( 10.9) | 0.000 |
| [46,51) | 1066 ( 5.8) | 30711 ( 9.2) | -0.131 | 1121.7 ( 9.6) | 29139.3 ( 9.6) | 0.000 |
| [51,56) | 1473 ( 8.0) | 28177 ( 8.5) | -0.017 | 1024.8 ( 8.8) | 26623.2 ( 8.8) | 0.000 |
| [56,61) | 1912 ( 10.4) | 25009 ( 7.5) | 0.101 | 898.8 ( 7.7) | 23349.2 ( 7.7) | 0.000 |
| [61,66) | 2301 ( 12.5) | 19809 ( 6.0) | 0.228 | 708.9 ( 6.1) | 18417.1 ( 6.1) | 0.000 |
| [66,71) | 2346 ( 12.8) | 14082 ( 4.2) | 0.310 | 483.6 ( 4.1) | 12564.4 ( 4.1) | 0.000 |
| [71,76) | 2031 ( 11.1) | 9476 ( 2.9) | 0.327 | 302.8 ( 2.6) | 7866.2 ( 2.6) | 0.000 |
| [76,81) | 1999 ( 10.9) | 6813 ( 2.0) | 0.365 | 208.2 ( 1.8) | 5409.8 ( 1.8) | 0.000 |
| [81,86) | 1919 ( 10.4) | 5529 ( 1.7) | 0.375 | 162.9 ( 1.4) | 4232.1 ( 1.4) | 0.000 |
| [86,91) | 1197 ( 6.5) | 3158 ( 1.0) | 0.297 | 86.6 ( 0.7) | 2248.4 ( 0.7) | 0.000 |
| [91,96) | 328 ( 1.8) | 1101 ( 0.3) | 0.142 | 23.6 ( 0.2) | 612.4 ( 0.2) | 0.000 |
| [96,101) | 39 ( 0.2) | 214 ( 0.1) | 0.040 | 2.1 ( 0.0) | 53.9 ( 0.0) | 0.000 |
| [101,106) | 2 ( 0.0) | 7 ( 0.0) | 0.011 | 0.0 ( 0.0) | 0.0 ( 0.0) | 0.000 |
| [106,111) | 0 ( 0.0) | 1 ( 0.0) | -0.002 | 0.0 ( 0.0) | 0.0 ( 0.0) | 0.000 |
| Sex |  |  |  |  |  |  |
| M | 7466 ( 40.6) | 162983 ( 49.0) | -0.169 | 5643.5 ( 48.3) | 146607.5 ( 48.3) | 0.000 |
| F | 10906 ( 59.4) | 169406 ( 51.0) | 0.169 | 6040.5 ( 51.7) | 156922.5 ( 51.7) | 0.000 |
| Vaccination status |  |  |  |  |  |  |
| 1/1, >90d | 0 ( 0.0) | 2 ( 0.0) | -0.003 | 0.0 ( 0.0) | 0.0 ( 0.0) | 0.000 |
| 1/1, <14d | 8 ( 0.0) | 47 ( 0.0) | 0.017 | 0.0 ( 0.0) | 0.0 ( 0.0) | 0.000 |
| 1/1, 14-90d | 1 ( 0.0) | 48 ( 0.0) | -0.009 | 0.0 ( 0.0) | 0.0 ( 0.0) | 0.000 |
| Not vaccinated | 16741 ( 91.1) | 322899 ( 97.1) | -0.258 | 11547.8 ( 98.8) | 299992.2 ( 98.8) | 0.000 |
| 1/2, >90d | 7 ( 0.0) | 35 ( 0.0) | 0.018 | 0.0 ( 0.0) | 0.0 ( 0.0) | 0.000 |
| 1/2, <14d | 422 ( 2.3) | 3549 ( 1.1) | 0.096 | 66.0 ( 0.6) | 1714.0 ( 0.6) | 0.000 |
| 1/2, 14-90d | 739 ( 4.0) | 3880 ( 1.2) | 0.180 | 63.8 ( 0.5) | 1656.2 ( 0.5) | 0.000 |
| 2/2, >90d | 127 ( 0.7) | 363 ( 0.1) | 0.092 | 0.6 ( 0.0) | 14.4 ( 0.0) | 0.000 |
| 2/2, <14d | 97 ( 0.5) | 420 ( 0.1) | 0.070 | 1.0 ( 0.0) | 27.0 ( 0.0) | 0.000 |
| 2/2, 14-90d | 230 ( 1.3) | 1146 ( 0.3) | 0.102 | 4.9 ( 0.0) | 126.1 ( 0.0) | 0.000 |
| Period |  |  |  |  |  |  |
| before 10.01.2021. | 11189 ( 60.9) | 192599 ( 57.9) | 0.060 | 7050.3 ( 60.3) | 183155.7 ( 60.3) | 0.000 |
| 10.01.-15.07.2021. | 6982 ( 38.0) | 132866 ( 40.0) | -0.040 | 4566.1 ( 39.1) | 118618.9 ( 39.1) | 0.000 |
| after 15.07.2021. | 201 ( 1.1) | 6924 ( 2.1) | -0.079 | 67.6 ( 0.6) | 1755.4 ( 0.6) | 0.000 |
| Comorbidities |  |  |  |  |  |  |
| Acute myocardial infarction | 1381 ( 7.5) | 2103 ( 0.6) | 0.354 | 44.6 ( 0.4) | 1159.4 ( 0.4) | 0.000 |
| Congestive heart failure | 2098 ( 11.4) | 3380 ( 1.0) | 0.441 | 46.6 ( 0.4) | 1209.4 ( 0.4) | 0.000 |
| Peripheral vascular disease | 1014 ( 5.5) | 2579 ( 0.8) | 0.274 | 25.1 ( 0.2) | 651.9 ( 0.2) | 0.000 |
| Cerebrovascular disease | 2121 ( 11.5) | 5189 ( 1.6) | 0.412 | 104.0 ( 0.9) | 2701.0 ( 0.9) | 0.000 |
| Alzheimer’s dementia | 813 ( 4.4) | 2294 ( 0.7) | 0.238 | 42.6 ( 0.4) | 1105.4 ( 0.4) | 0.000 |
| Other dementia | 3922 ( 21.3) | 21520 ( 6.5) | 0.440 | 653.6 ( 5.6) | 16979.4 ( 5.6) | 0.000 |
| Mild liver disease | 644 ( 3.5) | 3363 ( 1.0) | 0.168 | 65.6 ( 0.6) | 1703.4 ( 0.6) | 0.000 |
| Diabetes without complications | 5186 ( 28.2) | 21896 ( 6.6) | 0.595 | 622.5 ( 5.3) | 16171.5 ( 5.3) | 0.000 |
| Diabetes with complications | 745 ( 4.1) | 1735 ( 0.5) | 0.238 | 17.1 ( 0.1) | 443.9 ( 0.1) | 0.000 |
| Renal disease | 1424 ( 7.8) | 1904 ( 0.6) | 0.365 | 18.0 ( 0.2) | 468.0 ( 0.2) | 0.000 |
| Cancer | 2308 ( 12.6) | 10088 ( 3.0) | 0.361 | 233.0 ( 2.0) | 6052.0 ( 2.0) | 0.000 |
| Moderate to severe liver disease | 51 ( 0.3) | 66 ( 0.0) | 0.067 | 0.1 ( 0.0) | 1.9 ( 0.0) | 0.000 |
| Metastatic cancer | 220 ( 1.2) | 375 ( 0.1) | 0.135 | 2.7 ( 0.0) | 71.3 ( 0.0) | 0.000 |
| HIV/AIDS | 2 ( 0.0) | 21 ( 0.0) | 0.005 | 0.0 ( 0.0) | 0.0 ( 0.0) | 0.000 |
| RAAS inhibitors | 10065 ( 54.8) | 43193 ( 13.0) | 0.984 | 1491.2 ( 12.8) | 38739.8 ( 12.8) | 0.000 |
| Beta blockers | 7818 ( 42.6) | 22235 ( 6.7) | 0.916 | 724.2 ( 6.2) | 18812.8 ( 6.2) | 0.000 |
| Diuretics | 6604 ( 35.9) | 14387 ( 4.3) | 0.858 | 434.1 ( 3.7) | 11275.9 ( 3.7) | 0.000 |
| Immunosuppressant drugs | 691 ( 3.8) | 800 ( 0.2) | 0.253 | 11.4 ( 0.1) | 296.6 ( 0.1) | 0.000 |
| Systemic corticosteroids | 1076 ( 5.9) | 617 ( 0.2) | 0.336 | 12.1 ( 0.1) | 313.9 ( 0.1) | 0.000 |
| Antineoplastic drugs | 405 ( 2.2) | 1241 ( 0.4) | 0.163 | 15.8 ( 0.1) | 411.2 ( 0.1) | 0.000 |
| Antiviral agents | 187 ( 1.0) | 215 ( 0.1) | 0.130 | 0.7 ( 0.0) | 17.3 ( 0.0) | 0.000 |
| Death (composite) | 1889 ( 10.3) | 5599 ( 1.7) | 0.369 | 164.5 ( 1.4) | 3929.9 ( 1.3) | 0.005 |
| Death 1 | 1412 ( 7.7) | 4366 ( 1.3) | 0.311 | 130.7 ( 1.1) | 3134.8 ( 1.0) | 0.004 |
| Death 2 | 1215 ( 6.6) | 3698 ( 1.1) | 0.288 | 100.7 ( 0.9) | 2549.6 ( 0.8) | 0.001 |
| Death 3 | 1600 ( 8.7) | 4519 ( 1.4) | 0.341 | 134.8 ( 1.2) | 3129.3 ( 1.0) | 0.006 |
| Hospitalization 1 | 2348 ( 12.8) | 10905 ( 3.3) | 0.355 | 539.3 ( 4.6) | 8339.3 ( 2.7) | 0.070 |
| Hospitalization 2 | 5015 ( 27.3) | 26965 ( 8.1) | 0.519 | 1143.0 ( 9.8) | 21977.6 ( 7.2) | 0.069 |

**Table A19**. Sensitivity analysis: users (all) vs. possible (counts/weighted counts, %)

|  | Before matching | | | After matching | | |
| --- | --- | --- | --- | --- | --- | --- |
| Characteristics | Treated | Control | d | Treated | Control | d |
| N | 55098 | 18170 |  | 33272 | 16434 |  |
| Age (5-year bins) |  |  |  |  |  |  |
| [16,21) | 401 ( 0.7) | 540 ( 3.0) | -0.167 | 591.7 ( 1.8) | 292.3 ( 1.8) | 0.000 |
| [21,26) | 782 ( 1.4) | 759 ( 4.2) | -0.168 | 954.5 ( 2.9) | 471.5 ( 2.9) | 0.000 |
| [26,31) | 1137 ( 2.1) | 981 ( 5.4) | -0.177 | 1291.2 ( 3.9) | 637.8 ( 3.9) | 0.000 |
| [31,36) | 1703 ( 3.1) | 1142 ( 6.3) | -0.152 | 1737.0 ( 5.2) | 858.0 ( 5.2) | 0.000 |
| [36,41) | 2571 ( 4.7) | 1425 ( 7.8) | -0.131 | 2405.1 ( 7.2) | 1187.9 ( 7.2) | 0.000 |
| [41,46) | 3335 ( 6.1) | 1644 ( 9.0) | -0.114 | 2923.2 ( 8.8) | 1443.8 ( 8.8) | 0.000 |
| [46,51) | 4243 ( 7.7) | 1775 ( 9.8) | -0.073 | 3412.5 ( 10.3) | 1685.5 ( 10.3) | 0.000 |
| [51,56) | 5301 ( 9.6) | 1833 ( 10.1) | -0.016 | 3823.5 ( 11.5) | 1888.5 ( 11.5) | 0.000 |
| [56,61) | 6107 ( 11.1) | 1886 ( 10.4) | 0.023 | 3982.1 ( 12.0) | 1966.9 ( 12.0) | 0.000 |
| [61,66) | 6560 ( 11.9) | 1664 ( 9.2) | 0.090 | 3707.0 ( 11.1) | 1831.0 ( 11.1) | 0.000 |
| [66,71) | 6134 ( 11.1) | 1332 ( 7.3) | 0.132 | 2824.1 ( 8.5) | 1394.9 ( 8.5) | 0.000 |
| [71,76) | 5050 ( 9.2) | 1010 ( 5.6) | 0.138 | 2016.8 ( 6.1) | 996.2 ( 6.1) | 0.000 |
| [76,81) | 4480 ( 8.1) | 831 ( 4.6) | 0.146 | 1446.5 ( 4.3) | 714.5 ( 4.3) | 0.000 |
| [81,86) | 4104 ( 7.4) | 755 ( 4.2) | 0.141 | 1300.6 ( 3.9) | 642.4 ( 3.9) | 0.000 |
| [86,91) | 2400 ( 4.4) | 430 ( 2.4) | 0.111 | 684.1 ( 2.1) | 337.9 ( 2.1) | 0.000 |
| [91,96) | 693 ( 1.3) | 135 ( 0.7) | 0.052 | 158.0 ( 0.5) | 78.0 ( 0.5) | 0.000 |
| [96,101) | 95 ( 0.2) | 28 ( 0.2) | 0.005 | 14.1 ( 0.0) | 6.9 ( 0.0) | 0.000 |
| [101,106) | 2 ( 0.0) | 0 ( 0.0) | 0.009 | 0.0 ( 0.0) | 0.0 ( 0.0) | 0.000 |
| [106,111) | 0 ( 0.0) | 0 ( 0.0) | 0.000 | 0.0 ( 0.0) | 0.0 ( 0.0) | 0.000 |
| Sex |  |  |  |  |  |  |
| M | 22645 ( 41.1) | 7582 ( 41.7) | -0.013 | 12892.8 ( 38.7) | 6368.2 ( 38.7) | 0.000 |
| F | 32453 ( 58.9) | 10588 ( 58.3) | 0.013 | 20379.2 ( 61.3) | 10065.8 ( 61.3) | 0.000 |
| Vaccination status |  |  |  |  |  |  |
| 1/1, >90d | 2 ( 0.0) | 0 ( 0.0) | 0.009 | 0.0 ( 0.0) | 0.0 ( 0.0) | 0.000 |
| 1/1, <14d | 12 ( 0.0) | 1 ( 0.0) | 0.014 | 0.0 ( 0.0) | 0.0 ( 0.0) | 0.000 |
| 1/1, 14-90d | 5 ( 0.0) | 0 ( 0.0) | 0.013 | 0.0 ( 0.0) | 0.0 ( 0.0) | 0.000 |
| Not vaccinated | 51111 ( 92.8) | 17347 ( 95.5) | -0.115 | 32515.6 ( 97.7) | 16060.4 ( 97.7) | 0.000 |
| 1/2, >90d | 22 ( 0.0) | 7 ( 0.0) | 0.001 | 0.0 ( 0.0) | 0.0 ( 0.0) | 0.000 |
| 1/2, <14d | 1121 ( 2.0) | 295 ( 1.6) | 0.031 | 315.9 ( 0.9) | 156.1 ( 0.9) | 0.000 |
| 1/2, 14-90d | 1791 ( 3.3) | 355 ( 2.0) | 0.082 | 399.6 ( 1.2) | 197.4 ( 1.2) | 0.000 |
| 2/2, >90d | 239 ( 0.4) | 30 ( 0.2) | 0.049 | 4.0 ( 0.0) | 2.0 ( 0.0) | 0.000 |
| 2/2, <14d | 242 ( 0.4) | 44 ( 0.2) | 0.034 | 9.4 ( 0.0) | 4.6 ( 0.0) | 0.000 |
| 2/2, 14-90d | 553 ( 1.0) | 91 ( 0.5) | 0.058 | 27.4 ( 0.1) | 13.6 ( 0.1) | 0.000 |
| Period |  |  |  |  |  |  |
| before 10.01.2021. | 32317 ( 58.7) | 10566 ( 58.2) | 0.010 | 20363.1 ( 61.2) | 10057.9 ( 61.2) | 0.000 |
| 10.01.-15.07.2021. | 22102 ( 40.1) | 7257 ( 39.9) | 0.004 | 12637.8 ( 38.0) | 6242.2 ( 38.0) | 0.000 |
| after 15.07.2021. | 679 ( 1.2) | 347 ( 1.9) | -0.054 | 271.1 ( 0.8) | 133.9 ( 0.8) | 0.000 |
| Comorbidities |  |  |  |  |  |  |
| Acute myocardial infarction | 2830 ( 5.1) | 198 ( 1.1) | 0.235 | 254.4 ( 0.8) | 125.6 ( 0.8) | 0.000 |
| Congestive heart failure | 4234 ( 7.7) | 462 ( 2.5) | 0.235 | 309.9 ( 0.9) | 153.1 ( 0.9) | 0.000 |
| Peripheral vascular disease | 2238 ( 4.1) | 301 ( 1.7) | 0.145 | 184.7 ( 0.6) | 91.3 ( 0.6) | 0.000 |
| Cerebrovascular disease | 4552 ( 8.3) | 604 ( 3.3) | 0.213 | 742.3 ( 2.2) | 366.7 ( 2.2) | 0.000 |
| Alzheimer’s dementia | 1540 ( 2.8) | 249 ( 1.4) | 0.100 | 209.5 ( 0.6) | 103.5 ( 0.6) | 0.000 |
| Other dementia | 9700 ( 17.6) | 1979 ( 10.9) | 0.193 | 3488.1 ( 10.5) | 1722.9 ( 10.5) | 0.000 |
| Mild liver disease | 2849 ( 5.2) | 337 ( 1.9) | 0.181 | 634.6 ( 1.9) | 313.4 ( 1.9) | 0.000 |
| Diabetes without complications | 1724 ( 3.1) | 340 ( 1.9) | 0.081 | 403.0 ( 1.2) | 199.0 ( 1.2) | 0.000 |
| Diabetes with complications | 11803 ( 21.4) | 2449 ( 13.5) | 0.210 | 3901.8 ( 11.7) | 1927.2 ( 11.7) | 0.000 |
| Renal disease | 1499 ( 2.7) | 202 ( 1.1) | 0.118 | 91.0 ( 0.3) | 45.0 ( 0.3) | 0.000 |
| Cancer | 2805 ( 5.1) | 252 ( 1.4) | 0.210 | 119.8 ( 0.4) | 59.2 ( 0.4) | 0.000 |
| Moderate to severe liver disease | 6173 ( 11.2) | 971 ( 5.3) | 0.214 | 1419.7 ( 4.3) | 701.3 ( 4.3) | 0.000 |
| Metastatic cancer | 127 ( 0.2) | 7 ( 0.0) | 0.052 | 0.0 ( 0.0) | 0.0 ( 0.0) | 0.000 |
| HIV/AIDS | 611 ( 1.1) | 52 ( 0.3) | 0.099 | 23.4 ( 0.1) | 11.6 ( 0.1) | 0.000 |
| RAAS inhibitors | 8 ( 0.0) | 1 ( 0.0) | 0.009 | 0.0 ( 0.0) | 0.0 ( 0.0) | 0.000 |
| Beta blockers | 23376 ( 42.4) | 4841 ( 26.6) | 0.337 | 10210.0 ( 30.7) | 5043.0 ( 30.7) | 0.000 |
| Diuretics | 16969 ( 30.8) | 2450 ( 13.5) | 0.426 | 5316.2 ( 16.0) | 2625.8 ( 16.0) | 0.000 |
| Immunosuppressant drugs | 13757 ( 25.0) | 1872 ( 10.3) | 0.392 | 3547.0 ( 10.7) | 1752.0 ( 10.7) | 0.000 |
| Systemic corticosteroids | 1454 ( 2.6) | 75 ( 0.4) | 0.182 | 77.6 ( 0.2) | 38.4 ( 0.2) | 0.000 |
| Antineoplastic drugs | 2535 ( 4.6) | 72 ( 0.4) | 0.272 | 81.0 ( 0.2) | 40.0 ( 0.2) | 0.000 |
| Antiviral agents | 975 ( 1.8) | 118 ( 0.6) | 0.103 | 100.4 ( 0.3) | 49.6 ( 0.3) | 0.000 |
| Comorbidities | 417 ( 0.8) | 16 ( 0.1) | 0.103 | 0.0 ( 0.0) | 0.0 ( 0.0) | 0.000 |
| Death (composite) | 4356 ( 7.9) | 803 ( 4.4) | 0.145 | 1082.8 ( 3.3) | 609.8 ( 3.7) | -0.019 |
| Death 1 | 3195 ( 5.8) | 585 ( 3.2) | 0.125 | 854.1 ( 2.6) | 458.4 ( 2.8) | -0.011 |
| Death 2 | 2839 ( 5.2) | 519 ( 2.9) | 0.117 | 708.2 ( 2.1) | 395.1 ( 2.4) | -0.014 |
| Death 3 | 3670 ( 6.7) | 666 ( 3.7) | 0.136 | 885.2 ( 2.7) | 498.2 ( 3.0) | -0.017 |
| Hospitalization 1 | 5824 ( 10.6) | 1160 ( 6.4) | 0.151 | 1943.7 ( 5.8) | 910.2 ( 5.5) | 0.011 |
| Hospitalization 2 | 12735 ( 23.1) | 2680 ( 14.7) | 0.215 | 4821.5 ( 14.5) | 2280.9 ( 13.9) | 0.016 |

**Table A20**. Sensitivity analysis: users (1-3 prescriptions) vs. possible (counts/weighted counts, %)

|  | Before matching | | | After matching | | |
| --- | --- | --- | --- | --- | --- | --- |
| Characteristics | Treated | Control | d | Treated | Control | d |
| N | 18996 | 18170 |  | 14205 | 15405 |  |
| Age (5-year bins) |  |  |  |  |  |  |
| [16,21) | 305 ( 1.6) | 540 ( 3.0) | -0.091 | 383.3 ( 2.7) | 415.7 ( 2.7) | 0.000 |
| [21,26) | 515 ( 2.7) | 759 ( 4.2) | -0.080 | 580.5 ( 4.1) | 629.5 ( 4.1) | 0.000 |
| [26,31) | 726 ( 3.8) | 981 ( 5.4) | -0.075 | 767.1 ( 5.4) | 831.9 ( 5.4) | 0.000 |
| [31,36) | 1002 ( 5.3) | 1142 ( 6.3) | -0.043 | 969.1 ( 6.8) | 1050.9 ( 6.8) | 0.000 |
| [36,41) | 1378 ( 7.3) | 1425 ( 7.8) | -0.022 | 1261.7 ( 8.9) | 1368.3 ( 8.9) | 0.000 |
| [41,46) | 1596 ( 8.4) | 1644 ( 9.0) | -0.023 | 1427.7 ( 10.1) | 1548.3 ( 10.1) | 0.000 |
| [46,51) | 1825 ( 9.6) | 1775 ( 9.8) | -0.005 | 1559.1 ( 11.0) | 1690.9 ( 11.0) | 0.000 |
| [51,56) | 2101 ( 11.1) | 1833 ( 10.1) | 0.032 | 1646.0 ( 11.6) | 1785.0 ( 11.6) | 0.000 |
| [56,61) | 2099 ( 11.0) | 1886 ( 10.4) | 0.022 | 1586.5 ( 11.2) | 1720.5 ( 11.2) | 0.000 |
| [61,66) | 2054 ( 10.8) | 1664 ( 9.2) | 0.055 | 1383.1 ( 9.7) | 1499.9 ( 9.7) | 0.000 |
| [66,71) | 1680 ( 8.8) | 1332 ( 7.3) | 0.056 | 975.8 ( 6.9) | 1058.2 ( 6.9) | 0.000 |
| [71,76) | 1310 ( 6.9) | 1010 ( 5.6) | 0.055 | 655.8 ( 4.6) | 711.2 ( 4.6) | 0.000 |
| [76,81) | 1012 ( 5.3) | 831 ( 4.6) | 0.035 | 433.7 ( 3.1) | 470.3 ( 3.1) | 0.000 |
| [81,86) | 826 ( 4.3) | 755 ( 4.2) | 0.010 | 360.8 ( 2.5) | 391.2 ( 2.5) | 0.000 |
| [86,91) | 433 ( 2.3) | 430 ( 2.4) | -0.006 | 179.4 ( 1.3) | 194.6 ( 1.3) | 0.000 |
| [91,96) | 118 ( 0.6) | 135 ( 0.7) | -0.015 | 34.5 ( 0.2) | 37.5 ( 0.2) | 0.000 |
| [96,101) | 16 ( 0.1) | 28 ( 0.2) | -0.020 | 1.0 ( 0.0) | 1.0 ( 0.0) | 0.000 |
| [101,106) | 0 ( 0.0) | 0 ( 0.0) | 0.000 | 0.0 ( 0.0) | 0.0 ( 0.0) | 0.000 |
| [106,111) | 0 ( 0.0) | 0 ( 0.0) | 0.000 | 0.0 ( 0.0) | 0.0 ( 0.0) | 0.000 |
| Sex |  |  |  |  |  |  |
| M | 7707 ( 40.6) | 7582 ( 41.7) | -0.024 | 5553.9 ( 39.1) | 6023.1 ( 39.1) | 0.000 |
| F | 11289 ( 59.4) | 10588 ( 58.3) | 0.024 | 8651.1 ( 60.9) | 9381.9 ( 60.9) | 0.000 |
| Vaccination status |  |  |  |  |  |  |
| 1/1, >90d | 1 ( 0.0) | 0 ( 0.0) | 0.010 | 0.0 ( 0.0) | 0.0 ( 0.0) | 0.000 |
| 1/1, <14d | 2 ( 0.0) | 1 ( 0.0) | 0.006 | 0.0 ( 0.0) | 0.0 ( 0.0) | 0.000 |
| 1/1, 14-90d | 2 ( 0.0) | 0 ( 0.0) | 0.015 | 0.0 ( 0.0) | 0.0 ( 0.0) | 0.000 |
| Not vaccinated | 18000 ( 94.8) | 17347 ( 95.5) | -0.033 | 13918.6 ( 98.0) | 15094.4 ( 98.0) | 0.000 |
| 1/2, >90d | 7 ( 0.0) | 7 ( 0.0) | -0.001 | 0.0 ( 0.0) | 0.0 ( 0.0) | 0.000 |
| 1/2, <14d | 332 ( 1.7) | 295 ( 1.6) | 0.010 | 131.4 ( 0.9) | 142.6 ( 0.9) | 0.000 |
| 1/2, 14-90d | 431 ( 2.3) | 355 ( 2.0) | 0.022 | 139.6 ( 1.0) | 151.4 ( 1.0) | 0.000 |
| 2/2, >90d | 36 ( 0.2) | 30 ( 0.2) | 0.006 | 0.0 ( 0.0) | 0.0 ( 0.0) | 0.000 |
| 2/2, <14d | 56 ( 0.3) | 44 ( 0.2) | 0.010 | 3.4 ( 0.0) | 3.6 ( 0.0) | 0.000 |
| 2/2, 14-90d | 129 ( 0.7) | 91 ( 0.5) | 0.023 | 12.0 ( 0.1) | 13.0 ( 0.1) | 0.000 |
| Period |  |  |  |  |  |  |
| before 10.01.2021. | 10627 ( 55.9) | 10566 ( 58.2) | -0.045 | 8404.5 ( 59.2) | 9114.5 ( 59.2) | 0.000 |
| 10.01.-15.07.2021. | 8105 ( 42.7) | 7257 ( 39.9) | 0.055 | 5644.1 ( 39.7) | 6120.9 ( 39.7) | 0.000 |
| after 15.07.2021. | 264 ( 1.4) | 347 ( 1.9) | -0.041 | 156.4 ( 1.1) | 169.6 ( 1.1) | 0.000 |
| Comorbidities |  |  |  |  |  |  |
| Acute myocardial infarction | 515 ( 2.7) | 198 ( 1.1) | 0.119 | 54.2 ( 0.4) | 58.8 ( 0.4) | 0.000 |
| Congestive heart failure | 783 ( 4.1) | 462 ( 2.5) | 0.088 | 70.5 ( 0.5) | 76.5 ( 0.5) | 0.000 |
| Peripheral vascular disease | 476 ( 2.5) | 301 ( 1.7) | 0.060 | 56.1 ( 0.4) | 60.9 ( 0.4) | 0.000 |
| Cerebrovascular disease | 980 ( 5.2) | 604 ( 3.3) | 0.091 | 196.7 ( 1.4) | 213.3 ( 1.4) | 0.000 |
| Alzheimer’s dementia | 266 ( 1.4) | 249 ( 1.4) | 0.003 | 48.9 ( 0.3) | 53.1 ( 0.3) | 0.000 |
| Other dementia | 2626 ( 13.8) | 1979 ( 10.9) | 0.089 | 1282.8 ( 9.0) | 1391.2 ( 9.0) | 0.000 |
| Mild liver disease | 502 ( 2.6) | 340 ( 1.9) | 0.052 | 160.7 ( 1.1) | 174.3 ( 1.1) | 0.000 |
| Diabetes without complications | 2729 ( 14.4) | 2449 ( 13.5) | 0.026 | 1266.5 ( 8.9) | 1373.5 ( 8.9) | 0.000 |
| Diabetes with complications | 304 ( 1.6) | 202 ( 1.1) | 0.042 | 21.6 ( 0.2) | 23.4 ( 0.2) | 0.000 |
| Renal disease | 442 ( 2.3) | 252 ( 1.4) | 0.070 | 22.5 ( 0.2) | 24.5 ( 0.2) | 0.000 |
| Cancer | 1848 ( 9.7) | 971 ( 5.3) | 0.167 | 542.1 ( 3.8) | 587.9 ( 3.8) | 0.000 |
| Moderate to severe liver disease | 33 ( 0.2) | 7 ( 0.0) | 0.042 | 0.0 ( 0.0) | 0.0 ( 0.0) | 0.000 |
| Metastatic cancer | 183 ( 1.0) | 52 ( 0.3) | 0.086 | 9.6 ( 0.1) | 10.4 ( 0.1) | 0.000 |
| HIV/AIDS | 2 ( 0.0) | 1 ( 0.0) | 0.006 | 0.0 ( 0.0) | 0.0 ( 0.0) | 0.000 |
| RAAS inhibitors | 5450 ( 28.7) | 4841 ( 26.6) | 0.046 | 3196.5 ( 22.5) | 3466.5 ( 22.5) | 0.000 |
| Beta blockers | 3309 ( 17.4) | 2450 ( 13.5) | 0.109 | 1435.9 ( 10.1) | 1557.1 ( 10.1) | 0.000 |
| Diuretics | 2567 ( 13.5) | 1872 ( 10.3) | 0.099 | 893.7 ( 6.3) | 969.3 ( 6.3) | 0.000 |
| Immunosuppressant drugs | 203 ( 1.1) | 75 ( 0.4) | 0.077 | 21.6 ( 0.2) | 23.4 ( 0.2) | 0.000 |
| Systemic corticosteroids | 539 ( 2.8) | 72 ( 0.4) | 0.194 | 26.9 ( 0.2) | 29.1 ( 0.2) | 0.000 |
| Antineoplastic drugs | 233 ( 1.2) | 118 ( 0.6) | 0.060 | 33.1 ( 0.2) | 35.9 ( 0.2) | 0.000 |
| Antiviral agents | 80 ( 0.4) | 16 ( 0.1) | 0.066 | 0.0 ( 0.0) | 0.0 ( 0.0) | 0.000 |
| Death (composite) | 1070 ( 5.6) | 803 ( 4.4) | 0.056 | 363.2 ( 2.6) | 386.9 ( 2.5) | 0.002 |
| Death 1 | 761 ( 4.0) | 585 ( 3.2) | 0.042 | 288.2 ( 2.0) | 297.0 ( 1.9) | 0.005 |
| Death 2 | 704 ( 3.7) | 519 ( 2.9) | 0.048 | 244.2 ( 1.7) | 256.4 ( 1.7) | 0.003 |
| Death 3 | 900 ( 4.7) | 666 ( 3.7) | 0.053 | 292.9 ( 2.1) | 317.5 ( 2.1) | 0.000 |
| Hospitalization 1 | 1646 ( 8.7) | 1160 ( 6.4) | 0.087 | 803.4 ( 5.7) | 701.2 ( 4.6) | 0.042 |
| Hospitalization 2 | 3624 ( 19.1) | 2680 ( 14.7) | 0.116 | 1912.6 ( 13.5) | 1801.1 ( 11.7) | 0.047 |

**Table A21**. Sensitivity analysis: users (4-7 prescriptions) vs. possible (counts/weighted counts, %)

|  | Before matching | | | After matching | | |
| --- | --- | --- | --- | --- | --- | --- |
| Characteristics | Treated | Control | d | Treated | Control | d |
| N | 17730 | 18170 |  | 10315 | 15414 |  |
| Age (5-year bins) |  |  |  |  |  |  |
| [16,21) | 74 ( 0.4) | 540 ( 3.0) | -0.199 | 224.5 ( 2.2) | 335.5 ( 2.2) | 0.000 |
| [21,26) | 168 ( 0.9) | 759 ( 4.2) | -0.205 | 334.0 ( 3.2) | 499.0 ( 3.2) | 0.000 |
| [26,31) | 251 ( 1.4) | 981 ( 5.4) | -0.221 | 453.4 ( 4.4) | 677.6 ( 4.4) | 0.000 |
| [31,36) | 442 ( 2.5) | 1142 ( 6.3) | -0.186 | 585.7 ( 5.7) | 875.3 ( 5.7) | 0.000 |
| [36,41) | 707 ( 4.0) | 1425 ( 7.8) | -0.164 | 771.3 ( 7.5) | 1152.7 ( 7.5) | 0.000 |
| [41,46) | 1006 ( 5.7) | 1644 ( 9.0) | -0.129 | 937.3 ( 9.1) | 1400.7 ( 9.1) | 0.000 |
| [46,51) | 1352 ( 7.6) | 1775 ( 9.8) | -0.076 | 1086.5 ( 10.5) | 1623.5 ( 10.5) | 0.000 |
| [51,56) | 1727 ( 9.7) | 1833 ( 10.1) | -0.012 | 1172.7 ( 11.4) | 1752.3 ( 11.4) | 0.000 |
| [56,61) | 2096 ( 11.8) | 1886 ( 10.4) | 0.046 | 1228.8 ( 11.9) | 1836.2 ( 11.9) | 0.000 |
| [61,66) | 2205 ( 12.4) | 1664 ( 9.2) | 0.106 | 1083.7 ( 10.5) | 1619.3 ( 10.5) | 0.000 |
| [66,71) | 2108 ( 11.9) | 1332 ( 7.3) | 0.155 | 840.7 ( 8.2) | 1256.3 ( 8.2) | 0.000 |
| [71,76) | 1709 ( 9.6) | 1010 ( 5.6) | 0.154 | 578.9 ( 5.6) | 865.1 ( 5.6) | 0.000 |
| [76,81) | 1469 ( 8.3) | 831 ( 4.6) | 0.152 | 410.5 ( 4.0) | 613.5 ( 4.0) | 0.000 |
| [81,86) | 1359 ( 7.7) | 755 ( 4.2) | 0.149 | 367.2 ( 3.6) | 548.8 ( 3.6) | 0.000 |
| [86,91) | 770 ( 4.3) | 430 ( 2.4) | 0.110 | 191.6 ( 1.9) | 286.4 ( 1.9) | 0.000 |
| [91,96) | 247 ( 1.4) | 135 ( 0.7) | 0.063 | 42.5 ( 0.4) | 63.5 ( 0.4) | 0.000 |
| [96,101) | 40 ( 0.2) | 28 ( 0.2) | 0.016 | 5.6 ( 0.1) | 8.4 ( 0.1) | 0.000 |
| [101,106) | 0 ( 0.0) | 0 ( 0.0) | 0.000 | 0.0 ( 0.0) | 0.0 ( 0.0) | 0.000 |
| [106,111) | 0 ( 0.0) | 0 ( 0.0) | 0.000 | 0.0 ( 0.0) | 0.0 ( 0.0) | 0.000 |
| Sex |  |  |  |  |  |  |
| M | 7472 ( 42.1) | 7582 ( 41.7) | 0.008 | 4092.1 ( 39.7) | 6114.9 ( 39.7) | 0.000 |
| F | 10258 ( 57.9) | 10588 ( 58.3) | -0.008 | 6222.9 ( 60.3) | 9299.1 ( 60.3) | 0.000 |
| Vaccination status |  |  |  |  |  |  |
| 1/1, >90d | 1 ( 0.0) | 0 ( 0.0) | 0.011 | 0.0 ( 0.0) | 0.0 ( 0.0) | 0.000 |
| 1/1, <14d | 2 ( 0.0) | 1 ( 0.0) | 0.006 | 0.0 ( 0.0) | 0.0 ( 0.0) | 0.000 |
| 1/1, 14-90d | 2 ( 0.0) | 0 ( 0.0) | 0.015 | 0.0 ( 0.0) | 0.0 ( 0.0) | 0.000 |
| Not vaccinated | 16370 ( 92.3) | 17347 ( 95.5) | -0.132 | 10105.3 ( 98.0) | 15100.7 ( 98.0) | 0.000 |
| 1/2, >90d | 8 ( 0.0) | 7 ( 0.0) | 0.003 | 0.0 ( 0.0) | 0.0 ( 0.0) | 0.000 |
| 1/2, <14d | 367 ( 2.1) | 295 ( 1.6) | 0.033 | 87.4 ( 0.8) | 130.6 ( 0.8) | 0.000 |
| 1/2, 14-90d | 621 ( 3.5) | 355 ( 2.0) | 0.095 | 112.3 ( 1.1) | 167.7 ( 1.1) | 0.000 |
| 2/2, >90d | 76 ( 0.4) | 30 ( 0.2) | 0.048 | 2.0 ( 0.0) | 3.0 ( 0.0) | 0.000 |
| 2/2, <14d | 89 ( 0.5) | 44 ( 0.2) | 0.043 | 2.8 ( 0.0) | 4.2 ( 0.0) | 0.000 |
| 2/2, 14-90d | 194 ( 1.1) | 91 ( 0.5) | 0.067 | 5.2 ( 0.1) | 7.8 ( 0.1) | 0.000 |
| Period |  |  |  |  |  |  |
| before 10.01.2021. | 10501 ( 59.2) | 10566 ( 58.2) | 0.022 | 6370.9 ( 61.8) | 9520.1 ( 61.8) | 0.000 |
| 10.01.-15.07.2021. | 7015 ( 39.6) | 7257 ( 39.9) | -0.008 | 3868.4 ( 37.5) | 5780.6 ( 37.5) | 0.000 |
| after 15.07.2021. | 214 ( 1.2) | 347 ( 1.9) | -0.057 | 75.8 ( 0.7) | 113.2 ( 0.7) | 0.000 |
| Comorbidities |  |  |  |  |  |  |
| Acute myocardial infarction | 934 ( 5.3) | 198 ( 1.1) | 0.240 | 60.5 ( 0.6) | 90.5 ( 0.6) | 0.000 |
| Congestive heart failure | 1353 ( 7.6) | 462 ( 2.5) | 0.233 | 71.4 ( 0.7) | 106.6 ( 0.7) | 0.000 |
| Peripheral vascular disease | 748 ( 4.2) | 301 ( 1.7) | 0.152 | 38.1 ( 0.4) | 56.9 ( 0.4) | 0.000 |
| Cerebrovascular disease | 1451 ( 8.2) | 604 ( 3.3) | 0.210 | 192.4 ( 1.9) | 287.6 ( 1.9) | 0.000 |
| Alzheimer’s dementia | 461 ( 2.6) | 249 ( 1.4) | 0.088 | 47.3 ( 0.5) | 70.7 ( 0.5) | 0.000 |
| Other dementia | 3152 ( 17.8) | 1979 ( 10.9) | 0.197 | 991.4 ( 9.6) | 1481.6 ( 9.6) | 0.000 |
| Mild liver disease | 578 ( 3.3) | 340 ( 1.9) | 0.088 | 106.2 ( 1.0) | 158.8 ( 1.0) | 0.000 |
| Diabetes without complications | 3888 ( 21.9) | 2449 ( 13.5) | 0.223 | 1096.5 ( 10.6) | 1638.5 ( 10.6) | 0.000 |
| Diabetes with complications | 450 ( 2.5) | 202 ( 1.1) | 0.107 | 17.2 ( 0.2) | 25.8 ( 0.2) | 0.000 |
| Renal disease | 939 ( 5.3) | 252 ( 1.4) | 0.219 | 24.5 ( 0.2) | 36.5 ( 0.2) | 0.000 |
| Cancer | 2017 ( 11.4) | 971 ( 5.3) | 0.219 | 362.4 ( 3.5) | 541.6 ( 3.5) | 0.000 |
| Moderate to severe liver disease | 43 ( 0.2) | 7 ( 0.0) | 0.054 | 0.0 ( 0.0) | 0.0 ( 0.0) | 0.000 |
| Metastatic cancer | 208 ( 1.2) | 52 ( 0.3) | 0.104 | 2.8 ( 0.0) | 4.2 ( 0.0) | 0.000 |
| HIV/AIDS | 4 ( 0.0) | 1 ( 0.0) | 0.014 | 0.0 ( 0.0) | 0.0 ( 0.0) | 0.000 |
| RAAS inhibitors | 7861 ( 44.3) | 4841 ( 26.6) | 0.376 | 2970.7 ( 28.8) | 4439.3 ( 28.8) | 0.000 |
| Beta blockers | 5842 ( 32.9) | 2450 ( 13.5) | 0.474 | 1503.0 ( 14.6) | 2246.0 ( 14.6) | 0.000 |
| Diuretics | 4586 ( 25.9) | 1872 ( 10.3) | 0.413 | 975.0 ( 9.5) | 1457.0 ( 9.5) | 0.000 |
| Immunosuppressant drugs | 560 ( 3.2) | 75 ( 0.4) | 0.208 | 21.2 ( 0.2) | 31.8 ( 0.2) | 0.000 |
| Systemic corticosteroids | 920 ( 5.2) | 72 ( 0.4) | 0.294 | 22.5 ( 0.2) | 33.5 ( 0.2) | 0.000 |
| Antineoplastic drugs | 337 ( 1.9) | 118 ( 0.6) | 0.112 | 21.2 ( 0.2) | 31.8 ( 0.2) | 0.000 |
| Antiviral agents | 150 ( 0.8) | 16 ( 0.1) | 0.111 | 0.0 ( 0.0) | 0.0 ( 0.0) | 0.000 |
| Death (composite) | 1397 ( 7.9) | 803 ( 4.4) | 0.144 | 282.6 ( 2.7) | 519.7 ( 3.4) | -0.026 |
| Death 1 | 1022 ( 5.8) | 585 ( 3.2) | 0.123 | 216.0 ( 2.1) | 395.9 ( 2.6) | -0.023 |
| Death 2 | 920 ( 5.2) | 519 ( 2.9) | 0.119 | 179.8 ( 1.7) | 340.6 ( 2.2) | -0.024 |
| Death 3 | 1170 ( 6.6) | 666 ( 3.7) | 0.133 | 234.7 ( 2.3) | 421.2 ( 2.7) | -0.021 |
| Hospitalization 1 | 1830 ( 10.3) | 1160 ( 6.4) | 0.143 | 501.9 ( 4.9) | 812.1 ( 5.3) | -0.015 |
| Hospitalization 2 | 4096 ( 23.1) | 2680 ( 14.7) | 0.214 | 1322.5 ( 12.8) | 2031.5 ( 13.2) | -0.009 |

**Table A22**. Sensitivity analysis: users (≥8 prescriptions) vs. possible (counts/weighted counts, %)

|  | Before matching | | | | After matching | | |
| --- | --- | --- | --- | --- | --- | --- | --- |
| Characteristics | Treated | Control | d | Treated | | Control | d |
| N | 18372 | 18170 |  | 8752 | | 15337 |  |
| Age (5-year bins) |  |  |  |  | |  |  |
| [16,21) | 22 ( 0.1) | 540 ( 3.0) | -0.233 | 176.9 ( 2.0) | | 310.1 ( 2.0) | 0.000 |
| [21,26) | 99 ( 0.5) | 759 ( 4.2) | -0.242 | 273.2 ( 3.1) | | 478.8 ( 3.1) | 0.000 |
| [26,31) | 160 ( 0.9) | 981 ( 5.4) | -0.262 | 374.9 ( 4.3) | | 657.1 ( 4.3) | 0.000 |
| [31,36) | 259 ( 1.4) | 1142 ( 6.3) | -0.256 | 451.2 ( 5.2) | | 790.8 ( 5.2) | 0.000 |
| [36,41) | 486 ( 2.6) | 1425 ( 7.8) | -0.235 | 616.6 ( 7.0) | | 1080.4 ( 7.0) | 0.000 |
| [41,46) | 733 ( 4.0) | 1644 ( 9.0) | -0.206 | 743.0 ( 8.5) | | 1302.0 ( 8.5) | 0.000 |
| [46,51) | 1066 ( 5.8) | 1775 ( 9.8) | -0.148 | 860.7 ( 9.8) | | 1508.3 ( 9.8) | 0.000 |
| [51,56) | 1473 ( 8.0) | 1833 ( 10.1) | -0.072 | 942.8 ( 10.8) | | 1652.2 ( 10.8) | 0.000 |
| [56,61) | 1912 ( 10.4) | 1886 ( 10.4) | 0.001 | 1007.5 ( 11.5) | | 1765.5 ( 11.5) | 0.000 |
| [61,66) | 2301 ( 12.5) | 1664 ( 9.2) | 0.108 | 948.6 ( 10.8) | | 1662.4 ( 10.8) | 0.000 |
| [66,71) | 2346 ( 12.8) | 1332 ( 7.3) | 0.182 | 757.5 ( 8.7) | | 1327.5 ( 8.7) | 0.000 |
| [71,76) | 2031 ( 11.1) | 1010 ( 5.6) | 0.200 | 542.1 ( 6.2) | | 949.9 ( 6.2) | 0.000 |
| [76,81) | 1999 ( 10.9) | 831 ( 4.6) | 0.238 | 417.8 ( 4.8) | | 732.2 ( 4.8) | 0.000 |
| [81,86) | 1919 ( 10.4) | 755 ( 4.2) | 0.244 | 380.0 ( 4.3) | | 666.0 ( 4.3) | 0.000 |
| [86,91) | 1197 ( 6.5) | 430 ( 2.4) | 0.202 | 205.3 ( 2.3) | | 359.7 ( 2.3) | 0.000 |
| [91,96) | 328 ( 1.8) | 135 ( 0.7) | 0.093 | 50.9 ( 0.6) | | 89.1 ( 0.6) | 0.000 |
| [96,101) | 39 ( 0.2) | 28 ( 0.2) | 0.014 | 2.9 ( 0.0) | | 5.1 ( 0.0) | 0.000 |
| [101,106) | 2 ( 0.0) | 0 ( 0.0) | 0.015 | 0.0 ( 0.0) | | 0.0 ( 0.0) | 0.000 |
| [106,111) | 0 ( 0.0) | 0 ( 0.0) | 0.000 | 0.0 ( 0.0) | | 0.0 ( 0.0) | 0.000 |
| Sex |  |  |  |  | |  |  |
| M | 7466 ( 40.6) | 7582 ( 41.7) | -0.022 | 3395.9 ( 38.8) | | 5951.1 ( 38.8) | 0.000 |
| F | 10906 ( 59.4) | 10588 ( 58.3) | 0.022 | 5356.1 ( 61.2) | | 9385.9 ( 61.2) | 0.000 |
| Vaccination status |  |  |  |  | |  |  |
| 1/1, >90d | 0 ( 0.0) | 0 ( 0.0) | 0.000 | 0.0 ( 0.0) | | 0.0 ( 0.0) | 0.000 |
| 1/1, <14d | 8 ( 0.0) | 1 ( 0.0) | 0.024 | 0.0 ( 0.0) | | 0.0 ( 0.0) | 0.000 |
| 1/1, 14-90d | 1 ( 0.0) | 0 ( 0.0) | 0.010 | 0.0 ( 0.0) | | 0.0 ( 0.0) | 0.000 |
| Not vaccinated | 16741 ( 91.1) | 17347 ( 95.5) | -0.175 | 8590.3 ( 98.2) | | 15053.7 ( 98.2) | 0.000 |
| 1/2, >90d | 7 ( 0.0) | 7 ( 0.0) | 0.000 | 0.0 ( 0.0) | | 0.0 ( 0.0) | 0.000 |
| 1/2, <14d | 422 ( 2.3) | 295 ( 1.6) | 0.049 | 69.4 ( 0.8) | | 121.6 ( 0.8) | 0.000 |
| 1/2, 14-90d | 739 ( 4.0) | 355 ( 2.0) | 0.122 | 87.2 ( 1.0) | | 152.8 ( 1.0) | 0.000 |
| 2/2, >90d | 127 ( 0.7) | 30 ( 0.2) | 0.081 | 0.7 ( 0.0) | | 1.3 ( 0.0) | 0.000 |
| 2/2, <14d | 97 ( 0.5) | 44 ( 0.2) | 0.046 | 0.7 ( 0.0) | | 1.3 ( 0.0) | 0.000 |
| 2/2, 14-90d | 230 ( 1.3) | 91 ( 0.5) | 0.081 | 3.6 ( 0.0) | | 6.4 ( 0.0) | 0.000 |
| Period |  |  |  |  | |  |  |
| before 10.01.2021. | 11189 ( 60.9) | 10566 ( 58.2) | 0.056 | 5507.2 ( 62.9) | | 9650.8 ( 62.9) | 0.000 |
| 10.01.-15.07.2021. | 6982 ( 38.0) | 7257 ( 39.9) | -0.040 | 3205.6 ( 36.6) | | 5617.4 ( 36.6) | 0.000 |
| after 15.07.2021. | 201 ( 1.1) | 347 ( 1.9) | -0.067 | 39.2 ( 0.4) | | 68.8 ( 0.4) | 0.000 |
| Comorbidities |  |  |  |  | |  |  |
| Acute myocardial infarction | 1381 ( 7.5) | 198 ( 1.1) | 0.321 | 67.9 ( 0.8) | | 119.1 ( 0.8) | 0.000 |
| Congestive heart failure | 2098 ( 11.4) | 462 ( 2.5) | 0.354 | 78.8 ( 0.9) | | 138.2 ( 0.9) | 0.000 |
| Peripheral vascular disease | 1014 ( 5.5) | 301 ( 1.7) | 0.209 | 44.0 ( 0.5) | | 77.0 ( 0.5) | 0.000 |
| Cerebrovascular disease | 2121 ( 11.5) | 604 ( 3.3) | 0.317 | 198.0 ( 2.3) | | 347.0 ( 2.3) | 0.000 |
| Alzheimer’s dementia | 813 ( 4.4) | 249 ( 1.4) | 0.183 | 68.7 ( 0.8) | | 120.3 ( 0.8) | 0.000 |
| Other dementia | 3922 ( 21.3) | 1979 ( 10.9) | 0.287 | 891.2 ( 10.2) | | 1561.8 ( 10.2) | 0.000 |
| Mild liver disease | 644 ( 3.5) | 340 ( 1.9) | 0.101 | 85.7 ( 1.0) | | 150.3 ( 1.0) | 0.000 |
| Diabetes without complications | 5186 ( 28.2) | 2449 ( 13.5) | 0.369 | 1118.7 ( 12.8) | | 1960.3 ( 12.8) | 0.000 |
| Diabetes with complications | 745 ( 4.1) | 202 ( 1.1) | 0.186 | 24.3 ( 0.3) | | 42.7 ( 0.3) | 0.000 |
| Renal disease | 1424 ( 7.8) | 252 ( 1.4) | 0.308 | 32.3 ( 0.4) | | 56.7 ( 0.4) | 0.000 |
| Cancer | 2308 ( 12.6) | 971 ( 5.3) | 0.255 | 315.7 ( 3.6) | | 553.3 ( 3.6) | 0.000 |
| Moderate to severe liver disease | 51 ( 0.3) | 7 ( 0.0) | 0.060 | 0.0 ( 0.0) | | 0.0 ( 0.0) | 0.000 |
| Metastatic cancer | 220 ( 1.2) | 52 ( 0.3) | 0.106 | 4.0 ( 0.0) | | 7.0 ( 0.0) | 0.000 |
| HIV/AIDS | 2 ( 0.0) | 1 ( 0.0) | 0.006 | 0.0 ( 0.0) | | 0.0 ( 0.0) | 0.000 |
| RAAS inhibitors | 10065 ( 54.8) | 4841 ( 26.6) | 0.598 | 2894.6 ( 33.1) | | 5072.4 ( 33.1) | 0.000 |
| Beta blockers | 7818 ( 42.6) | 2450 ( 13.5) | 0.684 | 1511.8 ( 17.3) | | 2649.2 ( 17.3) | 0.000 |
| Diuretics | 6604 ( 35.9) | 1872 ( 10.3) | 0.638 | 1022.0 ( 11.7) | | 1791.0 ( 11.7) | 0.000 |
| Immunosuppressant drugs | 691 ( 3.8) | 75 ( 0.4) | 0.236 | 17.1 ( 0.2) | | 29.9 ( 0.2) | 0.000 |
| Systemic corticosteroids | 1076 ( 5.9) | 72 ( 0.4) | 0.318 | 15.3 ( 0.2) | | 26.7 ( 0.2) | 0.000 |
| Antineoplastic drugs | 405 ( 2.2) | 118 ( 0.6) | 0.131 | 24.0 ( 0.3) | | 42.0 ( 0.3) | 0.000 |
| Antiviral agents | 187 ( 1.0) | 16 ( 0.1) | 0.126 | 0.0 ( 0.0) | | 0.0 ( 0.0) | 0.000 |
| Death (composite) | 1889 ( 10.3) | 803 ( 4.4) | 0.226 | 294.1 ( 3.4) | | 626.1 ( 4.1) | -0.028 |
| Death 1 | 1412 ( 7.7) | 585 ( 3.2) | 0.198 | 235.3 ( 2.7) | | 467.1 ( 3.0) | -0.016 |
| Death 2 | 1215 ( 6.6) | 519 ( 2.9) | 0.178 | 188.1 ( 2.1) | | 401.7 ( 2.6) | -0.022 |
| Death 3 | 1600 ( 8.7) | 666 ( 3.7) | 0.210 | 238.5 ( 2.7) | | 509.0 ( 3.3) | -0.025 |
| Hospitalization 1 | 2348 ( 12.8) | 1160 ( 6.4) | 0.219 | 516.4 ( 5.9) | | 866.4 ( 5.6) | 0.009 |
| Hospitalization 2 | 5015 ( 27.3) | 2680 ( 14.7) | 0.312 | 1265.1 ( 14.5) | | 2153.2 ( 14.0) | 0.010 |

**Table A23**. Sensitivity analysis to account for potential misclassification of possible users: users (all) (treated) vs. unclassified patients (control) (count/weighted count, %)

|  | Before matching | | | After matching | | |
| --- | --- | --- | --- | --- | --- | --- |
| Characteristics | Treated | Control | d | Treated | Control | d |
| N | 55098 | 27952 |  | 43637 | 26320 |  |
| Age (5-year bins) |  |  |  |  |  |  |
| [16,21) | 401 ( 0.7) | 560 ( 2.0) | -0.110 | 567.6 ( 1.3) | 342.4 ( 1.3) | 0.000 |
| [21,26) | 782 ( 1.4) | 1082 ( 3.9) | -0.153 | 1123.4 ( 2.6) | 677.6 ( 2.6) | 0.000 |
| [26,31) | 1137 ( 2.1) | 1333 ( 4.8) | -0.149 | 1475.8 ( 3.4) | 890.2 ( 3.4) | 0.000 |
| [31,36) | 1703 ( 3.1) | 1770 ( 6.3) | -0.153 | 2064.7 ( 4.7) | 1245.3 ( 4.7) | 0.000 |
| [36,41) | 2571 ( 4.7) | 2319 ( 8.3) | -0.148 | 2898.7 ( 6.6) | 1748.3 ( 6.6) | 0.000 |
| [41,46) | 3335 ( 6.1) | 2683 ( 9.6) | -0.132 | 3551.7 ( 8.1) | 2142.3 ( 8.1) | 0.000 |
| [46,51) | 4243 ( 7.7) | 2792 (10.0) | -0.081 | 4110.0 ( 9.4) | 2479.0 ( 9.4) | 0.000 |
| [51,56) | 5301 ( 9.6) | 3113 (11.1) | -0.050 | 4823.6 (11.1) | 2909.4 (11.1) | 0.000 |
| [56,61) | 6107 (11.1) | 3105 (11.1) | -0.001 | 5182.9 (11.9) | 3126.1 (11.9) | 0.000 |
| [61,66) | 6560 (11.9) | 2812 (10.1) | 0.059 | 5001.4 (11.5) | 3016.6 (11.5) | 0.000 |
| [66,71) | 6134 (11.1) | 2136 ( 7.6) | 0.120 | 4155.5 ( 9.5) | 2506.5 ( 9.5) | 0.000 |
| [71,76) | 5050 ( 9.2) | 1524 ( 5.5) | 0.143 | 2995.3 ( 6.9) | 1806.7 ( 6.9) | 0.000 |
| [76,81) | 4480 ( 8.1) | 1122 ( 4.0) | 0.173 | 2337.9 ( 5.4) | 1410.1 ( 5.4) | 0.000 |
| [81,86) | 4104 ( 7.4) | 924 ( 3.3) | 0.184 | 2039.1 ( 4.7) | 1229.9 ( 4.7) | 0.000 |
| [86,91) | 2400 ( 4.4) | 510 ( 1.8) | 0.147 | 1054.2 ( 2.4) | 635.8 ( 2.4) | 0.000 |
| [91,96) | 693 ( 1.3) | 137 ( 0.5) | 0.083 | 233.9 ( 0.5) | 141.1 ( 0.5) | 0.000 |
| [96,101) | 95 ( 0.2) | 29 ( 0.1) | 0.018 | 21.2 ( 0.0) | 12.8 ( 0.0) | 0.000 |
| [101,106) | 2 ( 0.0) | 1 ( 0.0) | 0.000 | 0.0 ( 0.0) | 0.0 ( 0.0) | 0.000 |
| [106,111) | 0 ( 0.0) | 0 ( 0.0) | 0.000 | 0.0 ( 0.0) | 0.0 ( 0.0) | 0.000 |
| Sex |  |  |  |  |  |  |
| M | 22645 (41.1) | 11132 (39.8) | 0.026 | 17146.8 (39.3) | 10342.2 (39.3) | 0.000 |
| F | 32453 (58.9) | 16820 (60.2) | -0.026 | 26490.2 (60.7) | 15977.8 (60.7) | 0.000 |
| Vaccination status |  |  |  |  |  |  |
| 1/1, >90d | 2 ( 0.0) | 0 ( 0.0) | 0.009 | 0.0 ( 0.0) | 0.0 ( 0.0) | 0.000 |
| 1/1, <14d | 12 ( 0.0) | 5 ( 0.0) | 0.003 | 0.0 ( 0.0) | 0.0 ( 0.0) | 0.000 |
| 1/1, 14-90d | 5 ( 0.0) | 13 ( 0.0) | -0.022 | 0.0 ( 0.0) | 0.0 ( 0.0) | 0.000 |
| Not vaccinated | 51111 (92.8) | 26612 (95.2) | -0.103 | 42388.8 (97.1) | 25567.2 (97.1) | 0.000 |
| 1/2, >90d | 22 ( 0.0) | 6 ( 0.0) | 0.011 | 0.0 ( 0.0) | 0.0 ( 0.0) | 0.000 |
| 1/2, <14d | 1121 ( 2.0) | 419 ( 1.5) | 0.041 | 441.6 ( 1.0) | 266.4 ( 1.0) | 0.000 |
| 1/2, 14-90d | 1791 ( 3.3) | 595 ( 2.1) | 0.069 | 688.0 ( 1.6) | 415.0 ( 1.6) | 0.000 |
| 2/2, >90d | 239 ( 0.4) | 61 ( 0.2) | 0.038 | 15.0 ( 0.0) | 9.0 ( 0.0) | 0.000 |
| 2/2, <14d | 242 ( 0.4) | 79 ( 0.3) | 0.026 | 32.4 ( 0.1) | 19.6 ( 0.1) | 0.000 |
| 2/2, 14-90d | 553 ( 1.0) | 162 ( 0.6) | 0.048 | 71.1 ( 0.2) | 42.9 ( 0.2) | 0.000 |
| Period |  |  |  |  |  |  |
| before 10.01.2021. | 32317 (58.7) | 16822 (60.2) | -0.031 | 27021.7 (61.9) | 16298.3 (61.9) | 0.000 |
| 10.01.-15.07.2021. | 22102 (40.1) | 10664 (38.2) | 0.040 | 16319.0 (37.4) | 9843.0 (37.4) | 0.000 |
| after 15.07.2021. | 679 ( 1.2) | 466 ( 1.7) | -0.036 | 296.3 ( 0.7) | 178.7 ( 0.7) | 0.000 |
| Weighted CC index |  |  |  |  |  |  |
| 0 | 23477 (42.6) | 16823 (60.2) | -0.357 | 24388.1 (55.9) | 14709.9 (55.9) | 0.000 |
| 1-2 | 21575 (39.2) | 8827 (31.6) | 0.159 | 16179.3 (37.1) | 9758.7 (37.1) | 0.000 |
| 3-4 | 7228 (13.1) | 1741 ( 6.2) | 0.235 | 2607.4 ( 6.0) | 1572.6 ( 6.0) | 0.000 |
| >=5 | 2818 ( 5.1) | 561 ( 2.0) | 0.168 | 462.2 ( 1.1) | 278.8 ( 1.1) | 0.000 |
| Comorbidities |  |  |  |  |  |  |
| Atrial fibrillation | 5687 (10.3) | 1347 ( 4.8) | 0.209 | 1912.5 ( 4.4) | 1153.5 ( 4.4) | 0.000 |
| Autoimmune | 10034 (18.2) | 4398 (15.7) | 0.066 | 6598.2 (15.1) | 3979.8 (15.1) | 0.000 |
| Cancer | 6253 (11.3) | 2070 ( 7.4) | 0.136 | 3008.4 ( 6.9) | 1814.6 ( 6.9) | 0.000 |
| Congestive heart failure | 4234 ( 7.7) | 851 ( 3.0) | 0.207 | 1026.7 ( 2.4) | 619.3 ( 2.4) | 0.000 |
| COPD | 9700 (17.6) | 3819 (13.7) | 0.109 | 5793.6 (13.3) | 3494.4 (13.3) | 0.000 |
| IHD or CVD | 11683 (21.2) | 2907 (10.4) | 0.299 | 5529.7 (12.7) | 3335.3 (12.7) | 0.000 |
| Renal disease | 2580 ( 4.7) | 468 ( 1.7) | 0.172 | 324.4 ( 0.7) | 195.6 ( 0.7) | 0.000 |
| Immunocompromised | 1873 ( 3.4) | 400 ( 1.4) | 0.128 | 357.4 ( 0.8) | 215.6 ( 0.8) | 0.000 |
| RAAS inhibitors | 23376 (42.4) | 6640 (23.8) | 0.405 | 14261.2 (32.7) | 8601.8 (32.7) | 0.000 |
| Death (composite) | 4356 ( 7.9) | 907 ( 3.2) | 0.204 | 2063.2 ( 4.7) | 973.5 ( 3.7) | 0.045 |
| Death 1 | 3195 ( 5.8) | 691 ( 2.5) | 0.168 | 1573.6 ( 3.6) | 775.1 ( 2.9) | 0.033 |
| Death 2 | 2839 ( 5.2) | 605 ( 2.2) | 0.160 | 1341.0 ( 3.1) | 657.8 ( 2.5) | 0.031 |
| Death 3 | 3670 ( 6.7) | 734 ( 2.6) | 0.193 | 1695.8 ( 3.9) | 772.1 ( 2.9) | 0.045 |
| Hospitalization 1 | 5824 (10.6) | 1536 ( 5.5) | 0.188 | 3142.3 ( 7.2) | 1475.2 ( 5.6) | 0.059 |
| Hospitalization 2 | 12735 (23.1) | 3771 (13.5) | 0.251 | 7415.8 (17.0) | 3935.5 (15.0) | 0.053 |
